# Evolution-guided association mapping using long-read genomes from a self-identified Black or African American cohort in *All of Us*

**DOI:** 10.64898/2026.11.03.26367195

**Authors:** Natthapon Soisangwan, Kiran V. Garimella, Michael C. Schatz, Evan E. Eichler, Fritz J Sedlazeck, Michael E. Talkowski, Fabio Cunial, Samuel K. Lee, All of Us Research Program Long Read Working Group, PingHsun Hsieh

## Abstract

**Background:** Identifying functional variants that influence human health remains a challenge in human genetics. Conventional genotype-phenotype association studies test millions of variants, imposing a substantial multiple-testing burden as genomic and biobank datasets expand in size and variant diversity. Additionally, the contribution of structural variation, which is a substantial driver of genetic diversity, remains underexplored in these studies. Signatures of recent positive selection offer a powerful biological basis for prioritizing potentially functional variation, although demographic history can confound their detection and interpretation in these analyses.

**Results:** We performed evolution-guided association mapping using long-read genomes from 1,027 self-identified Black or African American participants in the *All of Us* Research Program and short-read genomes from an additional 38,552 matched participants of African-ancestry. Using demography-calibrated inferences, we identified 228 genomic regions with local ancestry enrichment or depletion and 1,616 candidate loci showing putative recent positive selection signals. These loci were enriched for genic and regulatory sequences, as well as previously reported eQTL, sQTL, and GWAS associations.

Leveraging the resolution of the long-read data, we mapped 15 structural variants to these candidate loci, including 10 located within 1,000 base pairs of putative *cis*-regulatory enhancers. Admixture- and selection-based phenome-wide association testing identified 13 and 446 nominal associations, respectively, 17 of which remained significant following replication and meta-analysis.

**Conclusions:** Our results support the use of evolutionary signatures as a robust framework for uncovering clinically relevant variation, including structural variants, in underrepresented populations.

## Background

Mapping the functional consequences of genomic variation remains a central challenge in human genetics. Population-scale biobanks produce a large amount of whole-genome sequencing (WGS) and electronic health records (EHR) from millions of participants [1, 2]. The recent advances in long-read sequencing now allow us to access difficult genomic regions that were inaccessible by short-read sequencing data and resolve complex variations, including structural variants (SVs) [3]. SVs, such as deletions, insertions, inversions that affect ≥ 50 base pairs (bp), alter substantially more base pairs than single-nucleotide variants (SNVs) [4], are disproportionately enriched for expression quantitative trait loci (eQTLs) and are associated with many diseases [5]. Incorporating long-read sequencing, these large biobanks now unveil hundreds of thousands of previously unresolved variants for biobank-scale association testing. While this sheer amount of biobank data offers unprecedented opportunity to map genotype to phenotype, testing millions of segregating sites, of which the vast majority of which are likely evolutionarily neutral, imposes a severe multiple-testing burden that limits statistical power and obscures true biological signals and disease associations [6, 7].

Recent positive selection serves as a powerful biological prior for variant prioritization. Natural selection acts directly on fitness, which is inherently governed by phenotype. A variant that rapidly rose in frequency under the classic model of positive selection did so by conferring a distinct functional advantage [8]. Genome-wide evolutionary scans have a proven record of recovering high-impact candidates, such as *LCT* and *MCM6* variants conferring lactase persistence [9], amylase copy number expansions in agricultural populations [10, 11], and immunity-driven phenotypes, e.g., the Duffy-null allele selected under malaria pressure [12]. It is worth noting that alleles targeted by historical selection may have additional phenotypic effects in modern environments through pleiotropy or gene-by-environment interactions. Examples include the malaria-protective Duffy-null allele, which is also associated with lower benign neutrophil counts and can lead to neutropenia misclassification, clinical trial ineligibility, and inappropriate withholding of medications [13, 14].

More broadly, alleles that were advantageous under past selective pressures can become maladaptive in modern contexts or reveal disease trade-offs. Classic examples include the *HbS* allele, which protects heterozygotes against severe malaria but causes sickle cell disease in homozygotes [15], and *APOL1* risk variants, which protect against *Trypanosoma brucei* infection but substantially increase chronic kidney disease risk [16]. Similarly, disease trade-offs are seen for *G6PD*-deficiency variants, which are enriched in malaria-endemic regions but predispose carriers to hemolysis after oxidative exposures. Another example is the diabetes-related *SLC16A11* haplotype, which is common in Latin American populations and increases type 2 diabetes risk, likely through effects on lipid metabolism in modern nutritional environments [17, 18]. Taken together, we hypothesize that by leveraging these evolutionary footprints of selection as a direct filter, we can effectively prioritize variants targeted by past selection for association testing because they carry a higher prior for harboring relevant biological and clinical functionality in the genome.

It is equally important to recognize that leveraging selection as a prior requires distinguishing adaptive signals from demographic confounders. This is particularly challenging in recently admixed populations, where gene flow, local genetic drift, and rapid population expansion distort the allele frequency spectrum and generate patterns of linkage disequilibrium that easily mimic selective sweeps [19, 20]. Furthermore, admixture also presents a unique analytical opportunity because it allows us to detect loci where alleles carried on divergent ancestral backgrounds exhibit differing phenotypic effects [21]. To isolate adaptive evolution from background demography, evolutionary inferences must be calibrated against simulations that explicitly model complex population histories [22]. Here, we integrate evolutionary population genetics with biobank-scale data to systematically link putatively functional variants to phenotypic outcomes. We leverage both SNVs and SVs identified from a newly generated long-read sequencing cohort of 1,027 self-identified Black or African American participants from the NIH *All of Us* (AoU) Research Program [23]. We also use this resource as a high-resolution WGS imputation panel to project variants into an additional 10,000 AoU individuals with matched predicted ancestry and linked EHR data [1]. By cross-referencing demographic-aware selection scans with phenome-wide associations (PheWAS), we construct a comprehensive map of adaptive variation and assess how evolution continues to shape complex disease risk and health outcomes today.

## Results

### Demographic inferences of the 1,025 self-identified Black or African Americans

Our AoU cohort includes 1,025 self-identified Black or African American (AoU_AFA) participants, recruited from multiple sampling locations across the continental U.S., with state-level sample sizes ranging from ≤20 to 180 participants (**Fig. 1A**). We removed two related individuals (kinship>0.1) from the initial 1,027 samples from analysis. To understand the genetic relationship of the AoU_AFA sample with respect to the rest of AoU WGS participants in the U.S. (n=245,388, v8) [1] and the global panel of the 1000 Genomes Project (1KG) [24], we performed principal component (PCA) and ADMIXTURE analyses (**Methods**). Our PCA results showed that most individuals in the AoU_AFA cohort well overlap with genetically inferred African (AFR)-like individuals in the main AoU biobank, while a subset falls within the range of European (EUR)-like participants. (**Fig. 1B**). Similarly, the joint unsupervised ADMIXTURE analysis supports that AoU_AFA samples are predominantly composed of AFR-like ancestry, with a median EUR-like admixture fraction of approximately 16.8% (K=2, **Fig. 1C and Fig.**

**Figure 1.**
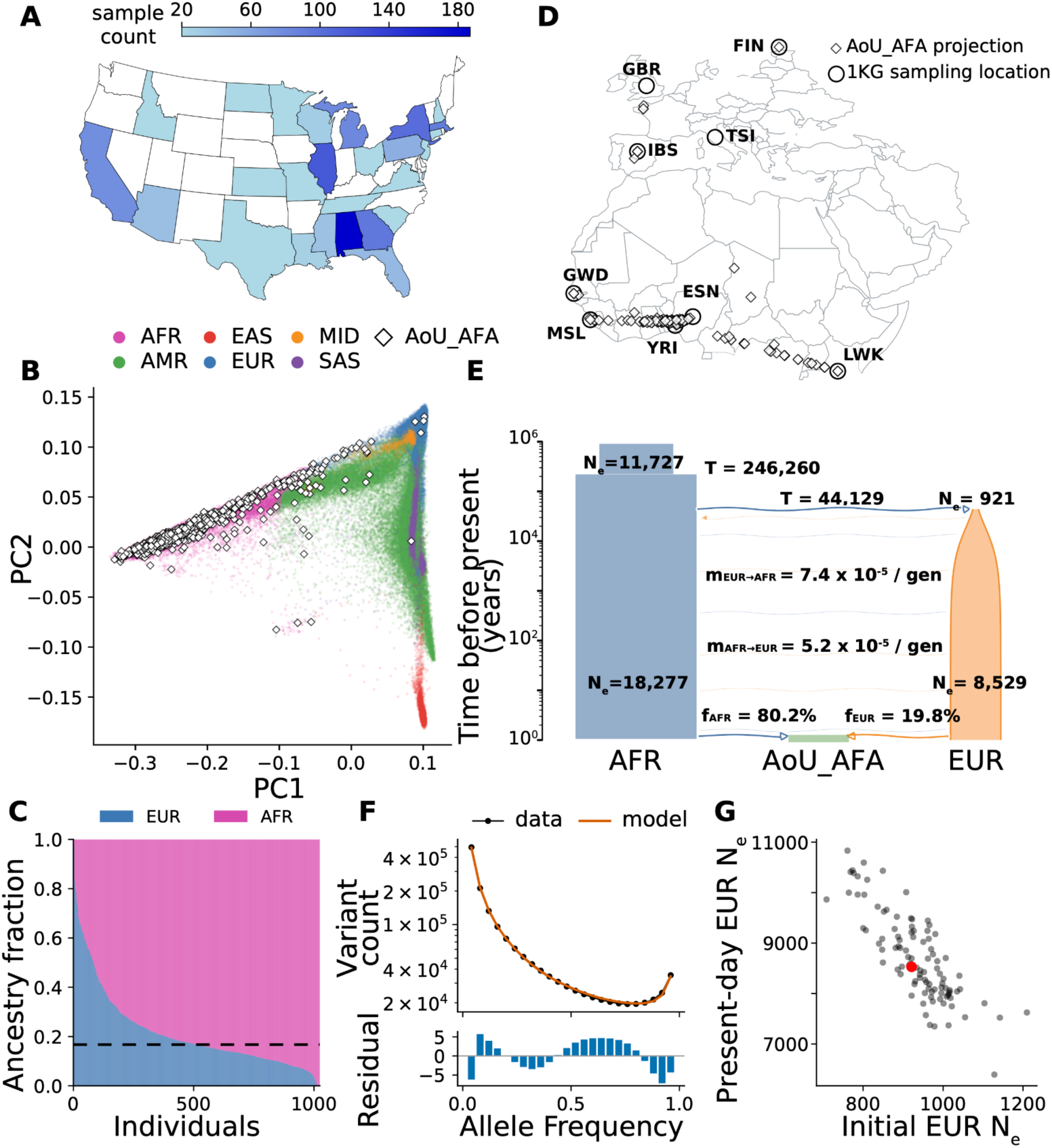
Demographic inference of 1,025 self-identified Black or African American AoU_AFA samples. **(A)** Sampling locations of the 1,025 AoU_AFA individuals by state. Counts <20 are shown as 20. (B) PCA of AoU_AFA individuals overlaid against the AoU CDR v8 cohort with predicted genetic ancestry. **(C)** Unsupervised ADMIXTURE analysis (k=2) of AoU_AFA individuals, with median admixture fraction of 17% EUR component indicated. Joint ADMIXTURE analysis for k=2-5 with 1000 Genomes AFR and EUR individuals are shown in Fig. S1. **(D)** Projected locations for AoU_AFA individuals with respect to 1000 Genomes AFR and EUR reference populations. Projection was computed from distance weighted geographic coordinates using the nearest 20 1000 Genomes samples and 16 variance-weighted PCs. **(E)** Inferred best fit demographic model for the AoU_AFA cohort jointly inferred with 1000 Genomes AFR and EUR individuals using a SFS method (∂a∂i). Bootstrap uncertainty and intermediate models are reported in Table S2 and Fig, S2. **(F)** Fit of the inferred model SFS compared to the empirical SFS, with residuals. **(G)** Model initial and final European effective population size is shown in red. Bootstrapping (n=100) was performed on the empirical SFS and re-optimized under the inferred model, with the best fit values for initial and final European deme effective population indicated. Bootstrapped pairwise uncertainty of all parameters is shown in Fig. S3.

**S1**). To further explore the diversity represented in the AoU_AFA at the global scale, we performed a PCA-based geospatial projection of these samples using the 1KG populations as a reference panel (**Methods**). We found that most of the AoU_AFA cohort is genetically spread throughout Central sub-Saharan Africa (**Fig. 1D**). Consistent with the findings of PCA and ADMIXTURE analysis, 2.34% (n=24) of our cohort share strong EUR-like components in their genomes and are geographically more affiliated with the 1KG European populations (**Fig. 1D**). These results together reveal a strong signal of historical admixture between EUR-like and AFR-like ancestries underlying the genetic diversity in the AoU_AFA cohort.

To better calibrate the evolutionary history of the AoU_AFA cohort for downstream analysis, we formally model their history used ∂a∂i [25], an allele-frequency spectrum based demographic inference method. We built a joint allele frequency spectrum using >22 million segregating intergenic SNVs after quality control filtering from our cohort and merging with 409 AFR and 418 EUR ancestry samples from the 1KG collection (Methods). Among the models we tested (**Fig. 1E, Figs. S2A-E, and Table S2**), our best-fit model (**Fig. S2E**, composite log-likelihood: −35,290.57) quantitatively reproduces the allele frequency spectrum and qualitatively fits the data (likelihood ratio test: adjusted *D* statistic=20.01, p=7.69×10^-6^, χ² degrees of freedom=1). Under the best-fit model, a population diverged from the ancestral population analogous to the Out of Africa event around 44,129 years ago (95% CI: 37,592– 53,392). This divergence results in the formation of a population with an effective population size of 921 (95% CI: 714–1,195) individuals, resembling the ancestral population of modern-day Europeans.

Following the divergence event, the effective population size of the EUR-like population grew exponentially (0.126% per generation) to 8,529 (95% CI: 7,078–10,867) in the present day. Consistent with the ADMIXTURE analysis, the AoU_AFA cohort comprises 80.2% (95% CI: 80.1–80.3%) AFR- like ancestry and 19.8% (95% CI: 19.7–19.9%) EUR-like ancestry. Finally, our best-fit model supports significant asymmetric gene flow between the present-day AFR-like and EUR-like populations (EUR-like to AFR-like: 7.4×10^-5^, 95% CI: 5.8×10^-5^– 9.4×10^-5^; AFR-like to EUR-like: 5.2×10^-5^, 95% CI: 4.0×10⁻⁵–6.3×10^-5^ per chromosome per generation). Our inferred model shares the core demographic events with the Gravel et al. (2011) demographic model [26], including comparable Out of Africa split times (44 kya vs. 51 kya). However, our model infers immediate exponential growth following the Out of Africa split rather than an extended bottleneck, along with a more modest European growth rate of 0.126% per generation compared to 0.38% per generation in Gravel et al. (2011) [26]. To further evaluate the quality of our demographic inference, we performed 100 nonparametric bootstraps of the intergenic data (**Methods**). Overall, the estimated parameters are well within the bootstrapping estimates, suggesting that our best-fit model is well-calibrated and accounts for sampling uncertainty within the data (**Fig. 1G, S2, and S3**).

**Figure 2.**
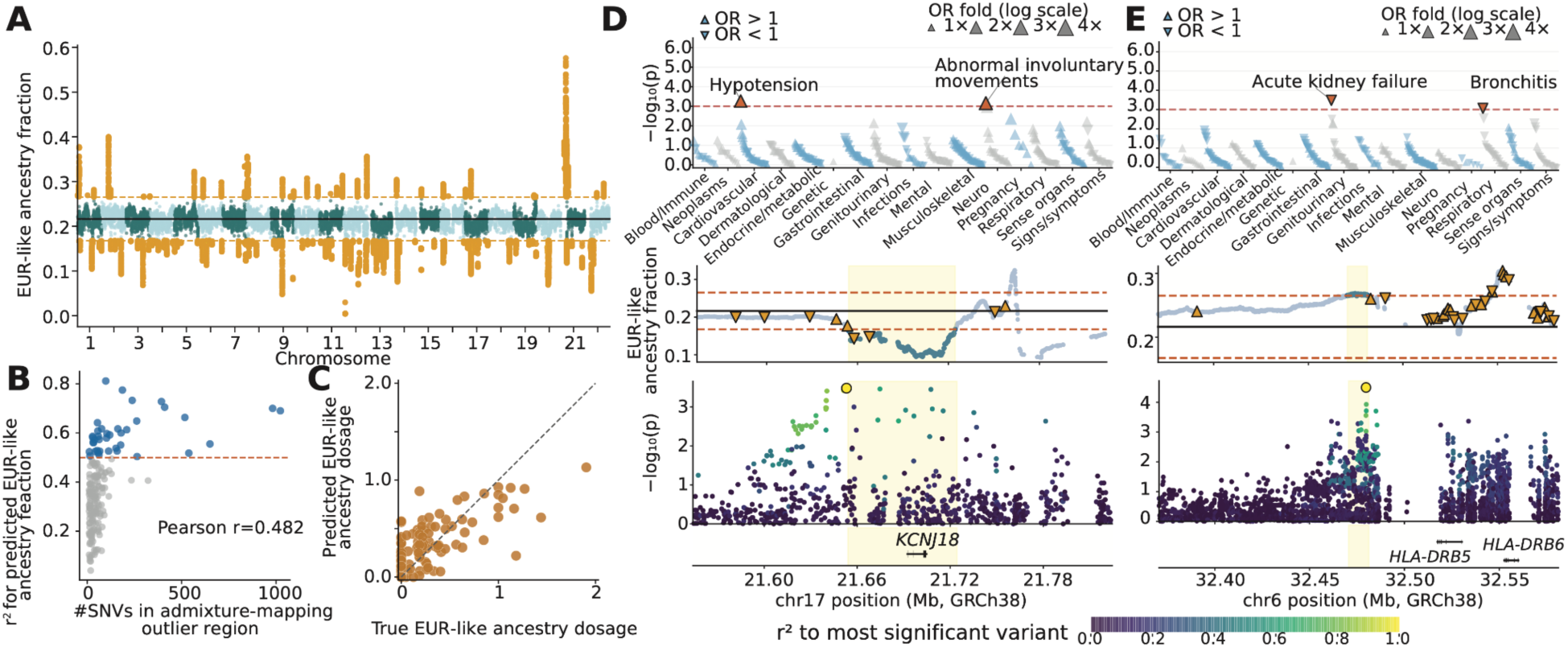
Admixture mapping-based PheWAS associations. **(A)** EUR-like ancestry fraction across the genome in AoU_AFA samples. Local ancestry was computed per shared marker using the 1KG AFR and EUR samples as the reference panel. Outlier loci (orange) with a EUR-like ancestry fraction at that locus of above or below the genome-wide mean ± 4.42 SD (0.167–0.265) are plotted. **(B)** Test performance across PLSR models to predict EUR-like ancestry dosage. Regions with model fit r^2^>0.5 are highlighted in blue. **(C)** Comparisons between observed and PLSR imputed EUR-like ancestry dosage for an outlier region (chr9:63,748,785-63,837,789) in 200 samples from the withheld test set in the AoU_AFA cohort. (**D**)&(**E**) Fine-mapping of PheWAS associations at the *KCNJ18* (chr17:21,654,025-21,724,463) and *HLA* (chr6:32,470,563-32,481,211, “acute kidney failure”) loci. Top panels: PheWAS manhattan plots across 447 phecodes in 16 categories. Upward and downward triangles represent increased and decreased odds- ratios for individual phecode outcomes, respectively. Middle panels: EUR-like ancestry fraction across markers. Outlier regions are indicated in yellow. Upward and downward triangles indicate insertion and deletion variants, respectively. Bottom panels: association signals of variants across individual loci. Color gradients of individual variants indicate linkage disequilibrium with respect to the lead test variants. Gene locations are indicated.

### Admixture-mapping analysis in the AoU_AFA cohort

Genetically admixed population cohorts provide an opportunity to test for associations between local ancestry and complex traits [27]. To systematically explore the contribution of EUR-like and AFR-like local ancestry in the *All of Us* cohort to complex traits, we performed a local ancestry scan and two-stage PheWAS in the AoU Biobank using AFR-like samples (**Methods**). Importantly, our objective was to investigate broader health outcomes since African Americans often carry a disproportionately high burden for disease prevalence. To infer marker-level local ancestry, we applied FLARE v0.6 [28] to the AoU_AFA individuals with the 1KG AFR and EUR reference panels (**Methods**) (**Fig. 2A**). We merged consecutive markers to identify 228 local ancestry outlier regions, defined as loci where local ancestry deviates by >4.42 standard deviations from the genome-wide mean (mean EUR-ancestry estimate = 21.6%; outlier thresholds: <16.7% or >26.5%, corresponding to a genome-wide significance of p < 1.0 × 10^-5^ in African Americans) [29]. These outlier regions range from 3.6 to 761 kbp in length and encompass a mean of 0.49 genes per region (range: 0 – 4). To improve the performance of association mapping, we built partial least squares regression (PLSR) models for each outlier region to predict the expected EUR-like ancestry dosage and applied the trained PLRS models to a discovery set of 10,000 ancestry-matched AFR-like imputation cohort (**Methods**) [23]. With an 80:20 training vs. testing split, we found that the performance of our PLRS models was highly correlated (Pearson’s r = 0.48) with the number of SNVs in individual regions (**Fig. 2B**). Using a prediction r^2^ cutoff of 0.5 (**Fig. 2B and 2C**), we selected 39 admixture outlier loci for association mapping.

To detect local-ancestry specific associations, we tested 447 phecodes with at least 200 cases and 200 controls in the discovery cohort for each region, using the predicted EUR-like ancestry dosage as the predictor along with age, age^2^, sex, and 20 PCs from a within-population PCA as covariates. We identified 13 associations at a nominal discovery significance threshold of p = 1.0 × 10^-3^ involving nine outlier regions (**Table S3**). Three loci (chr17:21,654,025-21,724,463, **Fig. 2D and S17**; chr6:32,470,563-32,481,211, **Fig. 2E and Fig. S13**); chr21:8,756,876-9,169,773, **Fig. S18 and S19**) contain distinct associations in different phecode categories, suggesting pleiotropic effects. For example, the region chr17:21,654,025-21,724,463 is depleted of EUR-like ancestry (mean EUR-like ancestry=12.73%), and has nominal associations for increased risk of “hypotension” (p = 5.4 × 10^-4^; β = 0.685, SE =0.20) and “abnormal involuntary movements” (p = 7.0 × 10^-4^; β = 0.792, SE=0.23) with increasing EUR-like dose (**Fig. 2D**). These results suggest that higher AFR-like ancestry may be protective against hypotension risk, which is interesting given that African Americans carry a lower prevalence for orthostatic hypotension (7.5%) than whites (14.5%) [30], but a high prevalence for hypertension (59%) [31]. Located in the EUR depletion region is *KCNJ18*, a potassium channel gene primarily expressed in skeletal muscle [32] and involved in hypokalemic periodic paralysis, which may partially account for the hypotension and abnormal involuntary movement associations found. In addition, the chr6:32,470,563–32,481,211 locus near *HLA-DRB5* is enriched for EUR-like ancestry (mean EUR-like ancestry=26.67%), but shows reduced risk for acute kidney failure (p = 3.3 × 10^-4^; β = −0.402, SE = 0.11) and bronchitis (p = 8.59 × 10^-4^; β = −0.372, SE = 0.11) with increasing EUR-like dose (**Fig. 2E**). *HLA* is suggested to be a risk loci for renal function [33], and *HLA-DRB5* is upregulated in interstitial lung disease [34]. Across our admixture associations, SVs do not appear to be driving the associations, with no SVs as the lead variant and low linkage of SVs (r^2^ > 0.5) to the lead fine-mapped variants. Finally, we attempted to replicate all 13 nominal associations in a replication cohort of 28,552 AFR-like AoU samples from short read data. Among these, nine associations were not testable in the PLSR models due to sparsity of SNVs in the replication cohort (**Table S3**). One association for chr9:63,748,785–63,837,789 was significant in meta-analysis for “acute pain” (β_meta_ = 0.449, SE = 0.165, p_meta_ = 3.1 × 10^-3^) (**Fig. S16**).

### Genome-wide identification of selection candidates

To detect potential signals of adaptation, we computed linkage-based integrated haplotype score (iHS) [35] at individual variants and recorded the maximum absolute normalized iHS score within each non-overlapping 20 kilo-base pairs (kbp) window across the autosomes. We evaluated significance of selection using 1,000 coalescent simulations incorporating our best-fit demographic model (**Fig. 1E**) and mutation and recombination rate heterogeneity across the genome (**Methods**). To account for model uncertainty in our simulations, demographic parameters were jointly drawn from the bootstraps using a multivariate sampling approach (**Methods**). We showed that the genome-wide mutation rate estimates from our simulations match those observed from the empirical data (Pearson r=0.98, **Fig. S4**). This suggests our approach is likely more robust to potential biases from variation in mutation rate and recombination rate across the genome [36].

At a significance threshold of simulation p ≤ 0.01, we identified 2,345 significant windows as putative selection targets (**Fig. 3A**). We observed a genome-wide left-skewed p-value distribution from our analysis, indicating a conservative inference of selection (**Fig. S5A**). A Quantile-Quantile comparison of our p-value distribution against an expected uniform distribution confirms that our approach is well-powered to detect putative targets of selection (**Fig. 3B**). Overall, we found that these candidate windows exhibit larger absolute iHS values (|iHS|) and a higher alternative allele frequency (median alternative allele frequency=22.7%) than the non-candidate windows (median alternative allele frequency=17.4%, p=7.2×10^-27^, one-sided Mann-Whitney U) (**Fig. S5B and S5C**). Notably, our simulation-based approach captured a distinct set of signals compared to a simple empirical ranking. Among the candidate windows, 1,287 significant windows fell outside the top 1% genome-wide empirical |iHS| values (Quadrant IV, **Fig. S6**), while 171 windows within the empirical top 1% were not significant in our simulations (Quadrant II, **Fig. S6**).

**Figure 3.**
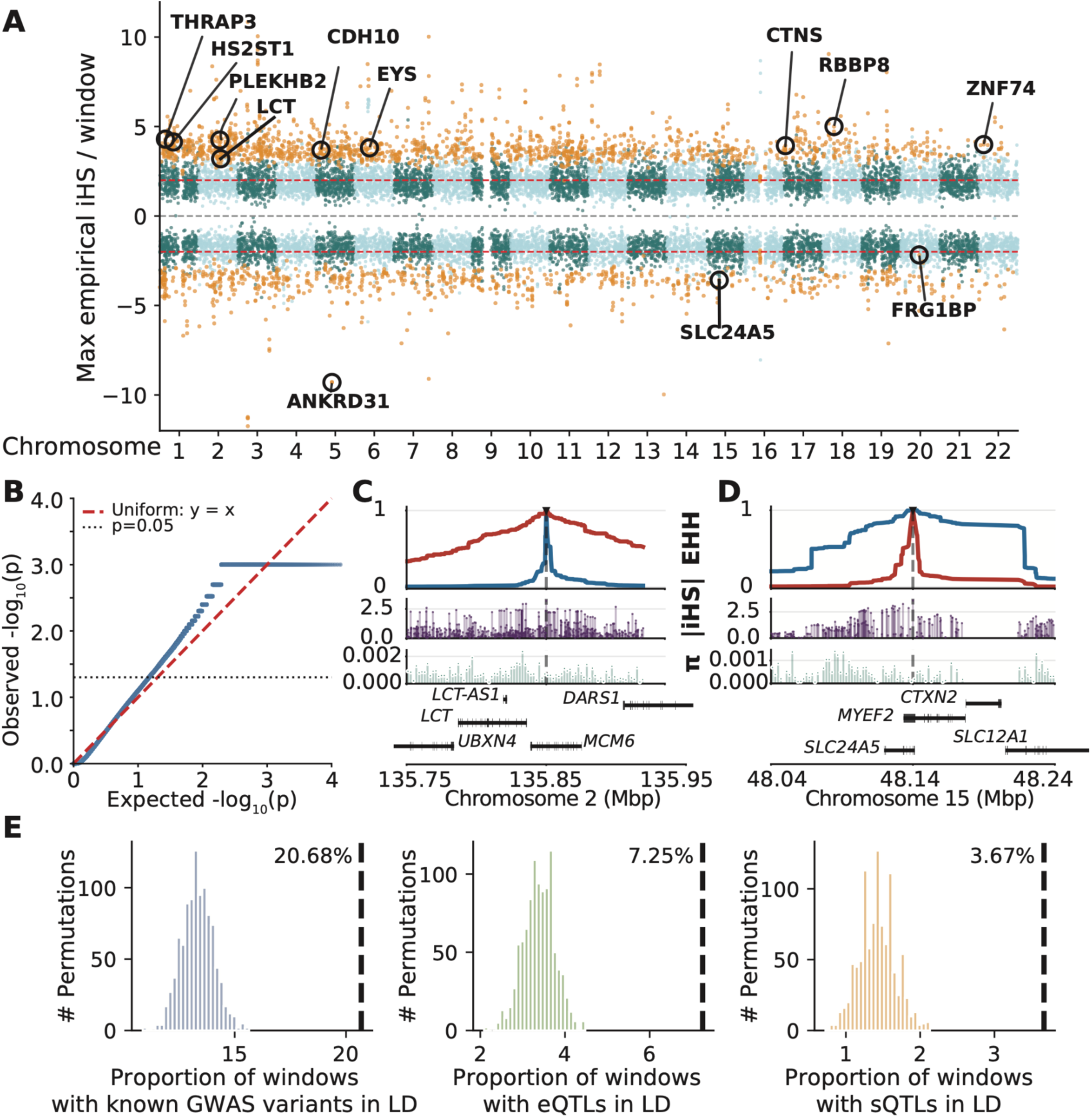
Selection across the genome enriches for gene regulation and trait QTLs. **(A)** iHS selection scan and simulation-calibrated genome wide hits, comprising a set of selected loci (n=2,345 at p≤0.01). A subset of genic loci in the candidate set is indicated. **(B)** Q-Q plot of expected and simulated p values across the genome. The minimum simulated p-value is −log_10_(1/1001) ≈ 3. **(C)** Extended haplotype homozygosity (EHH) of rs4988235 G>A for haplotypes carrying reference (G, blue) and alternate (A, red) alleles. Peak |iHS| and nucleotide diversity (π) per window in the region are shown. rs4988235 is located in a *MCM6* intron. **(D)** Extended haplotype homozygosity (EHH) of rs2470102 A>G for haplotypes carrying reference (A, blue) and alternate (G, red) alleles. rs2470102 is located in a *SLC24A5* intron. **(E)** Permutation testing for colocalization between selection and known association loci in GWAS and MAGE catalog. 1000 permutations were drawn from 2,345 random 20 kbp selected windows, where the lead iHS loci and linked loci (r^2^>0.5) were queried against the GWAS (p<1×10^-8^) or MAGE catalog (eQTL or sQTL in any causal set) for known loci. The empirical proportion of colocalized loci for each category is indicated. Selected loci show significant enrichment (p<0.001) for GWAS loci (1.6-fold), eQTLs (2.1-fold), and sQTLs (2.6-fold) in the candidate set compared to the median of permuted draws.

### Enrichment of selection candidates in genes and known GWAS signals

The 2,345 significant windows correspond to 1,616 distinct loci across the genome (length range: 20 to 160 kbp), spanning 46.9 million base pairs (Mbp) or 1.6% of the autosome. Among these, 963 loci overlap with UCSC RefGene genes (**Table S5**). Because false-positive signals are not expected to preferentially associate with genes, we evaluated the functional relevance of our candidate loci using a permutation test for gene enrichment (**Methods**). We observed significant enrichment for genic windows compared to intergenic windows (1.08-fold, permutation p<0.001, **Fig. S7A**). Such enrichment was also observed for both exonic windows (1.15-fold, permutation p<0.001, **Fig. S7B**) and intronic windows (1.08-fold, permutation p<0.001, **S7C**). Furthermore, we evaluate genic windows for gene constraint using probability of loss intolerance (pLI) scores and ENCODE4 candidate *cis*-regulatory elements (cCRE) [37, 38]. Among the genic candidate loci, we found no significant deviations from the null expectation for overlap with ENCODE4 cCREs (20.4% of the genic loci, permutation p=0.20, **Fig. S8A**) or gene constraint (mean probability of loss intolerance [pLI] in candidate loci=0.33, permutation p=0.29, **Fig. S8B**) or. However, our Gene Ontology analysis [39] revealed that these loci are significantly enriched in 17 out of 600 molecular functions tested, including catalytic (1.26-fold, p=2.5×10^-4^) and transferase (1.40-fold, 5.8×10^-4^) activities (**Table S7**).

Given the observed genic enrichment of our selection candidates, we further performed a colocalization analysis to identify overlapping signals between our lead iHS variants and previously reported association signals (**Methods**). We compiled published eQTL and sQTL credible variants from the MAGE database [40] and genome-wide significant GWAS variants from NHGRI-EBI GWAS catalog (v1.0.2) [41]. Compared to windows falling outside the candidate set, we found enrichment of GWAS signals, eQTL signals, and sQTL signals in the candidate set (permutation *P*<0.001) (**Fig. 3E**). Overall, 299 of the 963 genic candidates were linked to at least one eQTL (n=84), sQTL (n=46), or GWAS signal (n=241) (**Table S6**). 52 loci were linked to both regulatory (eQTL/sQTL) and GWAS signals, suggesting that selection may act on phenotypes through gene regulation at these variants. Among these, we recovered several well-characterized targets of selection (**Table S6**), including the European lactase-persistence variant rs4988235 in an *MCM6* intronic enhancer that increases lactase activity [9], and the *CCL5* variant rs2107538, which is associated with reduced HIV infection risk in African populations (p=0.0002) [42] and protection against SARS-CoV-2 infection (p<0.001) [43]. We also identified *LPA* variants, rs6455695 associated with lipoprotein(a) levels (p=5×10^-26^) and rs6905073, which is associated with coronary artery disease (p=2×10^-10^), angina (p=3×10^-13^), and the ratio of polyunsaturated fatty acids to total fatty acids (p=7×10^-14^). Together the observed gene enrichment and colocalization with known association signals in our results highlight the ability of our evolutionary framework to identify biologically meaningful signals.

### Selection targets and SV contributions to selection

Our analysis recovered both previously reported selection candidates and novel loci. For example, one of the candidates overlaps with rs4988235 (chr2:135851076 G>A), a regulatory variant in *MCM6* that drives lactase persistence by increasing *LCT* expression [44]. Around rs4988235, we observed a slow decay of extended haplotype homozygosity (EHH) on haplotypes carrying the derived allele A (**Fig. 3C**) and elevated |iHS| values (iHS=3.20) at the locus, with the A allele at 13.4% frequency in AoU_AFA, 53.5% in 1kg EUR, and 2.4% in 1kg AFR. Consistent with a classic selection model, we found a reduction in nucleotide diversity (π) around this variant (**Fig. 3C)**. Similarly, we identified a candidate at *SLC24A5*, which carries rs2470102 (chr15:48141297 A>G, iHS=-3.58). The derived allele A under selection is known to be associated with lighter skin pigmentation [45] and eye color [46] across human populations. The rs2470102 A allele frequency in AoU_AFA is 23.2%, contrasting with 99.4% in 1kg EUR and 6.8% in 1kg AFR. Haplotypes that carry rs2470102 A exhibit much longer homozygosity and reduced diversity than those with allele G at this locus (**Fig. 3D**). Furthermore, we identified a selection variant rs28470942 (chr4:99103864 C>T, alternative allele frequency=18.1%) in the *ADH* gene cluster. Notably, rs28470942 is strongly linked (r^2=^0.87) to rs2066702 (*ADH1B*3* or chr4:99307860 G>A), which encodes an isoform with increased alcohol dehydrogenase activity [47], and is found at 19.3% frequency in the 1KG AFR populations. The *ADH* locus has recurrently shown signals of selection [48, 49], with proposed selective pressures including agricultural and dietary changes [50] and reduced adverse effects of prenatal alcohol exposure [51]. We also identified candidates with high allele frequency, such as rs2190227 (chr7:90,206,732 G>A, iHS=2.85, alternative allele frequency=93.6%) in *STEAP2*, a prostate growth mediator [52].

The long-read data from our AoU_AFA cohort enabled us to explore the contribution of SVs to recent positive selection. Within the 963 candidate loci, we identified 18 SV-lead associations that are either lead iHS variants or in strong linkage disequilibrium (r^2^ > 0.5 within 100 kbp) with lead iHS SNVs over 15 unique SVs (allele frequency range: 5.7–81.2%) (**Table S5, Fig. S38-S52**). SVs are significantly enriched by selection (2.89-fold enrichment, 15 unique SVs observed vs mean 5.19 SVs observed in permutations, p<0.001, **Fig. S9**). These comprise 13 insertions (eight intronic, five intergenic, length range: 50 to 4,455 bp) and two deletions (one intronic, one intergenic, length range: 52 to 267 bp). Of the 9 genic SVs where an SV is the putative lead variant, nine genes are tolerant to loss-of-function mutations (pLI=0.00–0.54), with the exception of the highly constrained *THRAP3* (pLI=1.0) (**Table S5**). The 52-bp intronic deletion at *THRAP3* (rs55751877, AF=81.2%) removes one of two 52-bp tandem repeats **(Fig. S39 and Table S11)**. *THRAP3* is an RNA processing factor that promotes the DNA response pathway and variants within it are reported associated with nonalcoholic fatty liver disease when overexpressed [53]. The largest iHS lead SV we identified is a 4,455-bp partial endogenous retrovirus insertion in an intron of *ANKRD31* (AF=78.4%, **Fig. S46**). *ANKRD31* is a double-strand break mediator during meiosis, and variants in this gene are known associated with male and female infertility [54, 55] and neurodevelopmental disorders, with a candidate variant for Rett syndrome [56]. Furthermore, we find a 90-bp insertion (**Fig. S52**) at allele frequency 9.3% in *ZNF74*, a gene which is frequently found deleted in 22q11.2 deletion syndrome (DiGeorge syndrome) and is a candidate gene for affecting age-at-onset of schizophrenia [57].

Colocalization with published eQTLs and GWAS signals further supports the functional relevance of several candidate SVs (**Table S6**). For example, we found the 72-bp insertion at *HS2ST1* (**Fig. S40**) is in strong linkage (r^2=^0.56) to a known eQTL (rs1621247) [40]. *HS2ST1* is essential for heparan sulfate sulfonation and kidney formation [58]. Similarly, a 135-bp insertion at *RBBP8* (**Fig. S51**) is linked to rs7236869, a variant associated with eosinophil counts (p=3.0×10^-14^) [59], while the 985-bp TcMar-Tigger DNA transposon insertion (**Fig. S45 and Table S12)** at *CDH10* is in strong linkage (r^2=^0.74) with rs10942058, a variant associated with educational attainment (p=3.0×10^-10^) [60]. Notably, spatial overlap within 1 kbp of ENCODE candidate *cis*-regulatory elements (cCREs) suggests that most these SVs (10 of 15) possess expression-modulating potential (**Table S14**), including *RBBP8*, *HS2ST1*, *ANKRD31*, and *ZNF74*.

### PheWAS of selected loci

To systematically explore the phenotypic impact of the selection candidates, we conducted a two-stage PheWAS in the AoU Biobank using AFR-like samples. Similar to the admixture analysis, we take 10,000 genetically predicted AFR samples in All of Us that contain EHR data, age and sex covariates, and 20 PCs for PheWAS analysis. (**Methods**). Importantly, our objective was to investigate the broader health outcomes associated with these variants, rather than the specific traits originally under selection. Exact participant counts for categories <20 cannot be shown as per the *All of Us* Data and Statistics Dissemination Policy and are instead shown as 20. In the discovery stage, using all variants discovered from the 1,025 AoU_AFA samples as the reference, we imputed all variants into a cohort of 10,000 ancestry matched AFR-like short-read samples in the AoU (**Methods**). We tested 447 phecodes with at least 200 cases and 200 controls against our candidate set of 963 variants (**Table S5**). At a nominal discovery significance threshold of p=1.0×10^-3^, we identified 446 associations involving 272 unique phecodes across 263 variants (**Fig. 4A**). Consistent with previous findings [61, 62], we found an inverse correlation between variant effect size and allele frequency (**Fig. 4B**).

**Figure 4.**
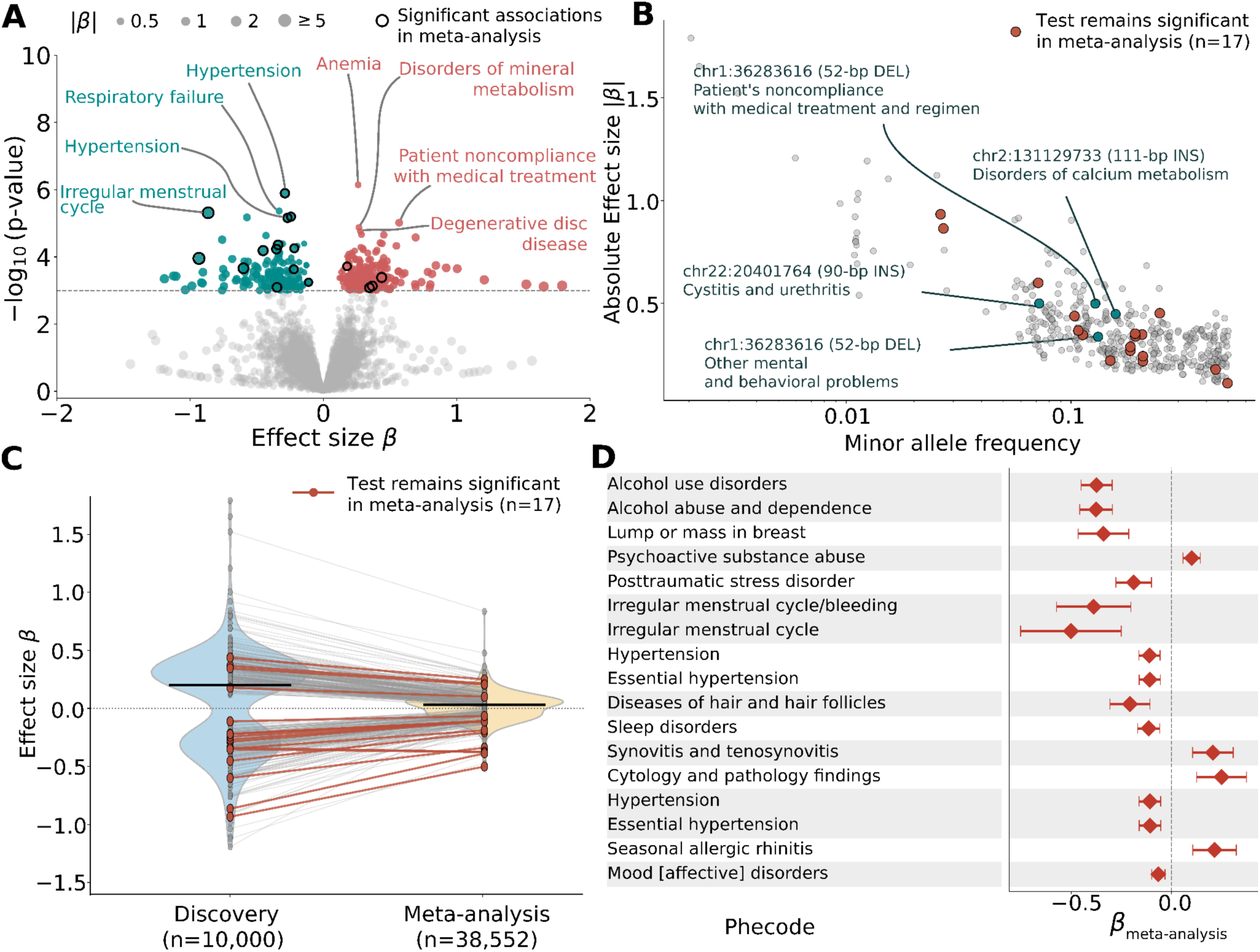
PheWAS on candidate set of selected loci. **(A)** Discovery effects found in the set of selected loci. A subset of discovered hits are indicated. At a discovery threshold of p<1×10^-3^, 446 genic associations involving 272 unique phecodes across 263 variants were found. **(B)**. Discovery effect sizes and minor allele frequency of each association is shown. Four associations involve loci that are linked to three unique SVs (r^2^>0.5 within 100 kb). After replication testing and inverse variance weighted meta-analysis, we find 17 meta-significant effects after Bonferroni correction (p < 1.11×10^-4^). **(C)** Discovery and meta-analyzed effect sizes. Discovery effects show attenuation after meta-analysis, with meta-effect sizes of approximately 21.9% compared to initial discovery. **(D)** 17 meta-significant associations and meta-analysis effect sizes, with effect sizes encoded as the alternate allele dosage. Associations are grouped by variant and sorted by significance (most significant first). Correlation between closely related phecodes may contribute to discovery of similar associations within variants.

We then performed replication analysis for these associations in an independent cohort of 28,552 short-read AFR-like AoU samples (**Methods)**. We also attempted to isolate the specific variants driving these associations using fine-mapping (**Methods**); however, the extended haplotype homozygosity resulting from positive selection creates extensive linkage around these loci, inherently confounding fine-mapping algorithms. In our analysis, we thus chose to report the lead index variant for each locus as the best available candidate for downstream evaluation (**Table S9**). Following inverse-variance weighted meta-analysis (**Methods**), 17 phecodes across 13 variants remained highly significant (Bonferroni p<1.1×10^-4^, **Fig. 4C and Table S8**). This is consistent with the action of selection against variants with large effects [63]. In addition, we found that effect sizes of variants significant in the meta-analysis attenuated to approximately 21.9% of those observed in the discovery cohort (**Figs. 4C and S10**), consistent with ascertainment bias in genetic association studies or differences between cohorts [64, 65].

The 13 variants significant in meta-analysis are associated with diverse phecodes (**Figs. 5A-C and S23-S37**), including five represented in the PheWAS mental category (alcohol use disorders, alcohol abuse and dependence, psychoactive substance abuse, posttraumatic stress disorder, mood disorders), and 16 phecodes in the 15 other categories (**Fig. 4D**). Across the meta-significant associations, the 13 putatively selected alleles are associated with decreased risk in eight cases (61.5%), and associated with increased risk in 5 cases (38.5%). One variant in *LINC01950,* rs73205915 (chr5:106,987,252 C>T), exhibits possible pleiotropic effect with an increased risk for “synovitis and tenosynovitis” but a nominally decreased risk for “diseases of esophagus” (**Fig. S31**). The variant rs2190227 (chr7:90,206,732 G>A) at the *STEAP2* locus showed the largest effect size for irregular menstrual cycle (β_meta_=-0.501, SE=0.128, p=9.1×10^-5^), with the putative selected alternative A allele associated with decreased risk (**Fig. S32**). rs2190227 fine-maps to a variant in *CFAP69* (chr7:90,254,854 A>G, rs17867598) with a PIP=0.03 (β=1.51, SE=0.357, p=2.1×10^-5^, alternative allele frequency=1.3%). rs17867598 is primarily found in European populations (1KG EUR frequency 3.5%, AFR frequency 0.6%). *CFAP69*, a cilia and flagella-associated protein, is highly expressed in the testes and fallopian tubes [66], and loss-of-function variants in this gene result in sperm morphology defects and dysfunction [67].

**Figure 5.**
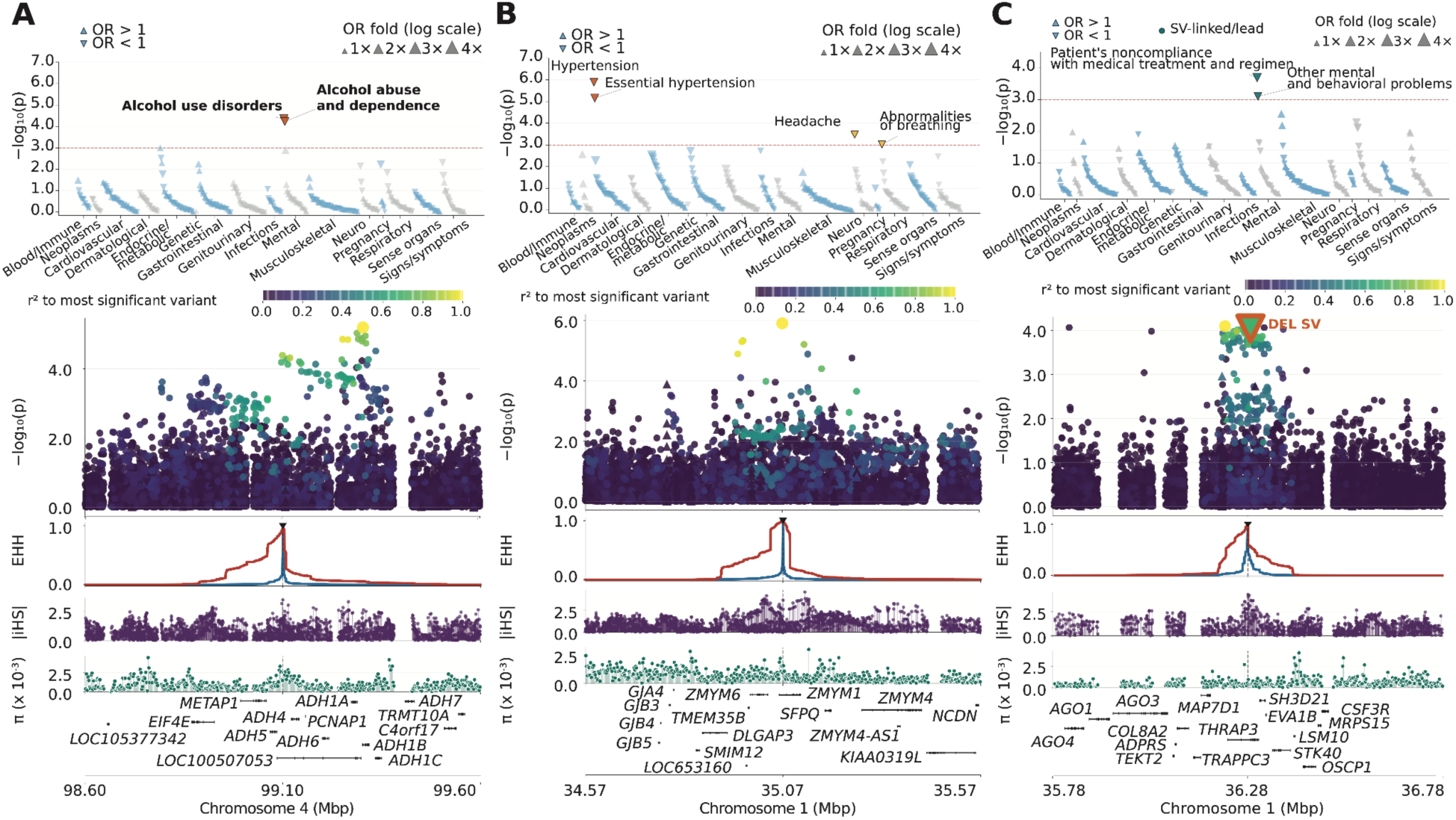
Fine-mapping of PheWAS associations at the (A) *ADH* (rs28470942, chr4:99103864 C>T), (B) *ZMYM1* (rs11264001, chr1:35,070,852 C>A), and (C) *THRAP3* (52-bp deletion at chr1:36,283,616 & rs2358719, chr1:36,277,076 C>G) loci. Top panels: PheWAS manhattan plots across 447 phecodes in 16 categories. Upward and downward triangles represent increased and decreased odds-ratios for individual phecode outcomes, respectively. Red and yellow colors are associations significant in discovery and meta-analysis, respectively. Second panels: association signals of variants across individual loci. Color gradients of individual variants indicate linkage disequilibrium with respect to the lead test variants. Third panels: extended haplotype homozygosity (EHH) centered at the lead test variants. Blue and red colors indicate haplotypes carrying reference and alternative alleles, respectively. Fourth to sixth panels: the absolute iHS scores, nucleotide diversity (π), and gene IDs, respectively, at individual loci.

At the *ADH1B* locus, fine-mapping recovers the missense variant rs2066702 (*ADH1B*3*) as a functional candidate with the highest posterior inclusion probability (PIP=0.06) associated with “alcohol use disorder” (β=-0.336, SE=0.074; p=6.3×10^-6^, alternative allele frequency=19.7%, **Fig. 5A**). This variant is exclusive to Africans (1KG AFR allele frequency 19.3%, EUR allele frequency 0.0%), with the direction of selection consistent with decreased risk. The *ADH1B*3* allele facilitates rapid ethanol oxidation and has been previously associated with reduced drinking behavior, protection against alcohol dependence, and/or lowered risk of alcohol-related birth defects in African Americans [51, 68, 69]. At *ZMYM1*, we find two variants (rs11264100, chr1:35,126,025 A>G; rs11264001, chr1:35,070,852 C>A, both with associations for “hypertension” and “essential hypertension”, with selection on the protective alternative allele (**Fig 5B and S23-S25**). Fine-mapping resolves these as one lead locus, rs11264001 (alternative allele frequency 21.96%, 1KG AFR=5.60%, 1KG EUR=88.78%), with similar effect sizes for hypertension (β=-0.288, SE=0.059; p=1.2×10^-6^, PIP=0.380) and essential hypertension (β=-0.269, SE=0.059, p=6.7×10^-6^, PIP=0.046). The protective allele is substantially lower in frequency in 1KG AFR than in 1KG EUR, of interest given the disproportionately high prevalence of hypertension among African Americans in the United States (59%) [31].

We further investigated phenotype associations involving SVs linked to selected SNVs. At the discovery stage, we find four nominal associations with a linked SV (r^2^ > 0.5) implicating three unique SVs (p_discovery_<1.0×10^-3^; **Table S10**). First, the 52-bp intronic deletion at chr1:36,283,616 at *THRAP3* (alternative allele frequency=81.2%) was linked to the tag allele rs2358719 (r^2=^0.52, chr1:36,277,076 C>G; iHS=4.25 AoU_AFA, alternative allele frequency=12.1%) (**Fig. S39 and S53**). The alternative allele G is almost exclusively found in African populations (1KG AFR alternative allele frequency=16.1%, EUR alternative allele frequency=0.0%), and is significantly associated with reduced risk for medical noncompliance (β_discovery_=-0.498, SE=0.134, p=1.9×10^-4^) and “other mental and behavioral problems” (β_discovery_=-0.337, SE=0.100, p=7.9×10^-4^). The alternative G allele of rs2358719 tags the non-deleted (reference) allele of this SV, suggesting selection for the non-deleted haplotype and increased risk of the deletion allele for these associated phenotypes. In addition, at *PLEKHB2*, a 111-bp insertion (chr2:131,129,733, alternative allele frequency=18.8%) at an intron of *PLEKHB2* is linked (r^2=^0.70) to SNV lead rs4850257 (chr2:131,094,186 A>G, iHS=4.25, AoU_AFA allele frequency=19.3%) (**Fig. S43 and S54**). The SNV is found more frequently in European populations (1KG AFR AF=6.8%, EUR AF=63.1%) and is linked to disorders of calcium metabolism (β_discovery_=0.448, SE=0.126, p=3.7×10^-4^). *PLEKHB2* is bound to recycling endosome membranes and regulates retrograde trafficking to the Golgi apparatus [70], and knockdown of *PLEKHB2* in a breast cancer cell line reduces proliferation rate [71]. Finally, our analysis revealed a 90-bp insertion (chr22:20,401,764, alternative allele frequency=9.3%) at *ZNF74* linked to rs76618551 (chr22:20,391,674 T>C; iHS=3.99, AoU_AFA allele frequency=6.2%) (**Fig. S52 and S55**). The insertion falls into the putative distal-like enhancer EH38E2150574, potentially affecting gene regulation through disruption of H3K27ac acetylation. rs76618551 is associated with cystitis and urethritis (β_discovery_=0.499, SE=0.151, p=9.2×10^-4^). The rs76618551 C allele is primarily found in African populations, with an AFR frequency of 6.9% in 1KG and is absent from all other populations except AMR (AF=0.8%). However, none of these SV-linked associations survived meta-analysis with the replication cohort. This lack of replication may be driven by subtle population structure differences, as the replication cohort exhibited a slightly different principal component analysis (PCA) composition compared to the discovery cohort (**Fig. S11**), potentially affecting the reproducibility of these specific effects.

## Discussion

In this study, we use recent positive selection as a biological prior to identify genetic variants affecting present-day phenotypes in the NIH All of Us self-identified African American cohort. A key strength of our framework is calibrating this selection scan using a realistic demographic model rather than traditional genome-wide empirical cutoffs. Such traditional cutoffs assume simplistic histories that can severely skew statistics in rapidly growing, admixed populations. Using simulations that incorporate neutral demographic forces and local variation in recombination and mutation rates, we restricted our subsequent phenome-wide analysis to these robustly identified loci, reducing the computational burden. This approach successfully prioritized functional variation, as evidenced by strong enrichment for genic sequences, regulatory elements, and diverse clinical diagnoses.

Typically, positive selection is expected to drive beneficial traits, such as lactase persistence [72] and adaptation to diet with consumption of fermented food and beverage [48] in certain populations. While 76% of our association signals with putatively selected alleles showing protective effects, including the reduced risk of alcohol use disorder associated with the *ADH1B*\*3 allele, we also identified instances of evolutionary mismatch, where historically adaptive alleles appear to confer increased disease risk in modern environments. For example, the rs2470102 allele A at the *SLC24A5* locus, which is likely selected for facilitating light skin pigmentation and vitamin D synthesis [73], reduces melanin production in the eye, a strong biomarker for ocular neoplasms [74]. Similarly, we observed an enrichment of inflammatory disease risks among recently selected loci, consistent with a known evolutionary tradeoff between historically beneficial immune hyper-responsiveness and modern autoimmune or inflammatory disorders [75]. These examples illustrate how alleles that increased fitness in past environments can become maladaptive as diet, lifespan, and pathogen exposures rapidly change.

Our framework highlights the critical role of SVs and regulatory elements as important drivers of phenotypic variation under selection. Because SVs alter substantial genomic base pairs and frequently occur in complex regions, they carry a strong prior for functional effects [4]. We identified 15 selection-linked SVs, including nine within genes and several overlapping or adjacent to ENCODE4 cCREs. This regulatory focus is further supported by our colocalization analysis, which revealed that nearly one-third of all genic candidate loci are linked to a GWAS association, eQTL, or sQTL. Notably, 52 loci linked to both a regulatory QTL and a GWAS hit, while we observed no significant shift in gene constraint (pLI) among the selected loci. Together, these observations demonstrate that recent selection could have frequently acted through regulatory changes, including those driven by SVs, rather than coding mutations [76]. This aligns with evolutionary expectations, as large-effect coding changes in constrained genes are typically deleterious, whereas regulatory alterations can beneficially tune gene expression in an environment-specific manner [77].

Several limitations should be considered. Our demographic model is a necessary simplification that cannot fully capture complex population substructure, and our reliance on iHS restricts our sensitivity to recent, incomplete sweeps [35]. Furthermore, the lack of SV-phecode replication highlights the limited statistical power and cohort-specific tagging differences inherent in imputing structural variants from short-read data [78]. Despite these challenges, our framework highlights the utility of integrating an evolutionary approach to biobank-scale analysis of trait and health outcomes.

## Conclusions

Our analysis establishes a practical framework for connecting evolutionary history to modern clinical outcomes. By utilizing positive selection as a biological prior, we can systematically prioritize functional variation and illuminate mechanistic links between past adaptation and modern disease susceptibility. As EHR-linked biobanks and long-read sequencing expand, this evolution-guided approach will become increasingly valuable for uncovering consequential genetic variation and advancing personalized medicine in diverse, understudied populations.

## Declarations

### Ethics approval and consent to

Not applicable.

### Consent for publication

Not applicable.

### Availability of data and materials

No new materials were generated for this study. This study used data from the All of Us Research Program’s Controlled Tier Dataset version 8, available to authorized users on the Researcher Workbench. Researchers must apply to the *All of Us* Research Program for access to Controlled Tier data. Exact participant counts for categories <20 cannot be shown as per the *All of Us* Data and Statistics Dissemination Policy and are instead shown as 20. Code and analysis used to generate figures are available on a public GitHub repository (https://github.com/soisa001/SelectionMapping). A demonstration workspace of the analysis is available on the *All of Us* Verily Workbench (https://workbench.verily.com/workspaces/aou-rw-a3e31f4d).

Adults 18 years and older who have the capacity to consent and reside in the USA or a US territory at present are eligible to participate in the *All of Us* Research Program. Informed consent for all participants is conducted in person or through an eConsent platform that includes primary consent, HIPAA Authorization for Research use of EHRs and other external health data, and Consent for Return of Genomic Results. This secondary data analysis research did not require IRB approval because it does not involve human participants. All data is now publicly available.

### Authors’ contributions

N.S. and P.H. conceived and designed the project. N.S., K.V.G, M.C.S., E.E.E., F.J.S., M.E.T., F.C., S.K.L, P.H., and the Long Reads Working Group selected samples and generated the long reads panel.

F.C. and S.K.L. performed imputation of structural variants into short read samples. N.S. and P.H. conducted analyses and wrote the manuscript with input from all authors.

## Data Availability

All data produced are available online at the All of Us Research Program's Controlled Tier Dataset version 8, available to authorized users on the Researcher Workbench.

## Acknowledgements

We gratefully acknowledge *All of Us* participants for their contributions, without whom this research would not have been possible. We also thank the National Institutes of Health’s *All of Us* Research Program for making available the participant data examined in this study. The content is solely the responsibility of the authors and does not necessarily represent the official views of the National Institutes of Health.

## Methods

### Demographic Inference

We modeled the demographic history of 1,025 unrelated African American samples (“AoU_AFA”) in the All of Us Research Program to provide a reasonable model for simulation and downstream analyses. Samples were sequenced using Illumina short read whole-genome sequencing to ∼30× coverage, and SNVs were jointly called [1]. We utilize the recommended site filters and mask difficult regions (difficult to call regions including centromeres, satellites, repetitive regions, segmental duplications >99% identity, X and Y chromosomes). To facilitate demographic inference, we included the 1KG high-coverage AFR and EUR individuals, serving as present-day samples from which we can jointly infer the AoU_AFA, AFR, and EUR demes [79]. First, we take AoU_AFA, AFR, and EUR samples and merge the intergenic SNVs (at least 100 kb away from refGene genes), and then remove related individuals using Hail pc_relate (k=10 PCs, kinship coefficient<0.1). This results in 1,025 AoU_AFA, 672 AFR, and 425 EUR samples. Then, we selected unadmixed AFR and EUR individuals by running unsupervised ADMIXTURE (v1.3) [80] using k=2 and 99,481 random intergenic SNPs, where only AFR and EUR individuals that had at least 98% ancestry assignment to their respective primary component were kept (Table S1). A high-quality chimpanzee genome was used as the ancestral outgroup for allele polarization [81]. In summary, we obtain a final set of 1KG 409 AFR, 418 EUR, and 1,025 AoU_AFA unrelated samples with 22,785,857 biallelic segregating SNVs across 481,020,725 bp of intergenic masked sequence. Due to computational limitations on inference, we project the joint site frequency spectrum down to 25 alleles in each population, resulting in 15,625 allele frequency bins. Iterative model fitting was then performed on the projected joint site frequency spectrum with dadi v2.2 (diffusion approximation for demographic inference) [25] on the unfolded SFS, where 100 perturbed parameter sets were fitted under each model to ensure that the parameters and likelihood converged. We correct for ancestral state misidentification with an explicit parameter in each model that flips a proportion of each half of the SFS. Log-likehood of models was computed using diffusion grid sizes [35,45,55]. We investigated various models including with or without migration, instantaneous or exponential growth, and admixture timings. Model selection was performed using likelihood ratio tests to obtain the best stable model. To estimate uncertainty, we generated 100 bootstrap projected replicates, and refitted the perturbed parameter set to obtain 95% confidence intervals. We use a generation time of 25 years and a mutation rate of 1.5×10^-8^ per base pair per generation to scale from genetic to physical units.

We project the 1025 AoU_AFA samples into geographic coordinates using genetic similarity to 1KG samples. We take 1KG samples of 409 AFR and 418 EUR individuals, composing four European sampling locations (GBR, IBS, TSI, FIN) and five African locations (GWD, MSL, YRI, ESN, LWK), and use these as reference locations. For each sample, we take 16 PCs from the shared PC space and compute the weight of each PC using their eigenvalues to weight the PCs by their genetic variance explained. Geographic coordinates (latitude and longitude) of the nearest 20 1KG samples by PC distance were then used to project the AoU_AFA sample location, weighting the contribution of each sample by their PC distance to the AoU_AFA sample such that closer samples by PC distance are given more weight. The projected latitude and longitude is then computed as the average of the PC-weighted latitude values and longitude values among the 20 samples and then plotted along with the 1KG locations.

### Selection scans and coalescent simulation

We next scanned the genome for recent positive selection in the AoU_AFA samples using the integrated haplotype score (iHS) statistic [35]. We use the 1,025 AoU_AFA panel, generated from long reads in the marker paper, to compute iHS using selscan v2.1 [82] and the hg38 genetic map [83], and then normalized scores genome-wide into 100 allele frequency bins using recommended parameters [84]. The genome was partitioned into non-overlapping 20 kb windows, and the maximum |iHS| value in each window was recorded.

To establish a null distribution under neutrality, we performed 1,000 coalescent simulations using msprime v1.3.3 [85]. These simulations incorporated the GRCh38 recombination map and the inferred demographic model from dadi, with parameter sets for effective population sizes, divergence time, migration rates, and admixture fraction independently drawn from the multivariate normal distribution of 100 bootstrap replicates. Each simulated dataset was processed in the same manner as the empirical data to produce a distribution of maximum |iHS| values per window. For each empirical window, a simulation-based p-value was calculated as the proportion of simulated windows with a maximum |iHS| value greater than or equal to the observed value. Over 122,828 20 kb windows across the genome, we find 2,345 significant windows (p<=0.01). We computed EHH (extended haplotype heterozygosity) with rEHH [86] using haplotypes carrying the reference or alternative allele at candidate iHS leads. For SVs of interest, we plot the locus using svbyeye [87] by querying for samples that carry the SV and aligning the region flanking to SV in hg38 to contigs in the sample assemblies.

We use the UCSC-provided hg38 RefGene track, updated Jan 10, 2020, and use all annotated genes [88]. We computed a Fisher exact test for enrichment, finding 1.08-fold genic enrichment, 1.15-fold exonic enrichment, and 1.08-fold intronic enrichment. We conduct permutation testing for gene enrichment against all 122,828 windows, where the permutation p-value is the number of times a permutation of 2,345 windows randomly drawn yields a greater proportion of genic (or exonic or intronic) windows. We merge consecutive windows with sim p ≤ 0.01 (n=1616). We focus on exonic windows as our primary candidate set of selected loci for downstream analysis (n=963). We conduct permutation testing by testing for enrichment in gene constraint (window contains gene with pLI > 0.9) [77], and ENCODE4 cCRE [38]. For gene constraint, we utilize the mean pLI on selected windows, draw 963 windows randomly from the 122,828 windows that contained a gene with annotated pLI, and compute the two-tailed p-value as the proportion of draws that had windows with mean pLI in window more extreme than observed. For cCRE enrichment, we draw 963 windows randomly from all 122,828 windows and compute the p value as the proportion of draws where the fraction of windows with the IHS lead overlapping a cCRE is more extreme than observed.

We conduct Panther GO-Slim enrichment analysis (molecular function, cellular component, biological process) on the exonic gene list of candidate loci against the Homo sapiens database (Panther v19.0, 20,580 genes) [89]. For Panther-GO-Slim molecular function at FDR=0.05, we find significant enrichment for catalytic activity (1.26-fold), transferase activity (1.4-fold), but depletion for transmembrane signaling receptor activity (0.54-fold), and cis-regulatory region sequence-specific DNA binding (0.52-fold) (Table S5). Cellular component and biological process results are in Table S5.

### Colocalization to MAGE catalog and GWAS catalog

We conduct colocalization analysis on our candidate set using the multi-ancestry database of gene expression fine-mapped results (MAGE sQTL and eQTL v1.0) [40] and the GWAS catalog (v1.0.2) [41]. For each candidate selected loci, we take all SNVs in linkage to the lead IHS variant (r2 > 0.5, +- 100 kb, 1,025 panel). We use variants in any sQTL or eQTL credible set for MAGE, and variants below the genome-wide significance threshold p=5e×10^-8^ for GWAS. 84 loci have a linked eQTL, 46 loci have a linked sQTL, and 241 loci have a linked GWAS trait. 52 loci have a linked eQTL/sQTL and a GWAS hit, and 12 loci have all three. Four loci are linked to an SV and a eQTL, sQTL, or GWAS hit.

We conduct permutation testing by testing the enrichment of the GWAS, eQTL and sQTL signal in the candidate set compared to the overall windows. We draw 963 windows randomly from the 122,828 windows, compute all linked variants to the IHS lead of the drawn windows, and compute the number of windows with colocalized hits (GWAS, eQTL, or sQTL). The p-value is the number of times that a draw had a larger proportion of windows with a colocalized hit than the empirical observed value.

### Phenome-wide association tests

We conducted PheWAS on the candidate exonic loci. We used PheTK (the Phenotype Toolkit) as the primary package for PheWAS analyses [90]. Phenotypes were defined using phecodeX v1.2 [91]. Individuals required at least two phecode counts to be counted as a case. Only phecodes with at least 200 cases and 200 controls were included in the analysis. Sex-specific phenotypes were tested only on their respective groups. The main discovery cohort consists of 10,000 self-identified African Americans, for which 8,771 samples were unrelated (kinship coefficient<0.1), had covariates and were a case or control for at least one phecode. We conduct replication on 28,552 (kinship coefficient<0.1) unrelated AFR-like samples, provided by All of Us (CDR v8 predicted ancestry cohorts which were generated from a random forest classifier trained on principal components with 1KG and Human Genome Diversity Project samples).

The PheWAS model is a logistic regression of the form:

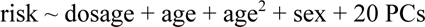

where dosage was coded as 0, 1, or 2 copies of the alternative allele.

PCs were computed from the pooled discovery and replication cohort in Hail (hail_normalized_pca) using the All of Us global PCA high-quality SNVs (Genomic Research Data Quality Report) [92]. In the case that the lead variant is an SV, we test the best linked SNV in lieu of the SV (r2 > 0.5, +- 100 kb).

Regression models were computed with Newton–Raphson optimization and a maximum of 100 iterations (statsmodels v0.14.0) [93]. Variants with minor allele counts <20 and unconverged regressions were dropped from further analysis. In total, 428,857 tests were performed in the AFR cohort. We applied a lenient threshold of p<1e-3 for candidate hits, obtaining 446 nominal associations. For nominal associations, fine-mapping was performed with SuSIE v0.16 [94] for each outlier region ±500 kb, with the same covariates as in the PheWAS, and with minor allele count>20.We retest each candidate phecode in the replication cohort of 28,552 samples, followed by inverse variance weighted meta-analysis, with 17 hits significant at the meta-analysis level after Bonferroni correction on 446 tests (p=1.12×10^-4^). Beta (β), odds ratios and 95% confidence intervals from the standard error were derived from the regression models for the genotype dosage.

### Local Ancestry

Local ancestry was performed using flare v0.6 [28] with default parameters, including minor allele count=50 and minimum allele count=0.005 in the reference panel. We use the 1,025 AoU_AFA [23] as the query and 1KG high coverage release [79, 95] of the phased variants as the reference panel, where the same AFR and EUR samples as in the demographic inference were used to construct the panel in bref3 format. We utilize the hg38 genetic map [96]. Each chromosome was run separately to generate the phased marker-level local ancestry per haplotype for each sample in AoU_AFA. At each marker, we compute the EUR and AFR ancestry fraction over all samples to obtain local ancestry outlier regions, defined as regions where the EUR marker local ancestry across all samples deviates significantly from the mean (±4.42 SD) [29]. We merge consecutive markers within 200 kb and use the first and last marker as the breakpoints. We remove regions within 2 Mb of telomeres or overlapping the centromere, obtaining 228 outlier regions. Then, we adjust the PheWAS method to explore regional associations using predicted EUR dosage by fitting local ancestry prediction models on the AOU_AFA samples and then applying these to the discovery and replication cohort. The 1,025 AoU_AFA samples were split 80:20 for training (n=820) and test (n=205) data. For each region, we fit a partial least squares (PLSR) model with up to 20 components to predict basepair-weighted EUR ancestry dosage per sample from their marker genotypes. Each sample may have a EUR dosage from 0 to 2, corresponding to no EUR ancestry called over a region to the entire region called EUR over both haplotypes. Partial dosages are allowed. We test performance over the holdout samples, keeping models with test r^2^ > 0.5 and p below 0.05 compared to the null model (mean-prediction without SNV information). We run PheWAS for 447 phecodes with case and control counts above 200 for each PLSR model, under the following PheWAS model:

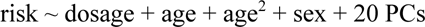

where dosage was coded as the predicted EUR dosage, such that risk effects are with respect to increasing EUR dosage. The PheWAS was run separately for the discovery and replication cohorts. We use p<1×10^-3^ for nominal significance, and conduct inverse variance weighted meta analysis on nominal associations. For discovery associations, fine-mapping was performed with SuSIE v0.16 [94] for each outlier region ±100 kb, with the same covariates as in the PheWAS and the local ancestry markers.

