## Supplementary Figures for "Evolution-guided association mapping using long-read genomes from a self-identified Black or African American cohort in *All of Us*"

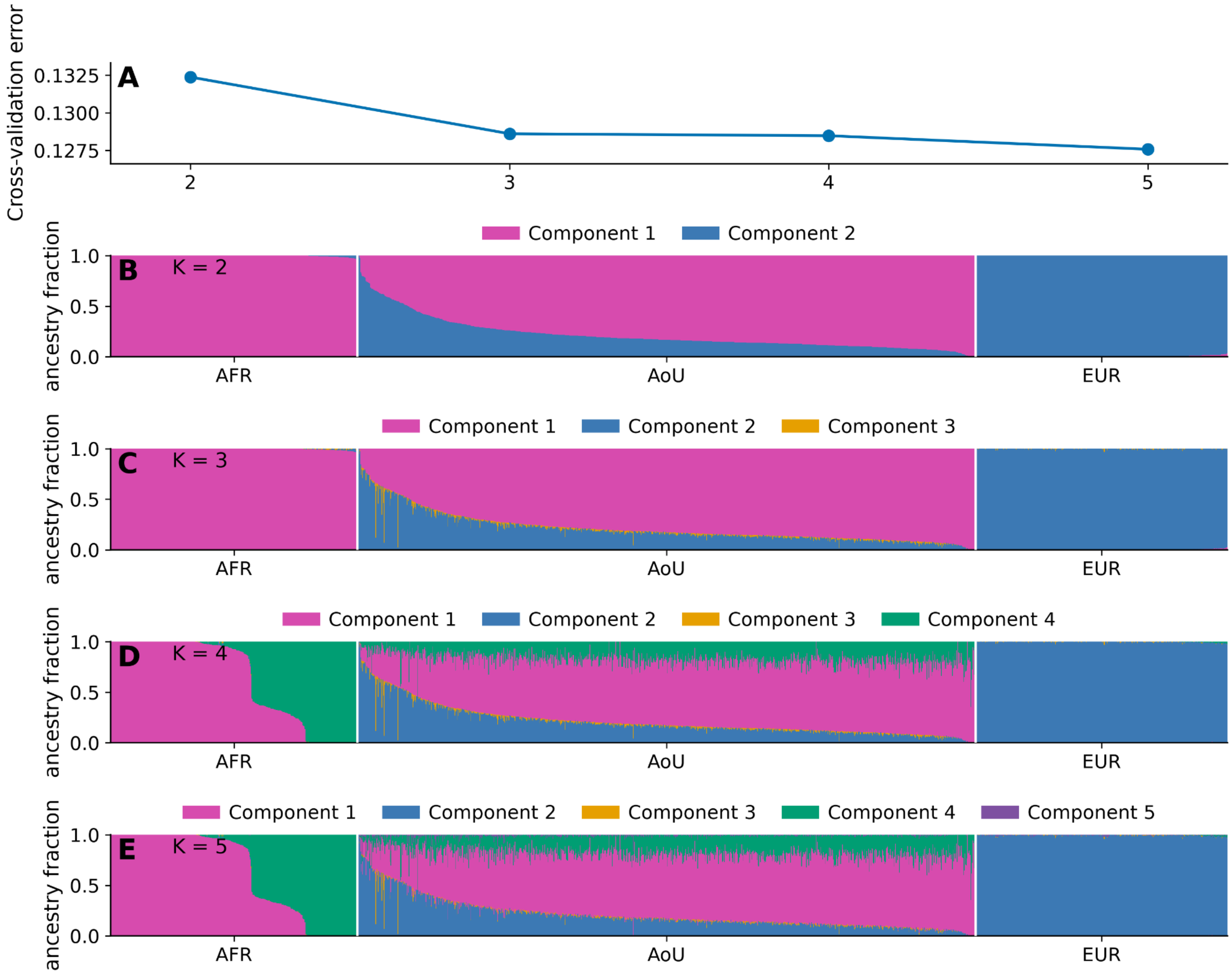


**F**

| Components $F_{ST}$ | K = 2 | K = 3 | K = 4 | K = 5 |
|---|---|---|---|---|
| 1 - 2 | 0.113 | 0.120 | 0.122 | 0.122 |
| 1 - 3 | | 0.144 | 0.146 | 0.146 |
| 1 - 4 | | | 0.009 | 0.009 |
| 1 - 5 | | | | 0.165 |
| 2 - 3 | | 0.097 | 0.097 | 0.098 |
| 2 - 4 | | | 0.122 | 0.122 |
| 2 - 5 | | | | 0.106 |
| 3 - 4 | | | 0.147 | 0.147 |
| 3 - 5 | | | | 0.090 |
| 4 - 5 | | | | 0.166 |

**Figure S1. ADMIXTURE analysis of AOU and 1,000 Genomes AFR and EUR samples.** A) 1,025 AOU samples, 409 AFR, and 418 EUR samples were used in unsupervised ADMIXTURE with k=2-5 components. CV validation error is shown (lowest CV at k=5). B) ADMIXTURE components of AFR, AOU, and EUR samples at k=2. C) ADMIXTURE components of AFR, AOU, and EUR samples at k=3. D) ADMIXTURE components of AFR, AOU, and EUR samples at k=4. E) ADMIXTURE components of AFR, AOU, and EUR samples at k=5. F) $F_{ST}$ of components at each k.

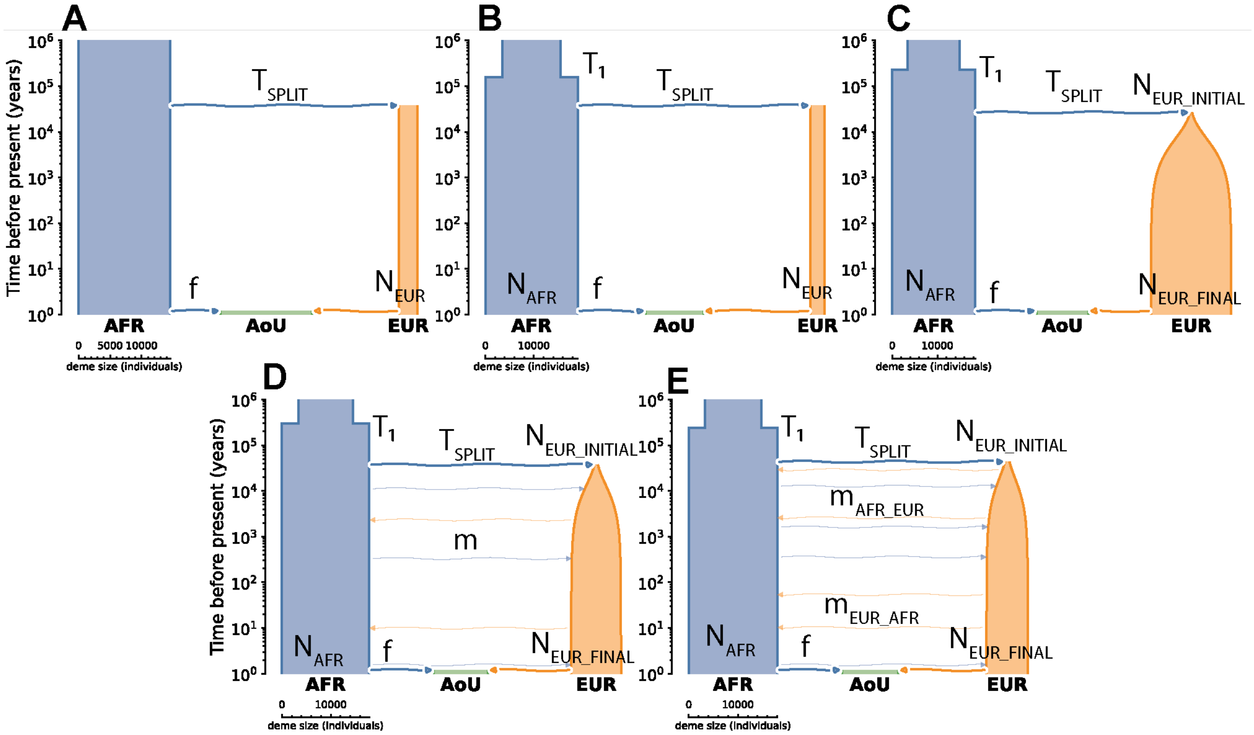


**Figure S2. Joint demographic inference of 1,025 AOU samples, 409 AFR, and 418 EUR samples.** Iteratively more complex demographic models were fit to the joint site frequency spectrum, with log-likelihoods (LL) of (A) −77,790.07, (B) −55,965.32, (C) −42,776.33. The two final models (D) and (E) were compared with log likelihood ratio test (panel D: one parameter for migration rate between AFR and EUR demes, LL= −35,557.40, and panel E: two parameters for migration rate between AFR and EUR demes, LL=−35,290.57), supporting the two parameter migration rate model (Godambe-adjusted *D* statistic = 20.01, chi-square p = $7.69 \times 10^{-6}$ at df=1). Best fit parameter values for each model in Table S2.

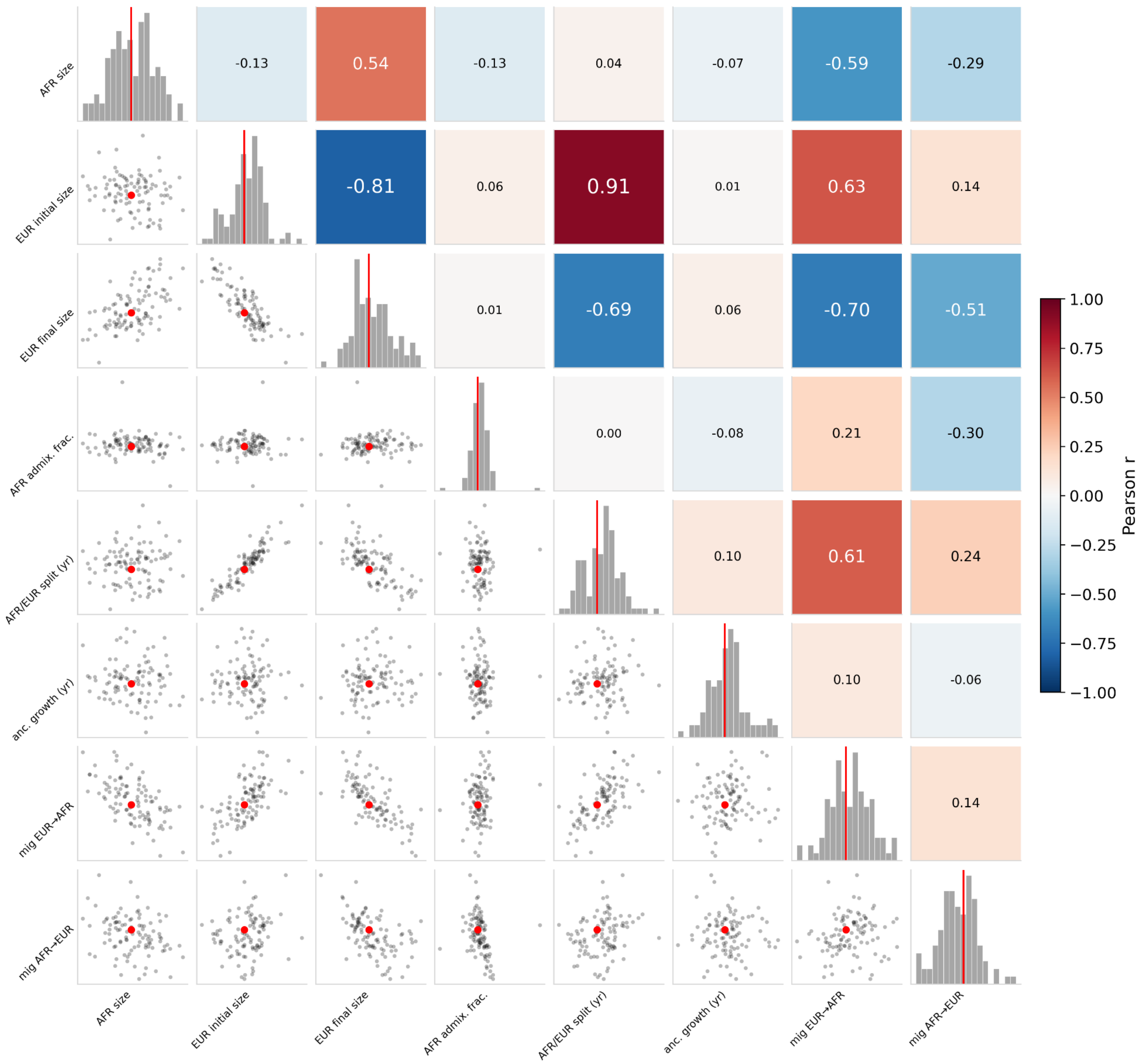


**Figure S3. Bootstrap correlogram of pairwise relationships between dadi inferred parameters for the best fit model.** Bootstrapped estimates are in black and best fit parameter value in red. Parameters, from left to right: $N_A$, ancestral population size; $N_{EUR\_INITIAL}$, European initial $N_e$; $N_{EUR\_FINAL}$, European final $N_e$; $f_{AFR}$, fraction of African genetic component at admixture event; $T_{SPLIT}$, EUR–AFR split time; $T_{GROWTH}$, ancestral expansion time; $m_{EUR\rightarrow AFR}$, migration rate EUR→AFR (per generation); $m_{AFR\rightarrow EUR}$, migration rate AFR→EUR (per generation). 95% confidence intervals for parameters were computed from bootstraps (Table S2).

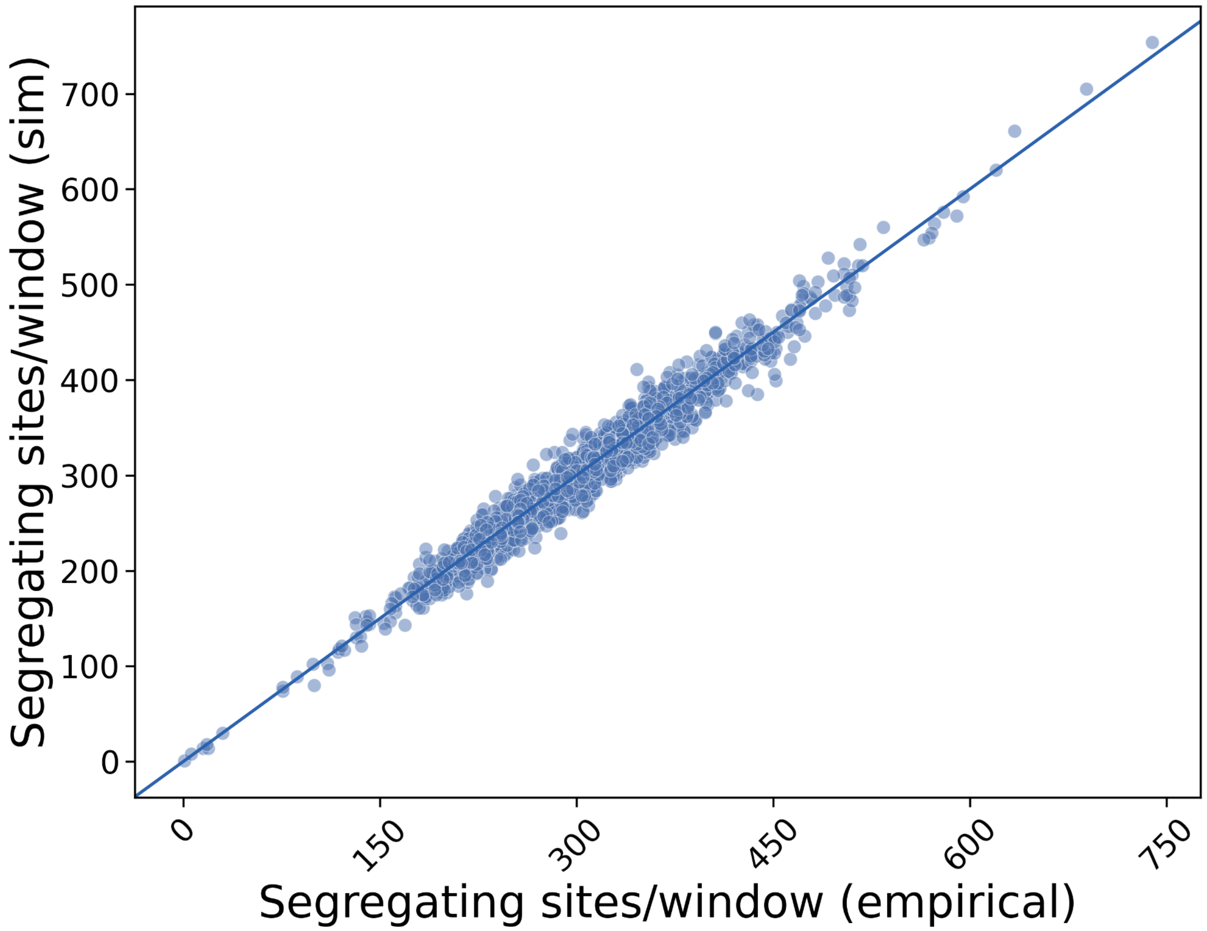


**Figure S4. Mutation rate matching between observed windows and simulated 20 kbp windows for one representative simulation on chr22.** Empirical counts of segregating sites (theta) are generated on a per-window basis, followed by mutations simulated in msprime, and then dropping sites proportionally to match the observed theta (Pearson r=0.98, $r^2$=0.96). Sites in masked bases were removed from empirical and simulated counts and not included in analysis.

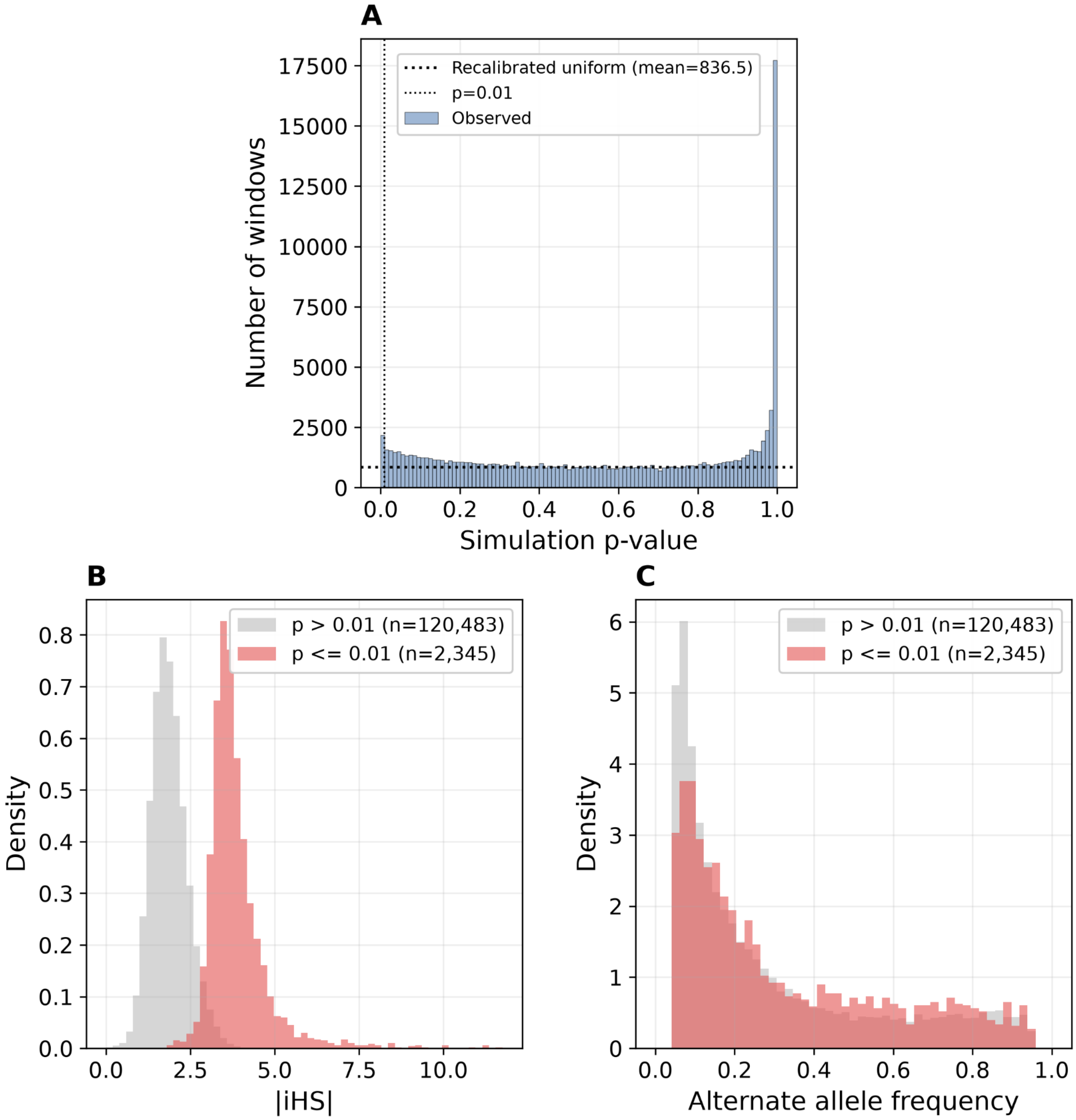


**Figure S5. Distribution of simulated p-values with |iHS| and allele frequency spectrum for windows sim p≤0.01.** Under the uniform null indicated where the null is taken as the first plateau where two consecutive bins of size 0.05 change by no more than 2.5%, FDR is 35.7% at $p<0.01$ (836.5 expected false positives over 2,345 hits). Actual FDR is likely between 20-30% taking into consideration linkage between windows (2,345 20 kbp windows collapse to 1,616 merged consecutive windows). **(B)** |iHS| distribution of the windows with sim $p <= 0.1$ and sim $p > 0.01$. **(C)** Alternate allele frequency spectrum of the windows with sim $p <= 0.1$ and sim $p > 0.01$. Candidate windows show a significantly higher alternate allele frequency (median AF=22.78%) than the non-candidate windows (median AF=17.48%, one-sided Mann-Whitney U test, $p=7.22 \times 10^{-27}$, rank-biserial r=0.128)

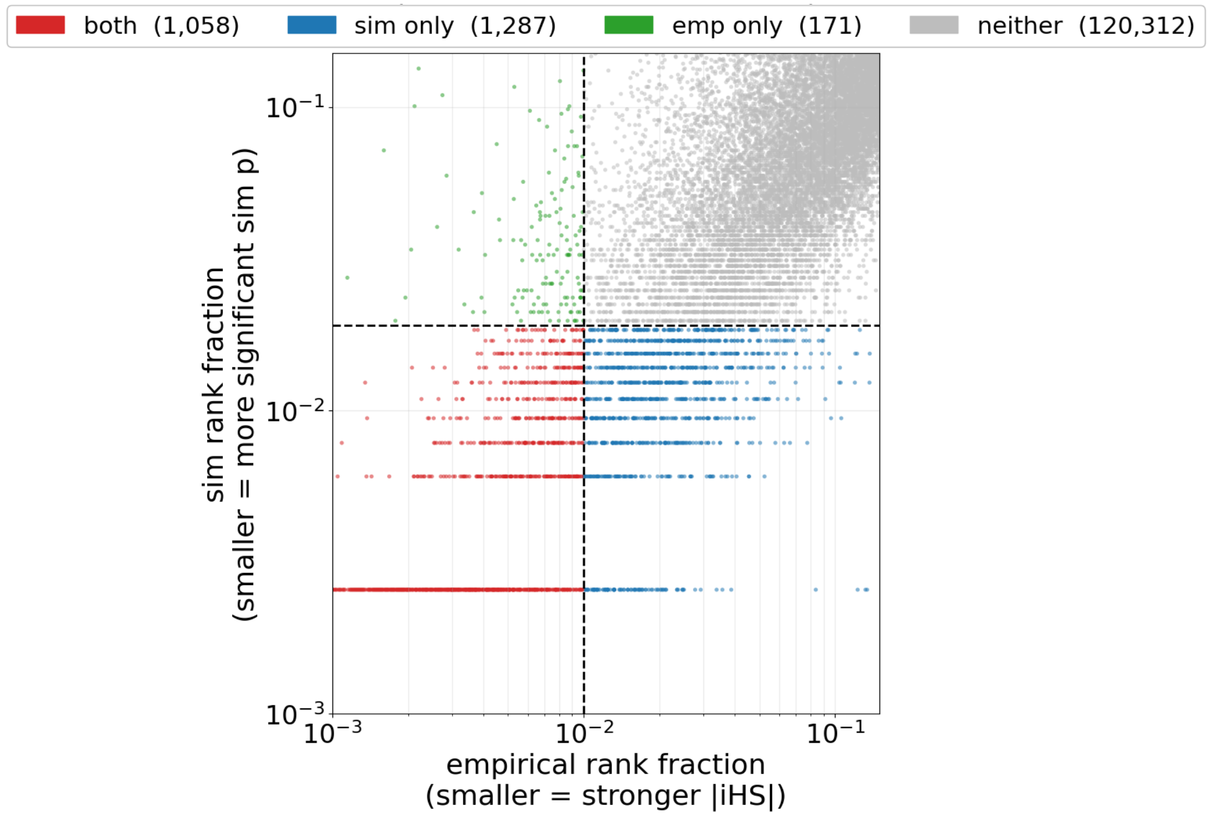


**Figure S6. Empirical rank vs simulation rank of selected windows.** Each quadrant shows windows that are above or below 0.01 by empirical rank or sim rank. Quadrant I shows windows not significant by either method. Quadrant III shows windows considered significant by both methods. The simulation recovers 1287 windows that are not in the top 0.01 by |IHS| empirical rank (Quadrant IV), and discards 171 false positives that are in the top 0.01 by empirical rank but are not significant by simulation (Quadrant II).

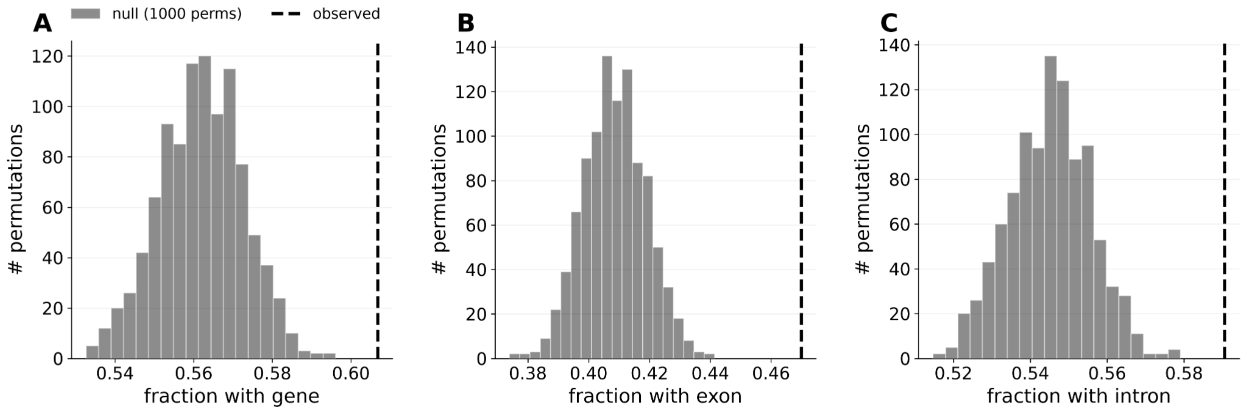


**Figure S7. Permutation test, 1000 permutations, on A) gene, B) exon, and C) intron enrichment, for fraction of windows with element of interest in randomly selected windows compared to empirical candidate set.** Enrichment is 1.08x for genes, 1.15x for exons, and 1.08x for introns (permutation p<0.001 for each).

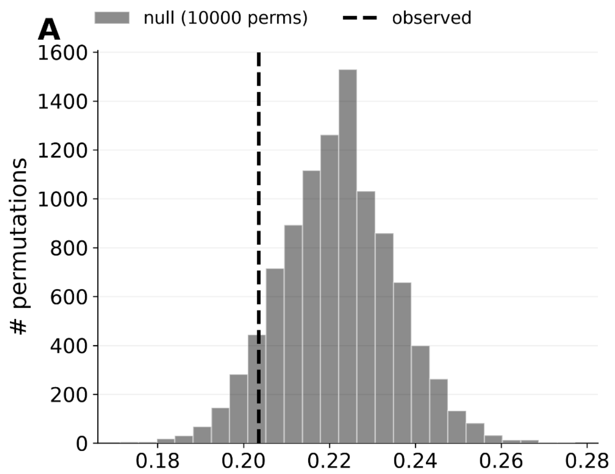


**Figure S8. Permutation test, 1000 permutations, on A) cCRE overlap and B) mean pLI, randomly selected windows vs candidate windows.** For mean pLI, the null includes only draws from windows that contain a gene with an annotated pLI. Enrichment is 0.92x for cCRE overlap (20.46% of the genic loci in candidate set, p=0.17) and 0.96x for mean pLI (mean probability of loss intolerance [pLI] in candidate set=0.33, p=0.30).

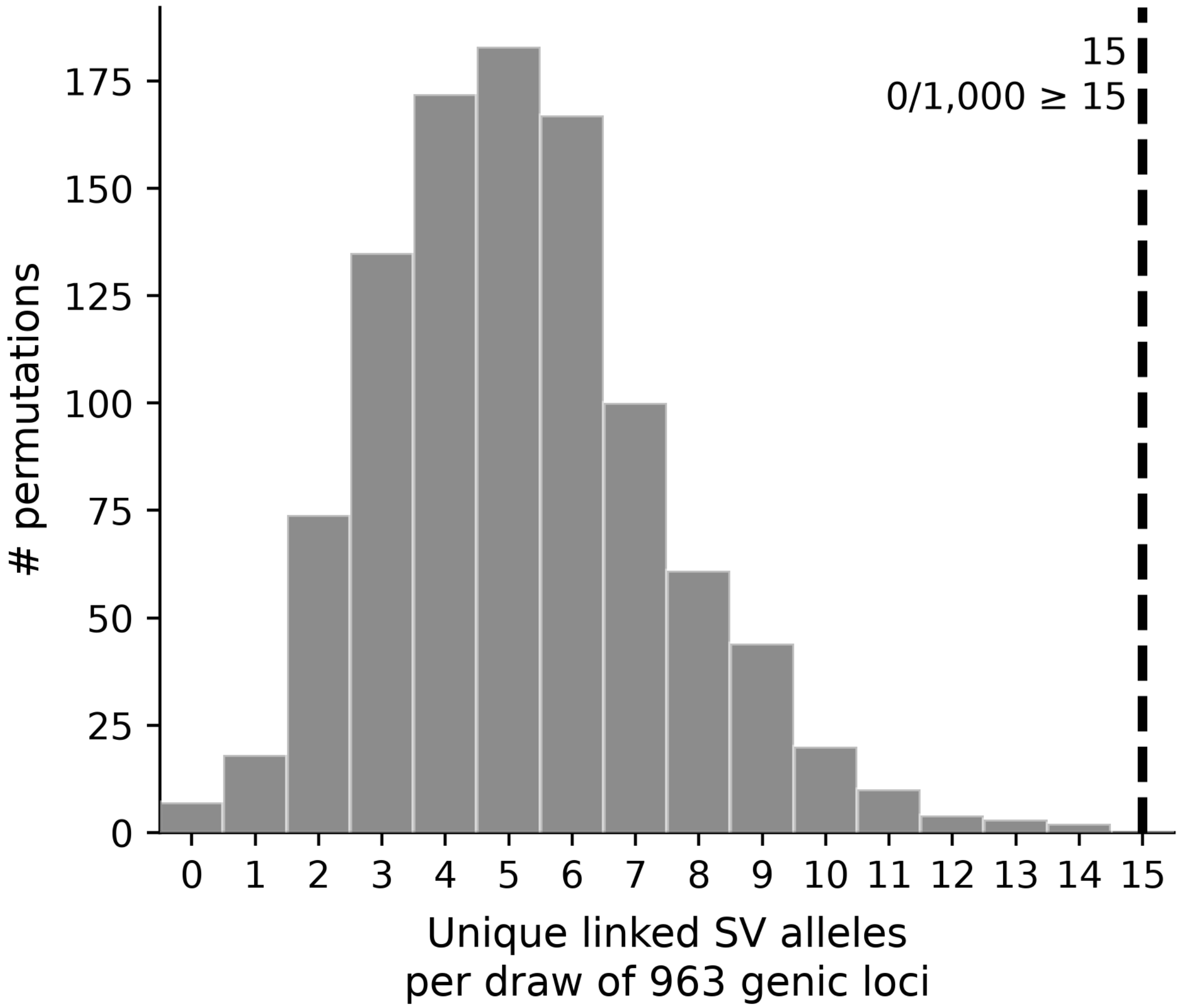


**Figure S9. Permutation test, 1000 permutations, on SV enrichment, randomly selected windows vs candidate windows.** 963 genic windows are randomly drawn, linkage to SVs computed to the lead iHS variant, and total number of unique SVs (SV leads or linked SVs) is counted ($r^2$>0.5 within 100 kb). The null includes only draws from genic windows not under selection (p>0.001). Enrichment is 2.89-fold enrichment, with 15 unique SVs observed in the genic candidate set vs mean 5.19 SVs observed in permutations.

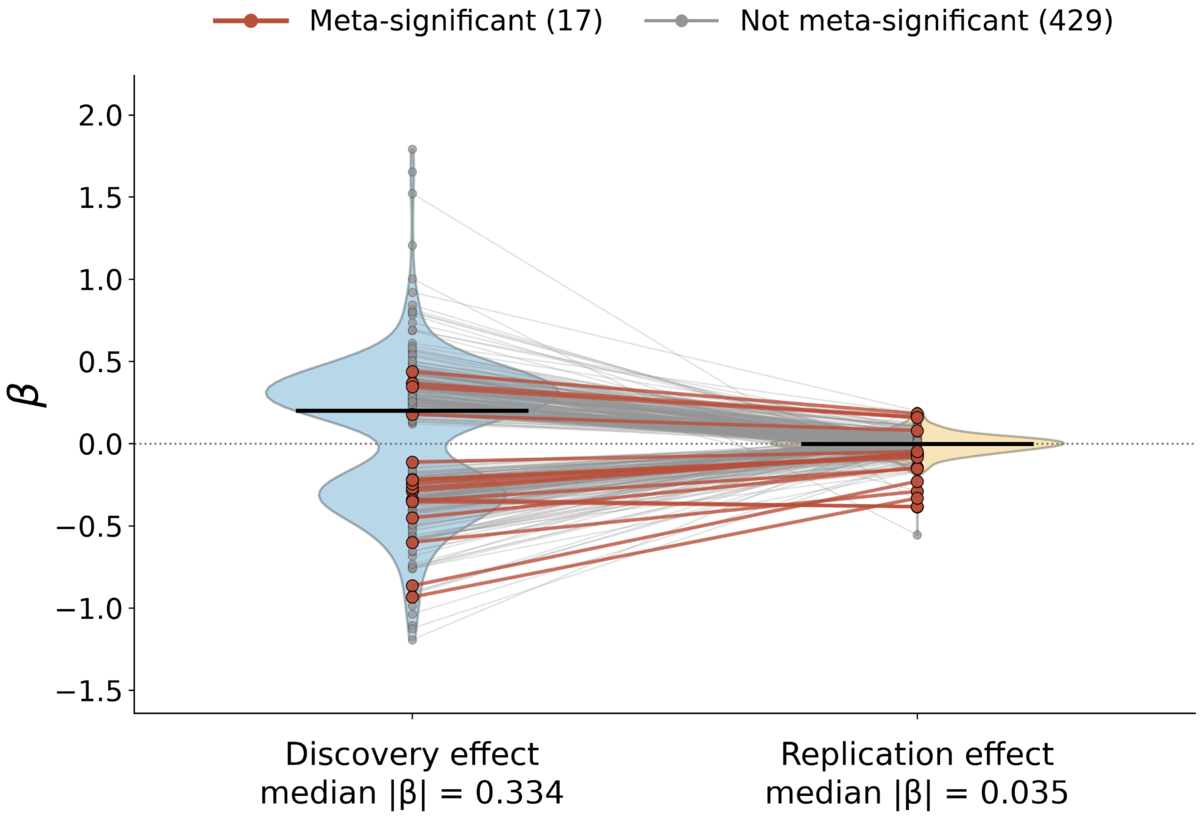


**Figure S10. Discovery effect size vs replication effect size**. Out of 446 phecodes that were significant in the discovery stage (discovery $p < 1x10^{-3}$), 17 phecodes were significant after meta-analysis with the replication cohort. Nine phecodes significant in discovery were not testable in replication due to replication minor allele count <20 or case and control count <200. Effect sizes are attenuated approximately 10-fold in replication compared to discovery.

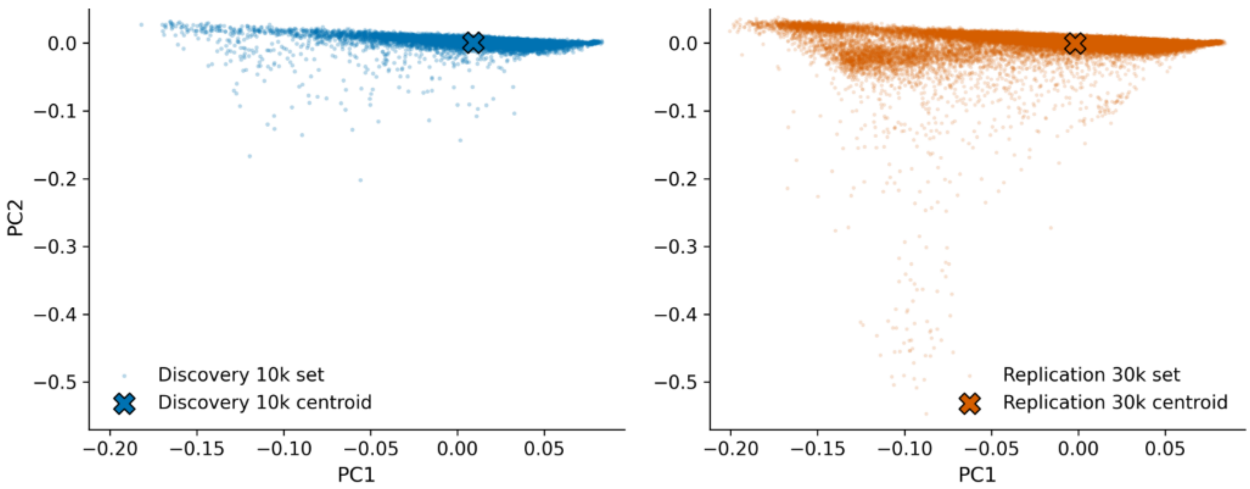


**Figure S11. Plot of PC1 and PC2 for the A) discovery cohort and the B) replication cohort.** 20 PCs were jointly computed using samples in both cohorts from short read data, from the subset of high-quality sites provided by All of Us for use in PCA.

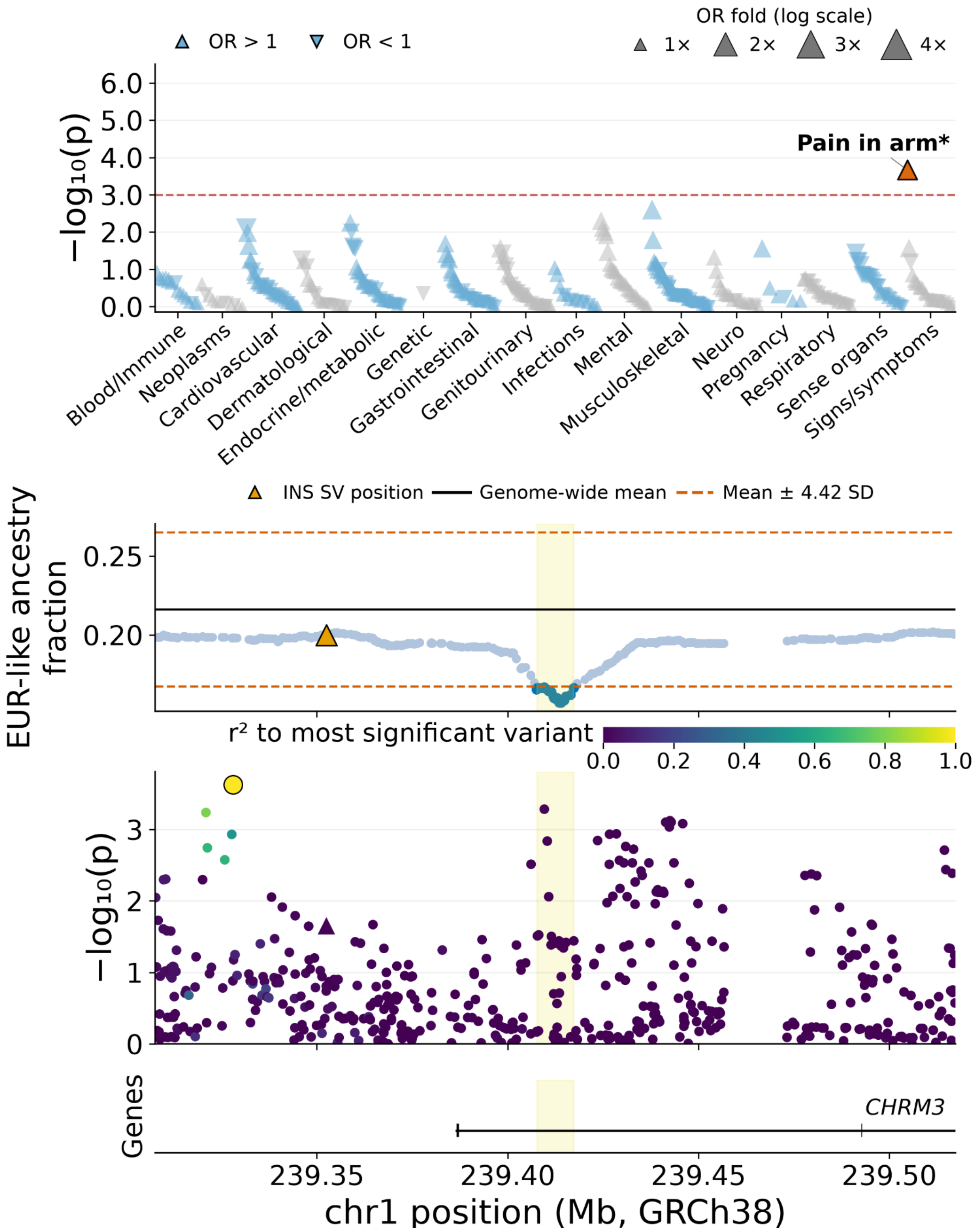


**Figure S12. Fine-mapping of the PheWAS association for phecode "pain in arm" (SS_809.31) at admixture outlier region chr1:239,407,597–239,417,310.** Top panel: PheWAS Manhattan plot across 446 phecodes in 16 categories. Upward and downward triangles represent increased and decreased odds for individual phecode outcomes, respectively. Middle panel: EUR-like ancestry fraction across markers. The outlier region is indicated in yellow. Bottom panel: Variant association signals across the locus. Variant colors indicate linkage disequilibrium ($r^2$) with the lead test variant. Gene locations are indicated.

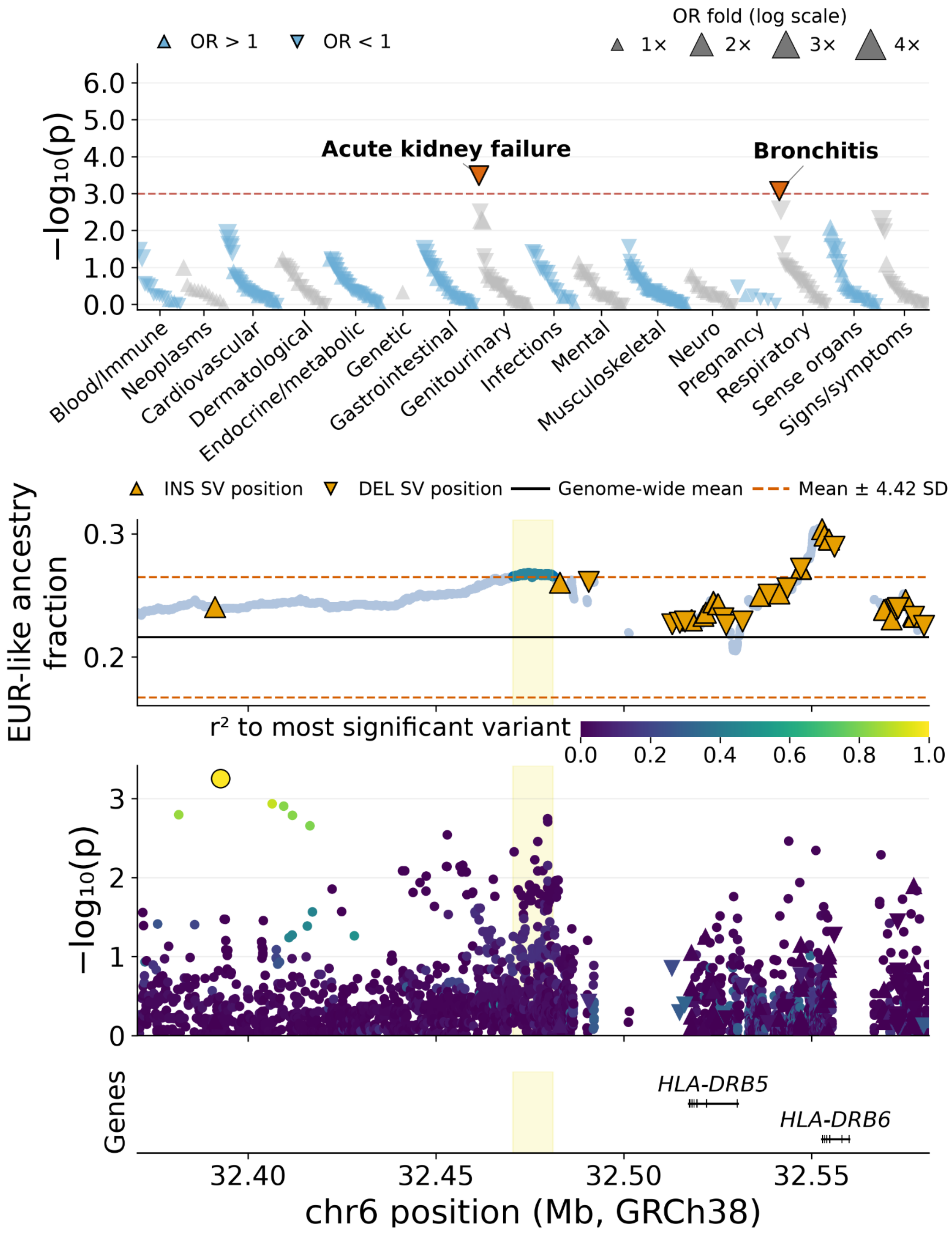


**Figure S13. Fine-mapping of the PheWAS association for phecode "bronchitis" (RE_469) at admixture outlier region chr6:32,470,563–32,481,211.** Top panel: PheWAS Manhattan plot across 446 phecodes in 16 categories. Upward and downward triangles represent increased and decreased odds for individual phecode outcomes, respectively. Middle panel: EUR-like ancestry fraction across markers. The outlier region is indicated in yellow. Bottom panel: Variant association signals across the locus. Variant colors indicate linkage disequilibrium ($r^2$) with the lead test variant. Gene locations are indicated.

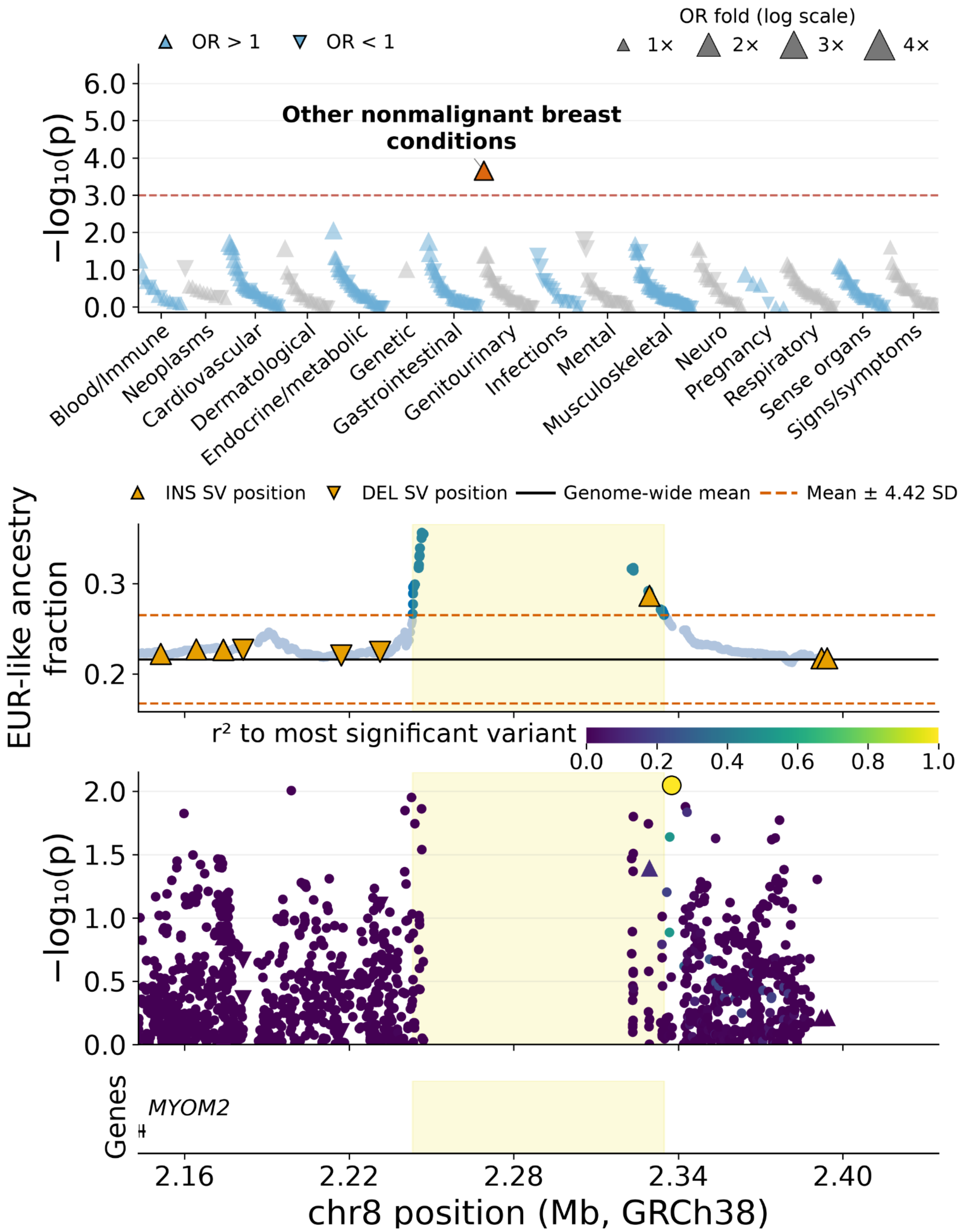


**Figure S14. Fine-mapping of the PheWAS association for phecode "other nonmalignant breast conditions" (GU_612) at admixture outlier region chr8:2,243,194–2,334,803.** Top panel: PheWAS Manhattan plot across 446 phecodes in 16 categories. Upward and downward triangles represent increased and decreased odds for individual phecode outcomes, respectively. Middle panel: EUR-like ancestry fraction across markers. The outlier region is indicated in yellow. Bottom panel: Variant association signals across the locus. Variant colors indicate linkage disequilibrium ($r^2$) with the lead test variant. Gene locations are indicated.

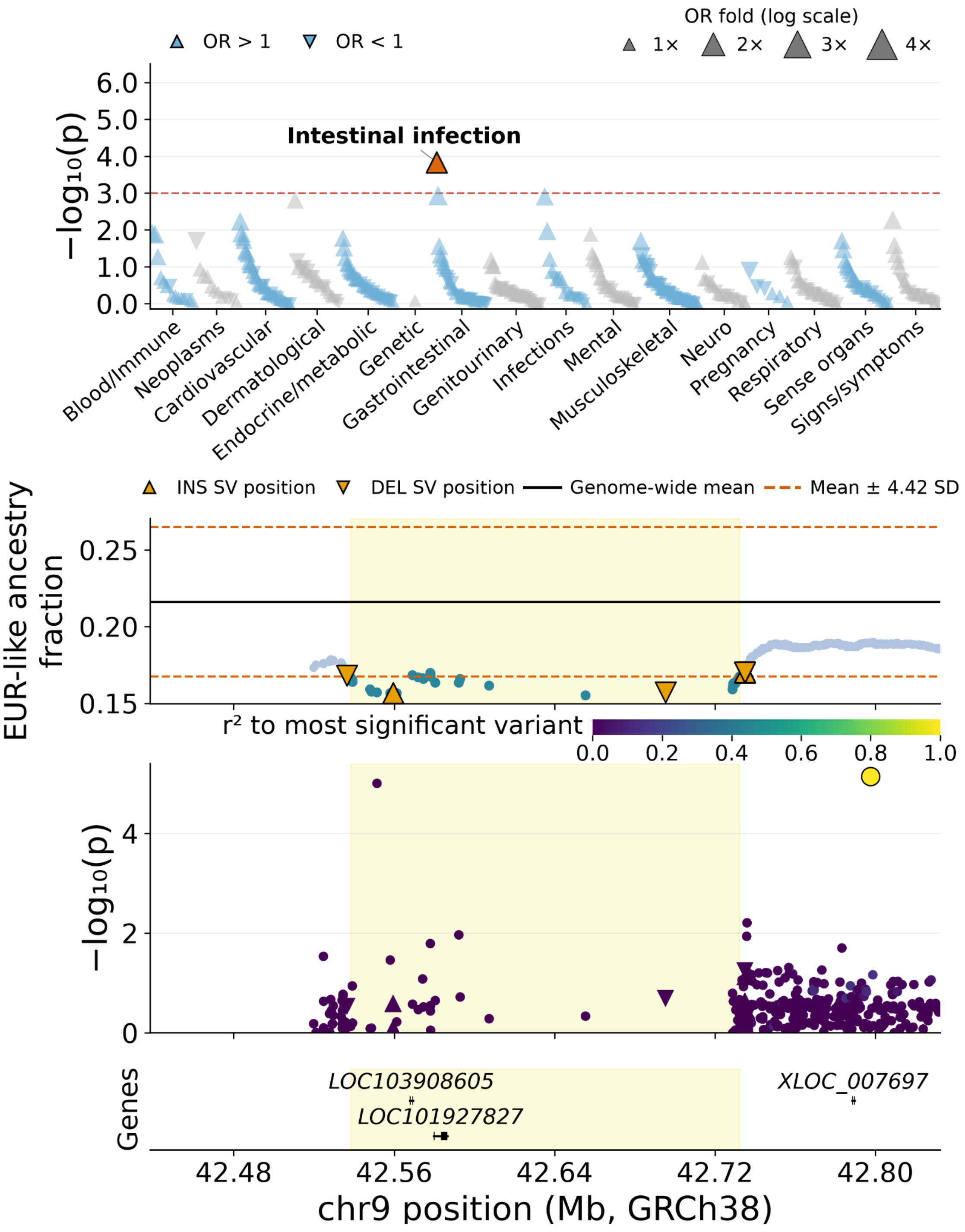


**Figure S15. Fine-mapping of the PheWAS association for phecode "intestinal infection" (GI_526) at admixture outlier region chr9:42,538,337–42,732,343.** Top panel: PheWAS Manhattan plot across 446 phecodes in 16 categories. Upward and downward triangles represent increased and decreased odds for individual phecode outcomes, respectively. Middle panel: EUR-like ancestry fraction across markers. The outlier region is indicated in yellow. Bottom panel: Variant association signals across the locus. Variant colors indicate linkage disequilibrium ($r^2$) with the lead test variant. Gene locations are indicated.

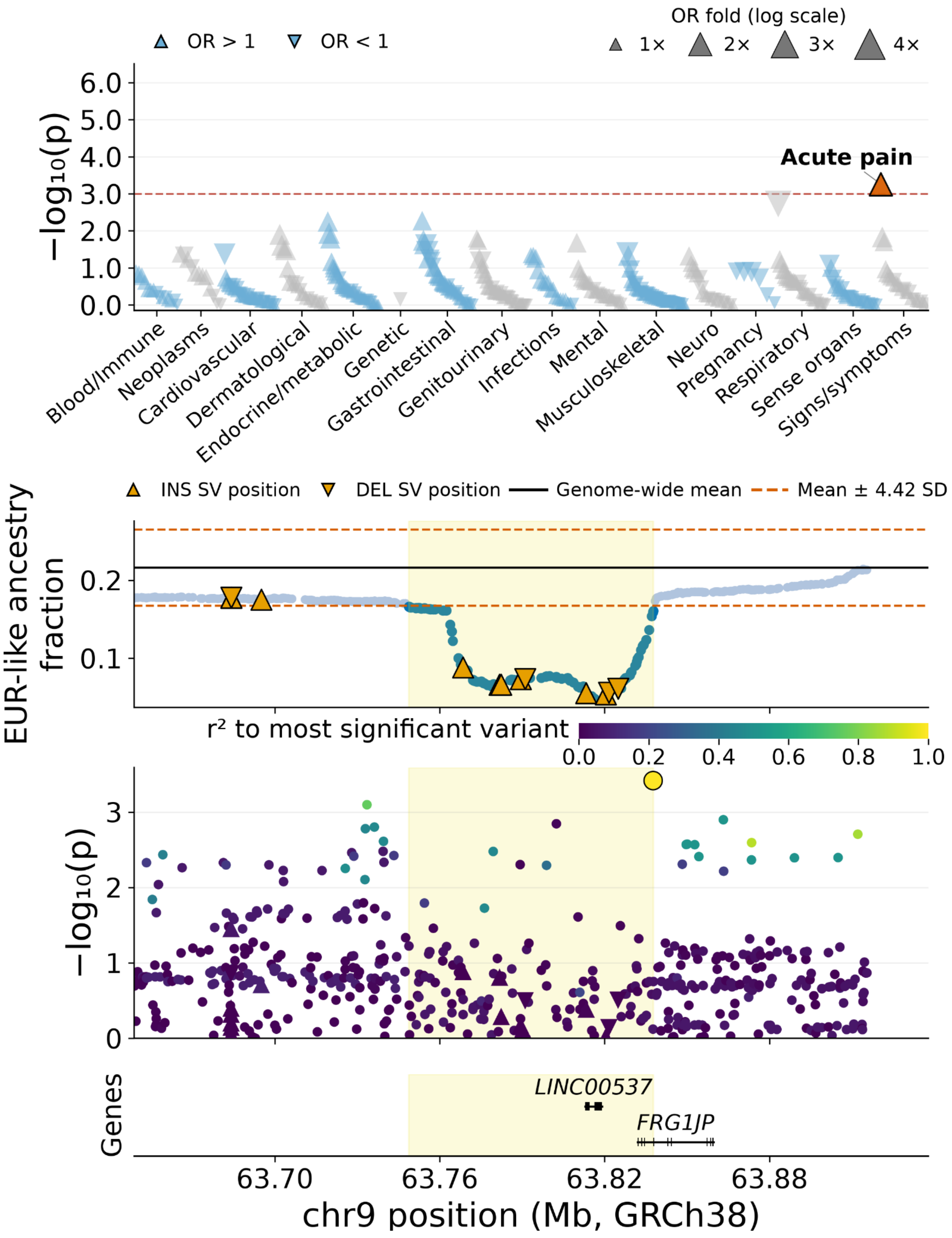


**Figure S16. Fine-mapping of the PheWAS association for phecode "acute pain" (SS_809.1) at admixture outlier region chr9:63,748,785–63,837,789.** Top panel: PheWAS Manhattan plot across 446 phecodes in 16 categories. Upward and downward triangles represent increased and decreased odds for individual phecode outcomes, respectively. Middle panel: EUR-like ancestry fraction across markers. The outlier region is indicated in yellow. Bottom panel: Variant association signals across the locus. Variant colors indicate linkage disequilibrium ($r^2$) with the lead test variant. Gene locations are indicated.

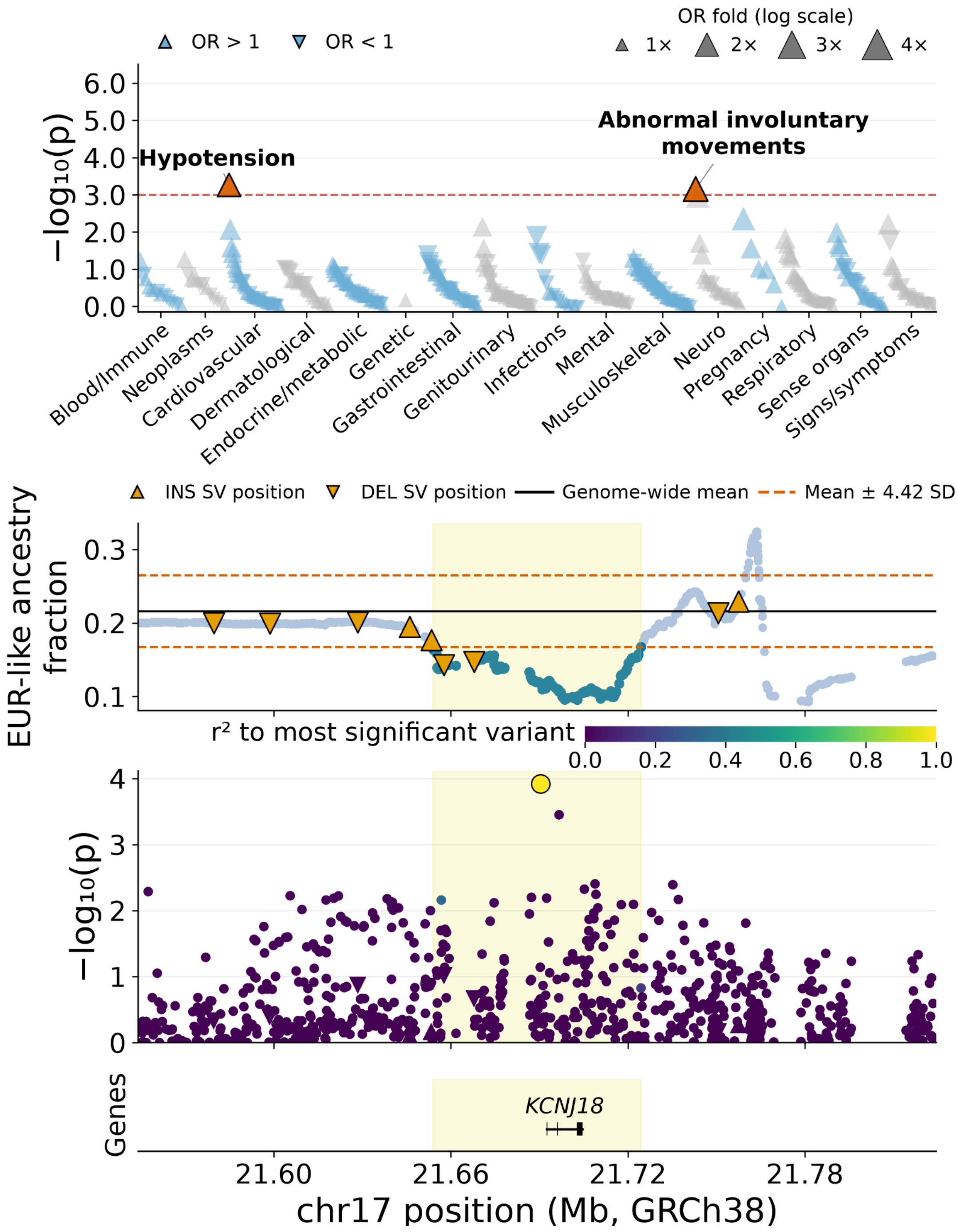


**Figure S17. Fine-mapping of the PheWAS association for phecode "abnormal involuntary movements" (NS_324.3) at admixture outlier region chr17:21,654,025–21,724,463.** Top panel: PheWAS Manhattan plot across 446 phecodes in 16 categories. Upward and downward triangles represent increased and decreased odds for individual phecode outcomes, respectively. Middle panel: EUR-like ancestry fraction across markers. The outlier region is indicated in yellow. Bottom panel: Variant association signals across the locus. Variant colors indicate linkage disequilibrium ($r^2$) with the lead test variant. Gene locations are indicated.

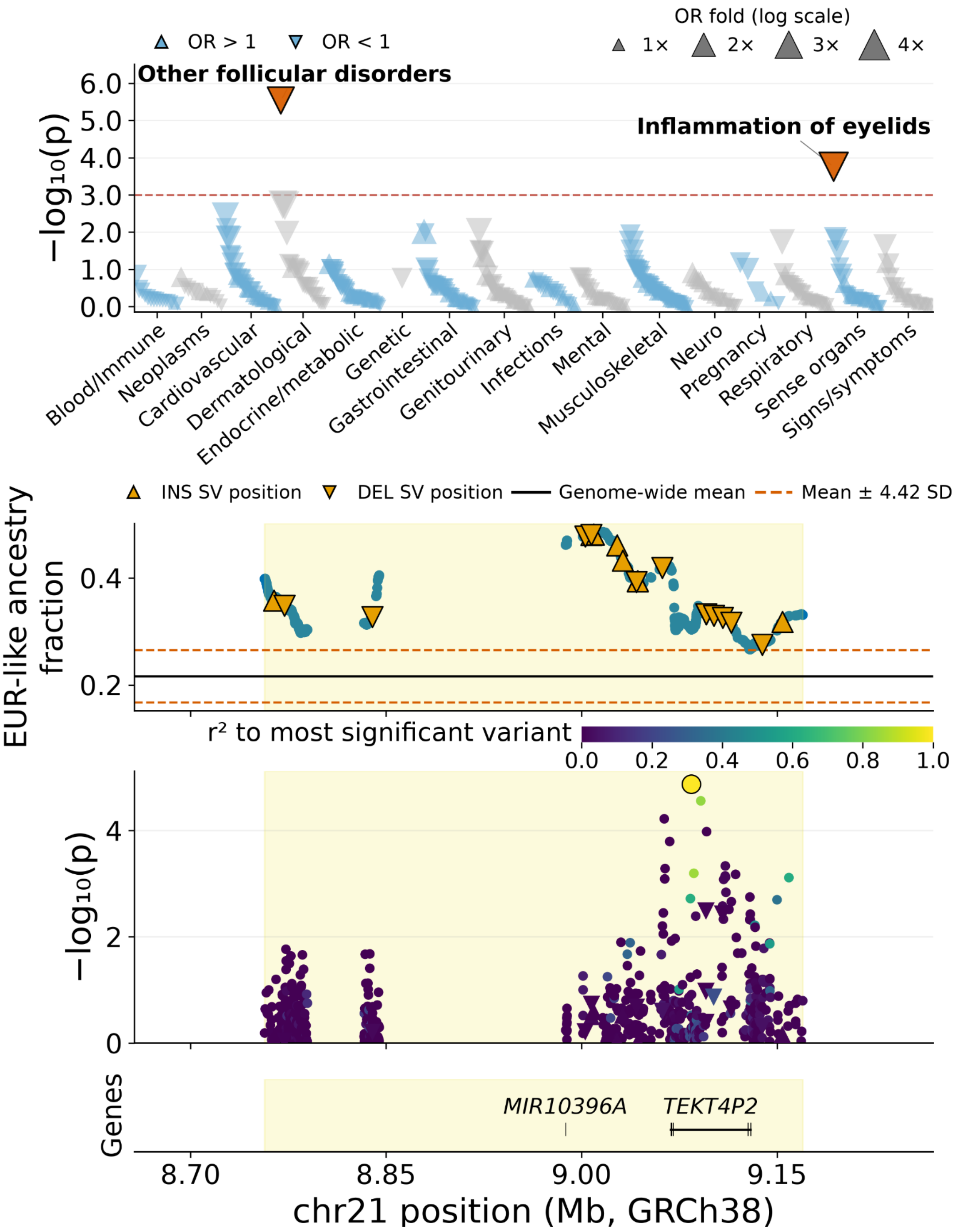


**Figure S18. Fine-mapping of the PheWAS association for phecode "other follicular disorders" (DE_682) at admixture outlier region chr21:8,756,876–9,169,773.** Top panel: PheWAS Manhattan plot across 446 phecodes in 16 categories. Upward and downward triangles represent increased and decreased odds for individual phecode outcomes, respectively. Middle panel: EUR-like ancestry fraction across markers. The outlier region is indicated in yellow. Bottom panel: Variant association signals across the locus. Variant colors indicate linkage disequilibrium ($r^2$) with the lead test variant. Gene locations are indicated.

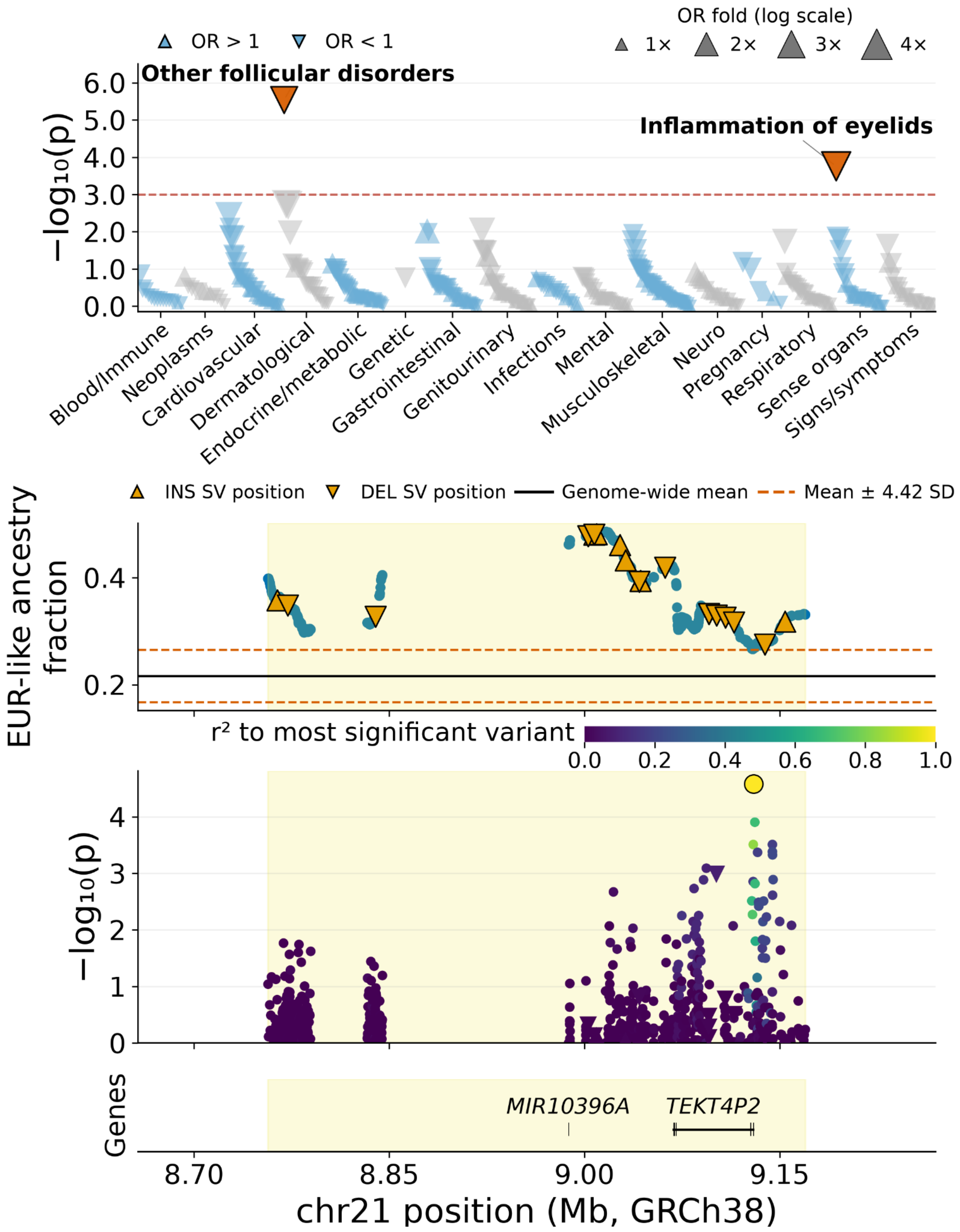


**Figure S19. Fine-mapping of the PheWAS association for phecode "inflammation of eyelids" (SO_360) at admixture outlier region chr21:8,756,876–9,169,773.** Top panel: PheWAS Manhattan plot across 446 phecodes in 16 categories. Upward and downward triangles represent increased and decreased odds for individual phecode outcomes, respectively. Middle panel: EUR-like ancestry fraction across markers. The outlier region is indicated in yellow. Bottom panel: Variant association signals across the locus. Variant colors indicate linkage disequilibrium ($r^2$) with the lead test variant. Gene locations are indicated.

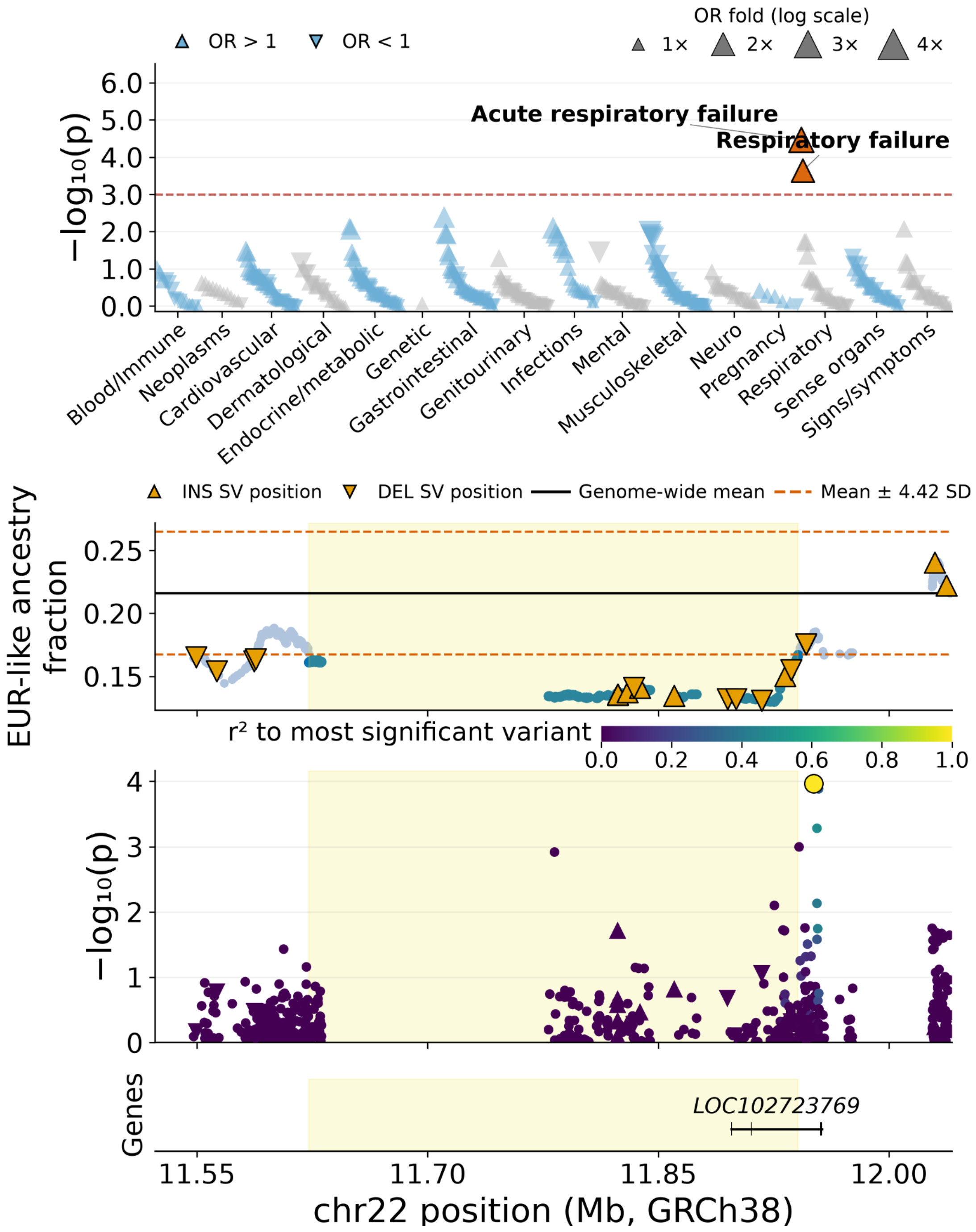


**Figure S20. Fine-mapping of the PheWAS association for phecode "respiratory failure" (RE_479.3) at admixture outlier region chr22:11,622,874–11,940,952.** Top panel: PheWAS Manhattan plot across 446 phecodes in 16 categories. Upward and downward triangles represent increased and decreased odds for individual phecode outcomes, respectively. Middle panel: EUR-like ancestry fraction across markers. The outlier region is indicated in yellow. Bottom panel: Variant association signals across the locus. Variant colors indicate linkage disequilibrium ($r^2$) with the lead test variant. Gene locations are indicated.

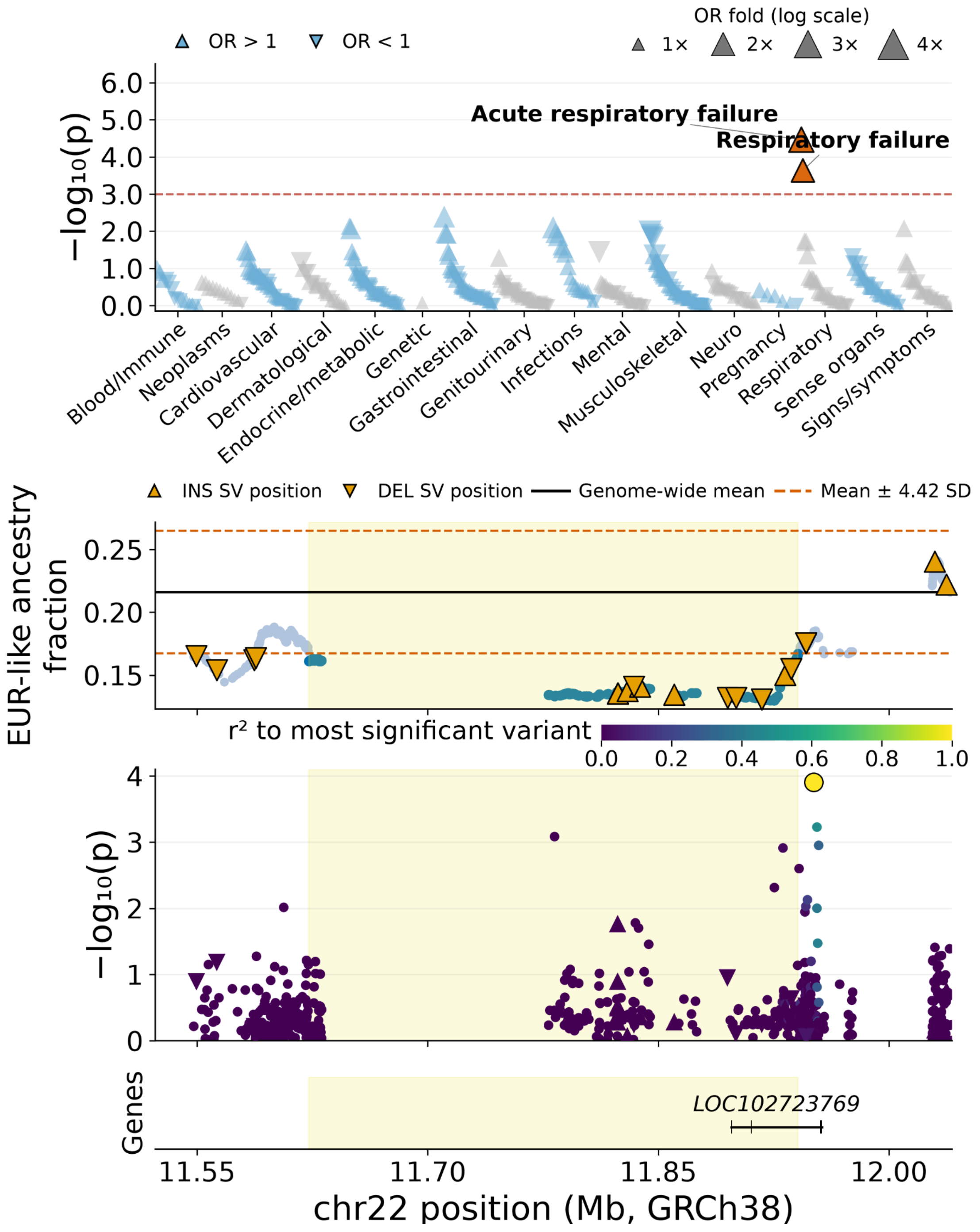


**Figure S21. Fine-mapping of the PheWAS association for phecode "acute respiratory failure" (RE_479.31) at admixture outlier region chr22:11,622,874–11,940,952.** Top panel: PheWAS Manhattan plot across 446 phecodes in 16 categories. Upward and downward triangles represent increased and decreased odds for individual phecode outcomes, respectively. Middle panel: EUR-like ancestry fraction across markers. The outlier region is indicated in yellow. Bottom panel: Variant association signals across the locus. Variant colors indicate linkage disequilibrium ($r^2$) with the lead test variant. Gene locations are indicated.

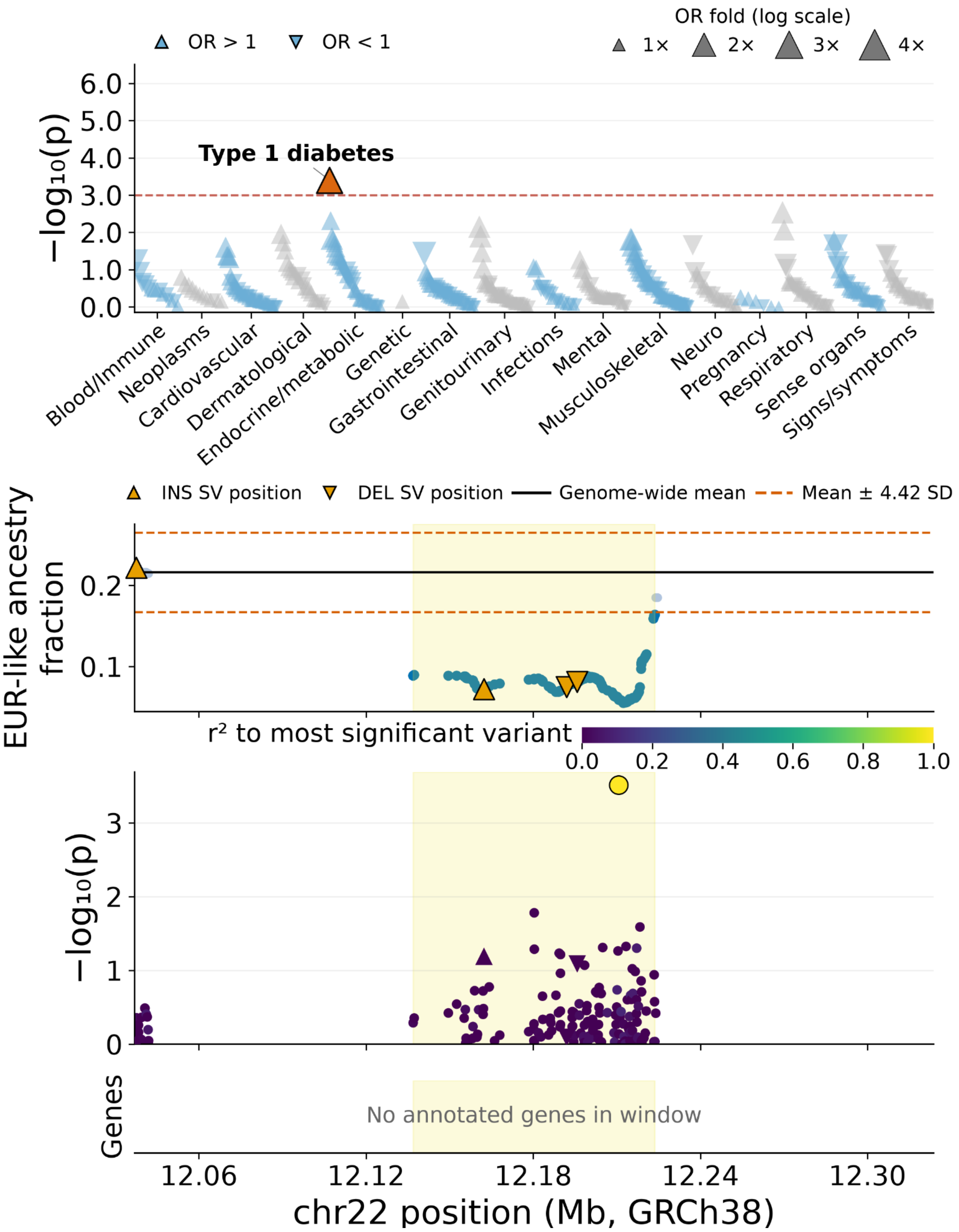


**Figure S22. Fine-mapping of the PheWAS association for phecode "type 1 diabetes" (EM_202.1) at admixture outlier region chr22:12,136,917–12,223,689.** Top panel: PheWAS Manhattan plot across 446 phecodes in 16 categories. Upward and downward triangles represent increased and decreased odds for individual phecode outcomes, respectively. Middle panel: EUR-like ancestry fraction across markers. The outlier region is indicated in yellow. Bottom panel: Variant association signals across the locus. Variant colors indicate linkage disequilibrium ($r^2$) with the lead test variant. No annotated genes occur in this window.

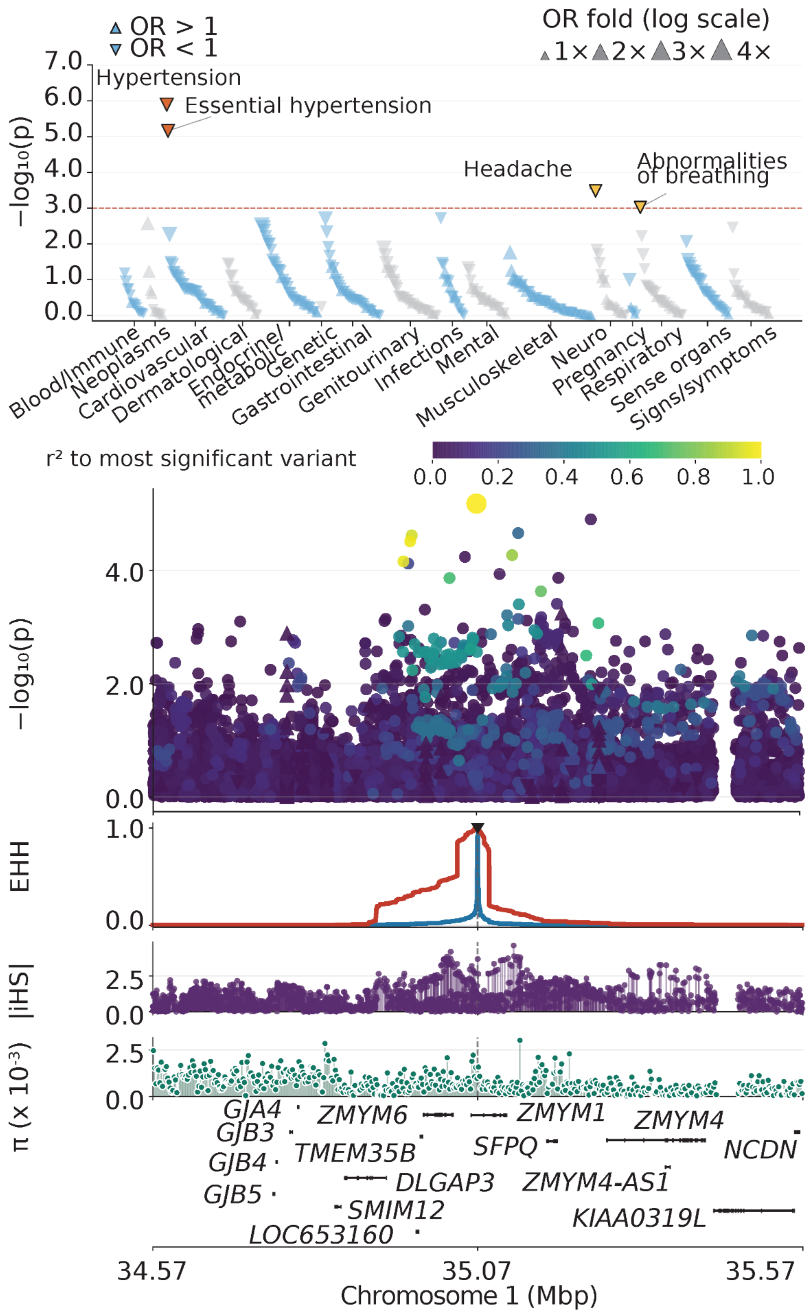


**Figure S23. Fine-mapping of the PheWAS association for "essential hypertension" at the chr1:35,070,852 C>A locus.** Top panel: PheWAS Manhattan plot across 447 phecodes in 16 categories. Upward and downward triangles represent increased and decreased odds for individual phecode outcomes, respectively. Orange-red and yellow indicate meta-analysis significance and nominal discovery significance, respectively. Second panel: Variant association signals across the locus. Variant colors indicate linkage disequilibrium ($r^2$) with the most significant variant. Third panel: Extended haplotype homozygosity (EHH) centered at the index variant. Blue and red indicate reference and alternative alleles, respectively. Fourth to sixth panels: Absolute normalized iHS scores, nucleotide diversity ($\pi$), and gene locations, respectively.

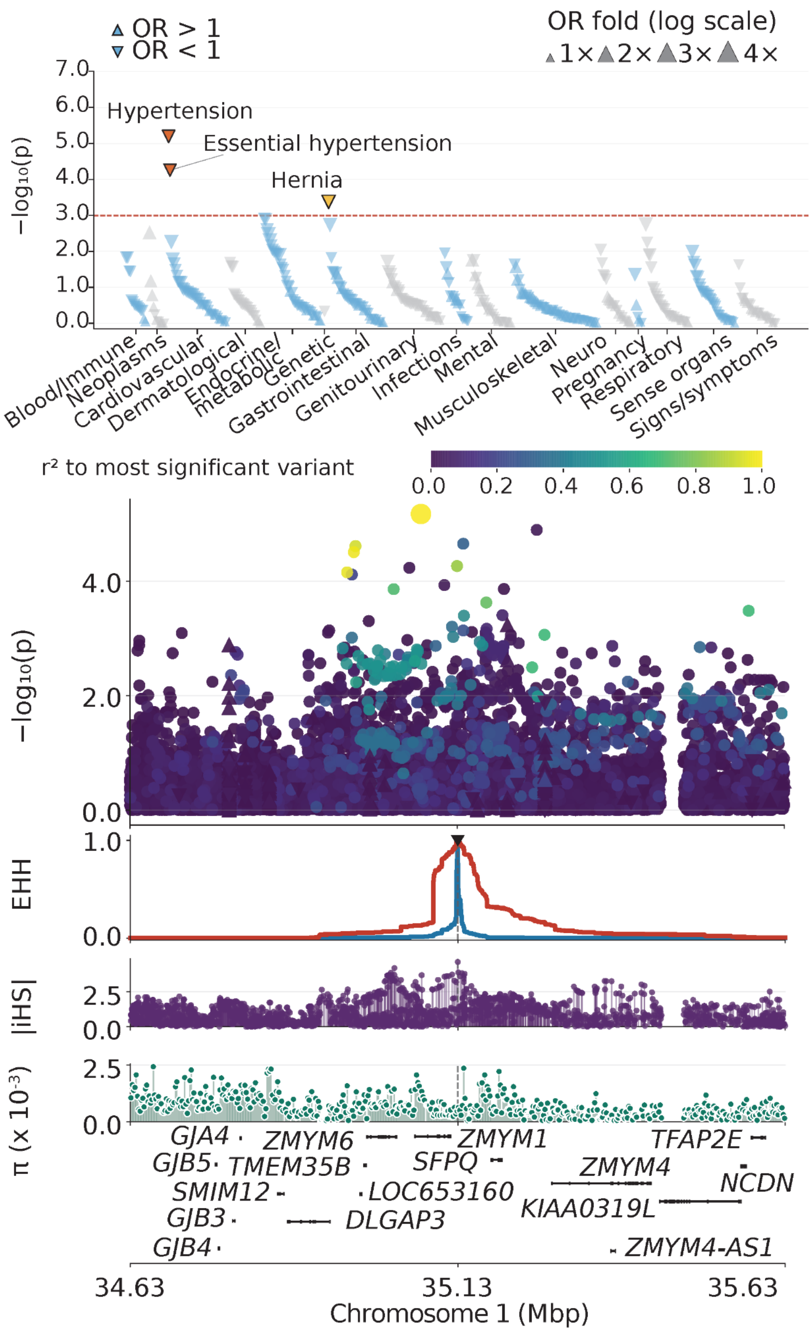


**Figure S24. Fine-mapping of the PheWAS association for "essential hypertension" at the chr1:35,126,025 A>G locus.** Top panel: PheWAS Manhattan plot across 447 phecodes in 16 categories. Upward and downward triangles represent increased and decreased odds for individual phecode outcomes, respectively. Orange-red and yellow indicate meta-analysis significance and nominal discovery significance, respectively. Second panel: Variant association signals across the locus. Variant colors indicate linkage disequilibrium ($r^2$) with the most significant variant. Third panel: Extended haplotype homozygosity (EHH) centered at the index variant. Blue and red indicate reference and alternative alleles, respectively. Fourth to sixth panels: Absolute normalized iHS scores, nucleotide diversity ($\pi$), and gene locations, respectively.

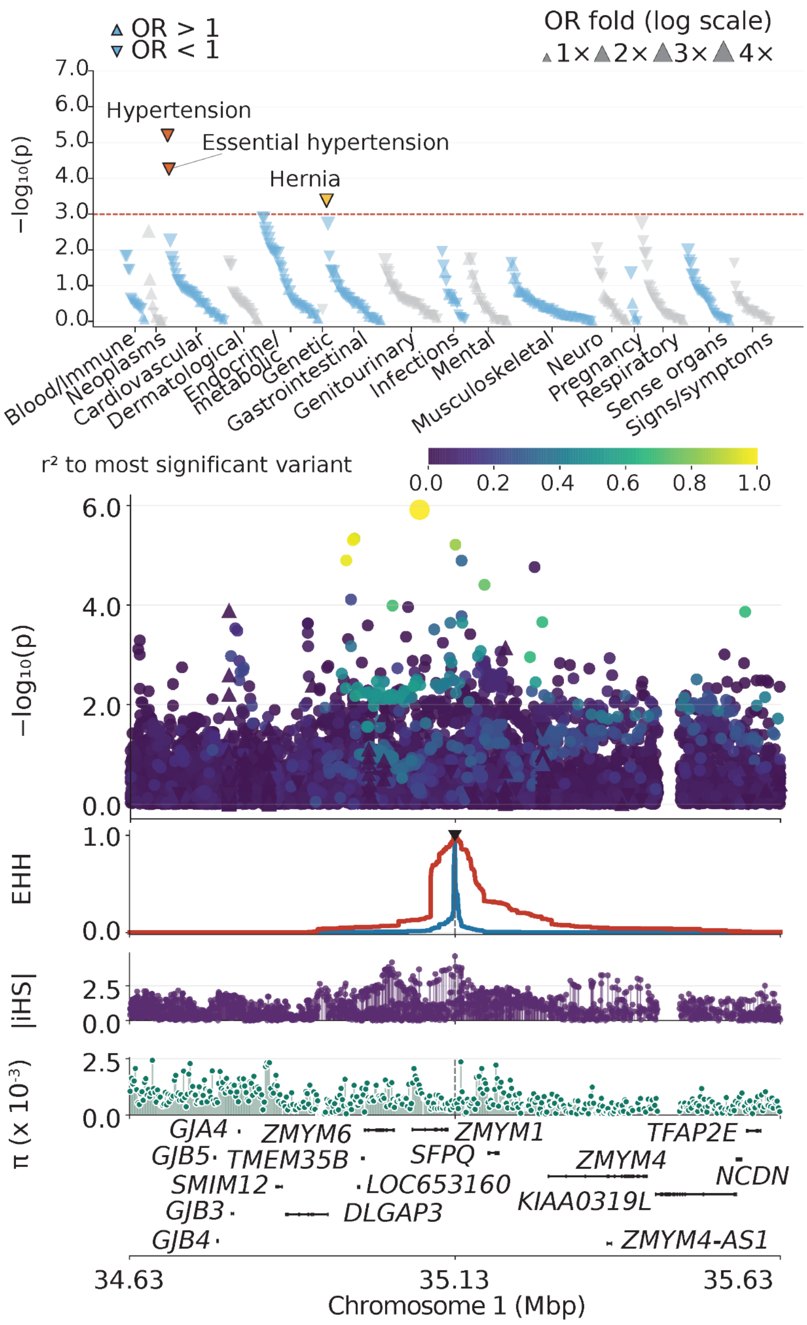


**Figure S25. Fine-mapping of the PheWAS association for "hypertension" at the chr1:35,126,025 A>G locus.** Top panel: PheWAS Manhattan plot across 447 phecodes in 16 categories. Upward and downward triangles represent increased and decreased odds for individual phecode outcomes, respectively. Orange-red and yellow indicate meta-analysis significance and nominal discovery significance, respectively. Second panel: Variant association signals across the locus. Variant colors indicate linkage disequilibrium ($r^2$) with the most significant variant. Third panel: Extended haplotype homozygosity (EHH) centered at the index variant. Blue and red indicate reference and alternative alleles, respectively. Fourth to sixth panels: Absolute normalized iHS scores, nucleotide diversity ($\pi$), and gene locations, respectively.

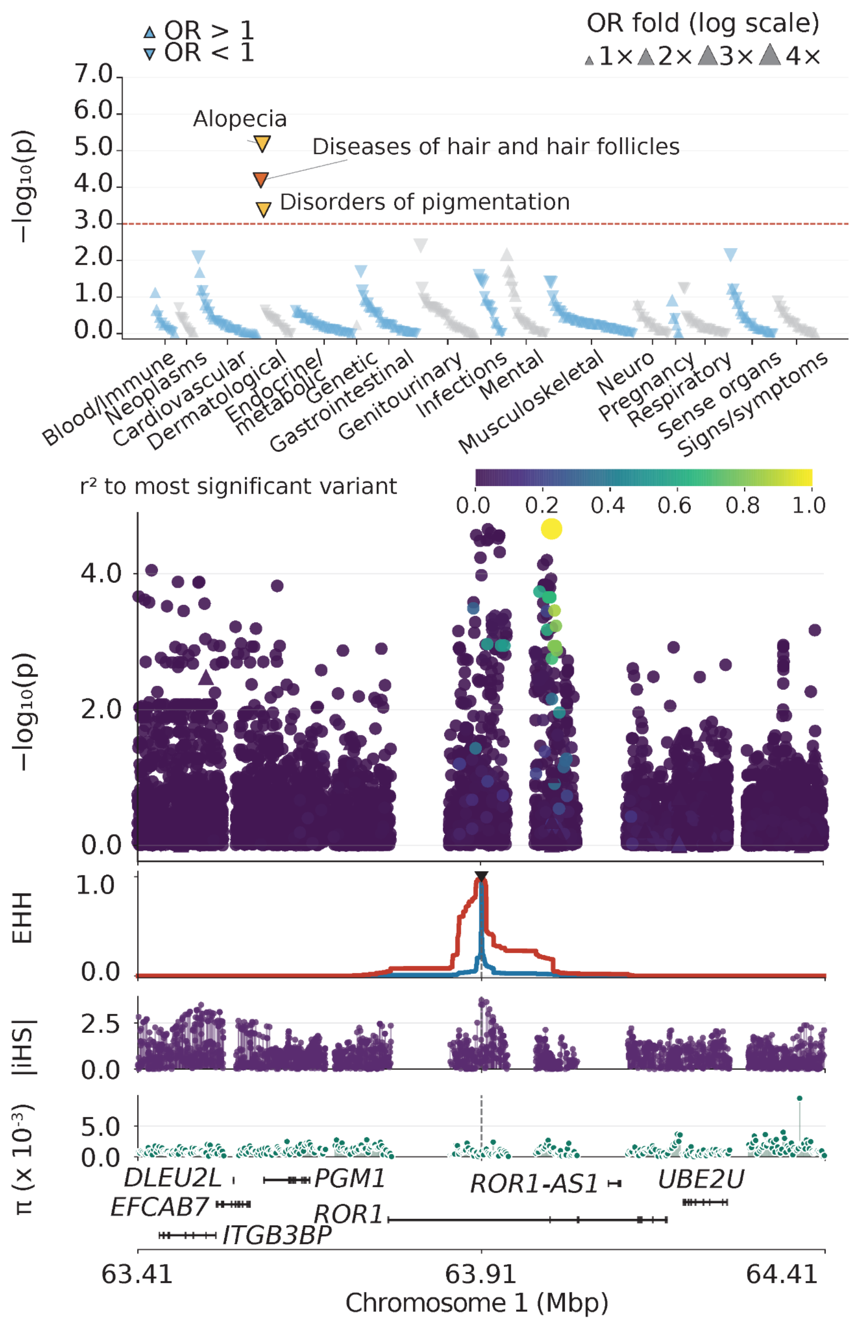


**Figure S26. Fine-mapping of the PheWAS association for "diseases of hair and hair follicles" at the chr1:63,910,558 G>T locus.** Top panel: PheWAS Manhattan plot across 447 phecodes in 16 categories. Upward and downward triangles represent increased and decreased odds for individual phecode outcomes, respectively. Orange-red and yellow indicate meta-analysis significance and nominal discovery significance, respectively. Second panel: Variant association signals across the locus. Variant colors indicate linkage disequilibrium ($r^2$) with the most significant variant. Third panel: Extended haplotype homozygosity (EHH) centered at the index variant. Blue and red indicate reference and alternative alleles, respectively. Fourth to sixth panels: Absolute normalized iHS scores, nucleotide diversity ($\pi$), and gene locations, respectively.

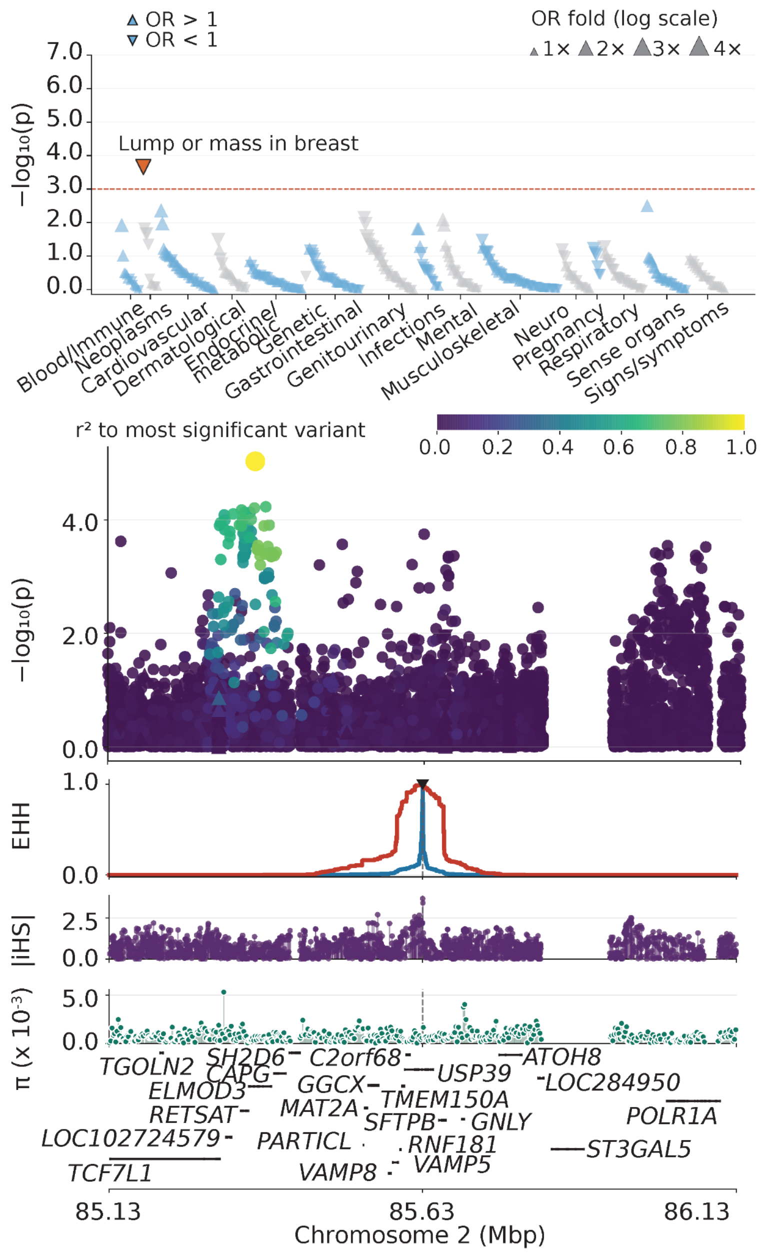


**Figure S27. Fine-mapping of the PheWAS association for "lump or mass in breast" at the chr2:85,632,712 T>A locus.** Top panel: PheWAS Manhattan plot across 447 phecodes in 16 categories. Upward and downward triangles represent increased and decreased odds for individual phecode outcomes, respectively. Orange-red and yellow indicate meta-analysis significance and nominal discovery significance, respectively. Second panel: Variant association signals across the locus. Variant colors indicate linkage disequilibrium ($r^2$) with the most significant variant. Third panel: Extended haplotype homozygosity (EHH) centered at the index variant. Blue and red indicate reference and alternative alleles, respectively. Fourth to sixth panels: Absolute normalized iHS scores, nucleotide diversity ($\pi$), and gene locations, respectively.

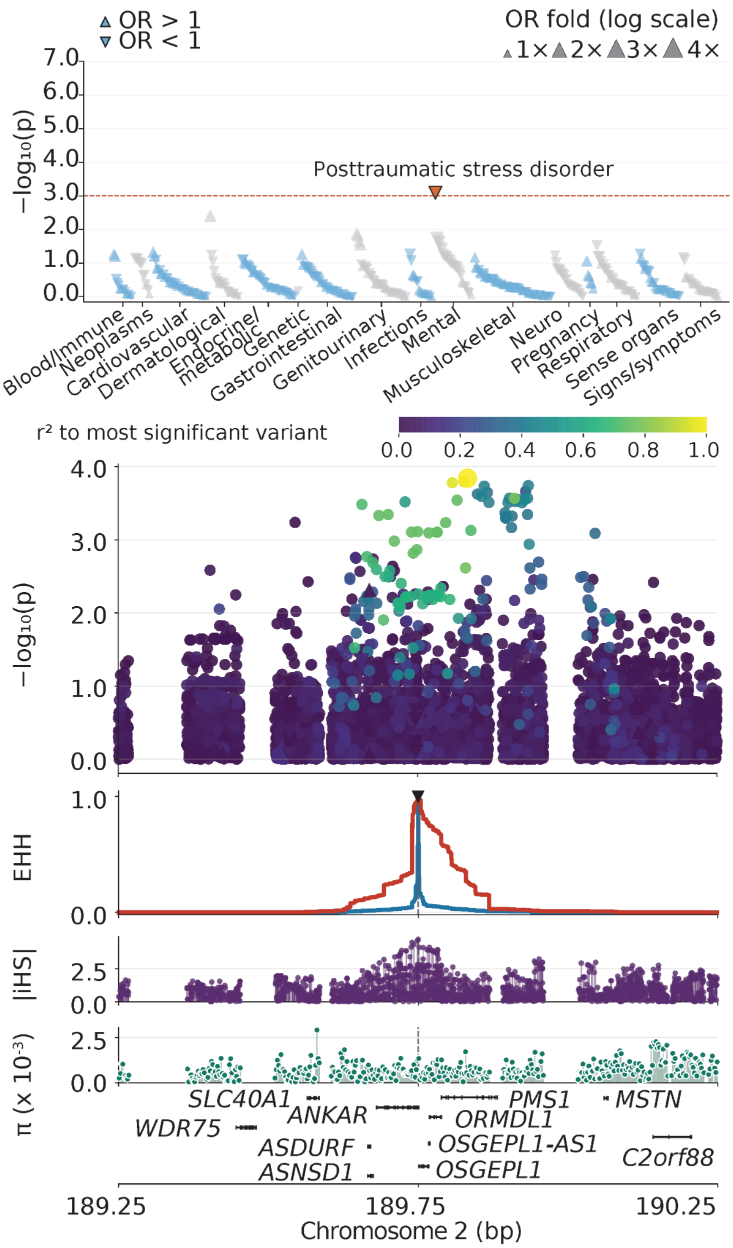


**Figure S28. Fine-mapping of the PheWAS association for "posttraumatic stress disorder" at the chr2:189,746,358 T>C locus.** Top panel: PheWAS Manhattan plot across 447 phecodes in 16 categories. Upward and downward triangles represent increased and decreased odds for individual phecode outcomes, respectively. Orange-red and yellow indicate meta-analysis significance and nominal discovery significance, respectively. Second panel: Variant association signals across the locus. Variant colors indicate linkage disequilibrium ($r^2$) with the most significant variant. Third panel: Extended haplotype homozygosity (EHH) centered at the index variant. Blue and red indicate reference and alternative alleles, respectively. Fourth to sixth panels: Absolute normalized iHS scores, nucleotide diversity ($\pi$), and gene locations, respectively.

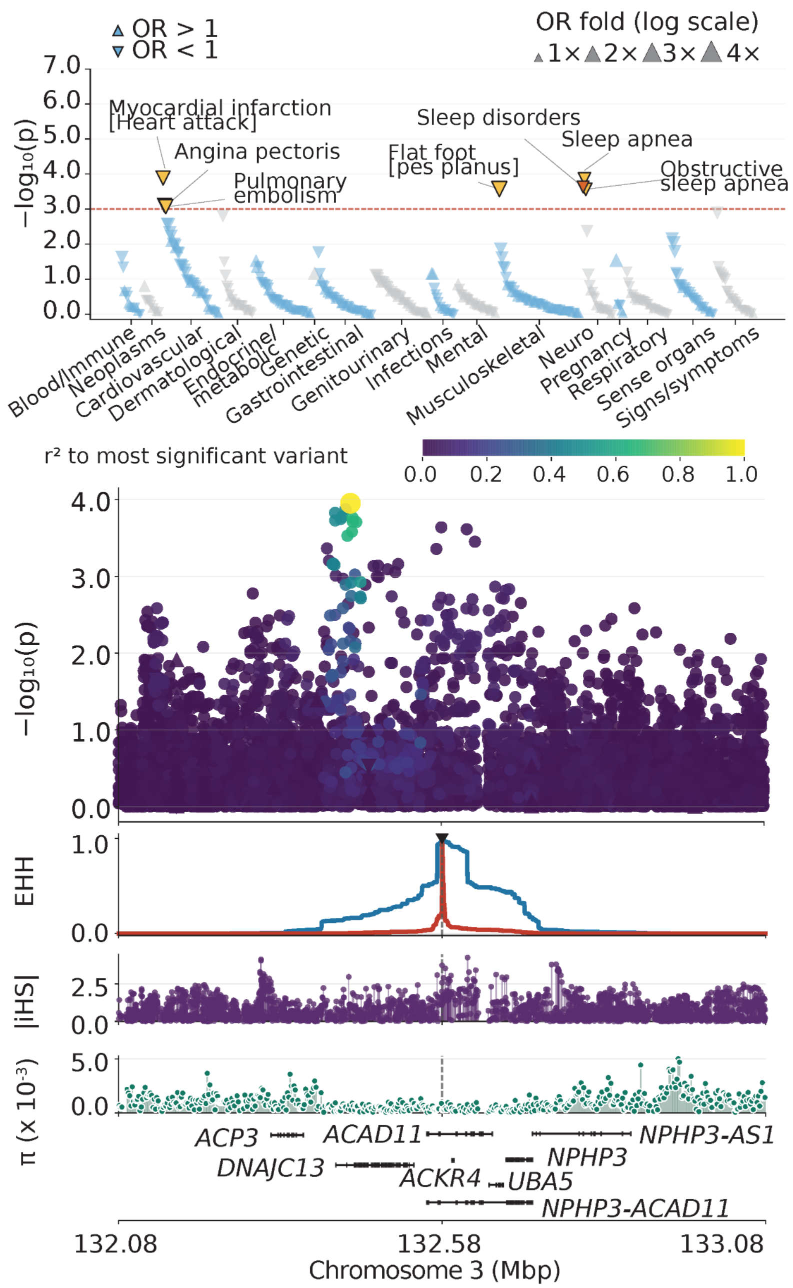


**Figure S29. Fine-mapping of the PheWAS association for "sleep disorders" at the chr3:132,582,905 A>C locus.** Top panel: PheWAS Manhattan plot across 447 phecodes in 16 categories. Upward and downward triangles represent increased and decreased odds for individual phecode outcomes, respectively. Orange-red and yellow indicate meta-analysis significance and nominal discovery significance, respectively. Second panel: Variant association signals across the locus. Variant colors indicate linkage disequilibrium ($r^2$) with the most significant variant. Third panel: Extended haplotype homozygosity (EHH) centered at the index variant. Blue and red indicate reference and alternative alleles, respectively. Fourth to sixth panels: Absolute normalized iHS scores, nucleotide diversity ($\pi$), and gene locations, respectively.

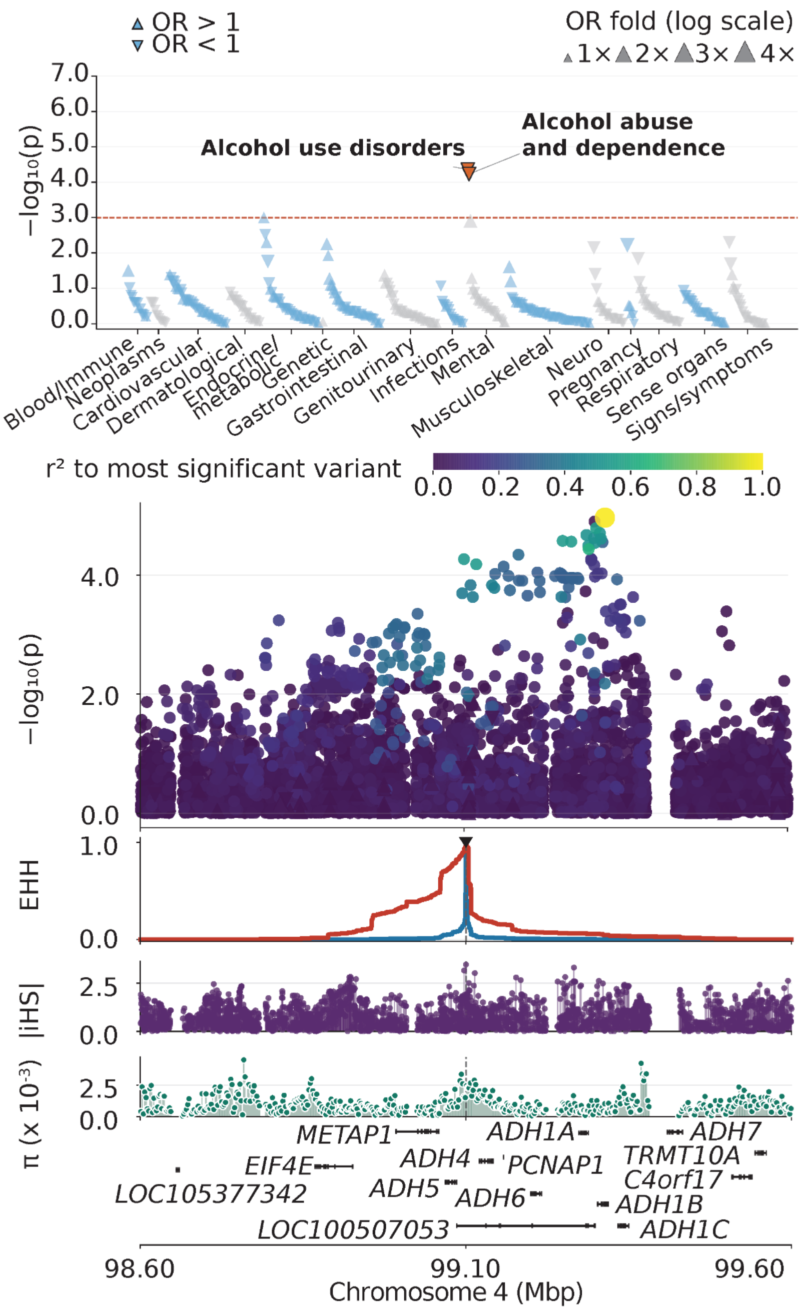


**Figure S30. Fine-mapping of the PheWAS association for "alcohol abuse and dependence" at the chr4:99,103,864 C>T locus.** Top panel: PheWAS Manhattan plot across 447 phecodes in 16 categories. Upward and downward triangles represent increased and decreased odds for individual phecode outcomes, respectively. Orange-red and yellow indicate meta-analysis significance and nominal discovery significance, respectively. Second panel: Variant association signals across the locus. Variant colors indicate linkage disequilibrium ($r^2$) with the most significant variant. Third panel: Extended haplotype homozygosity (EHH) centered at the index variant. Blue and red indicate reference and alternative alleles, respectively. Fourth to sixth panels: Absolute normalized iHS scores, nucleotide diversity ($\pi$), and gene locations, respectively.

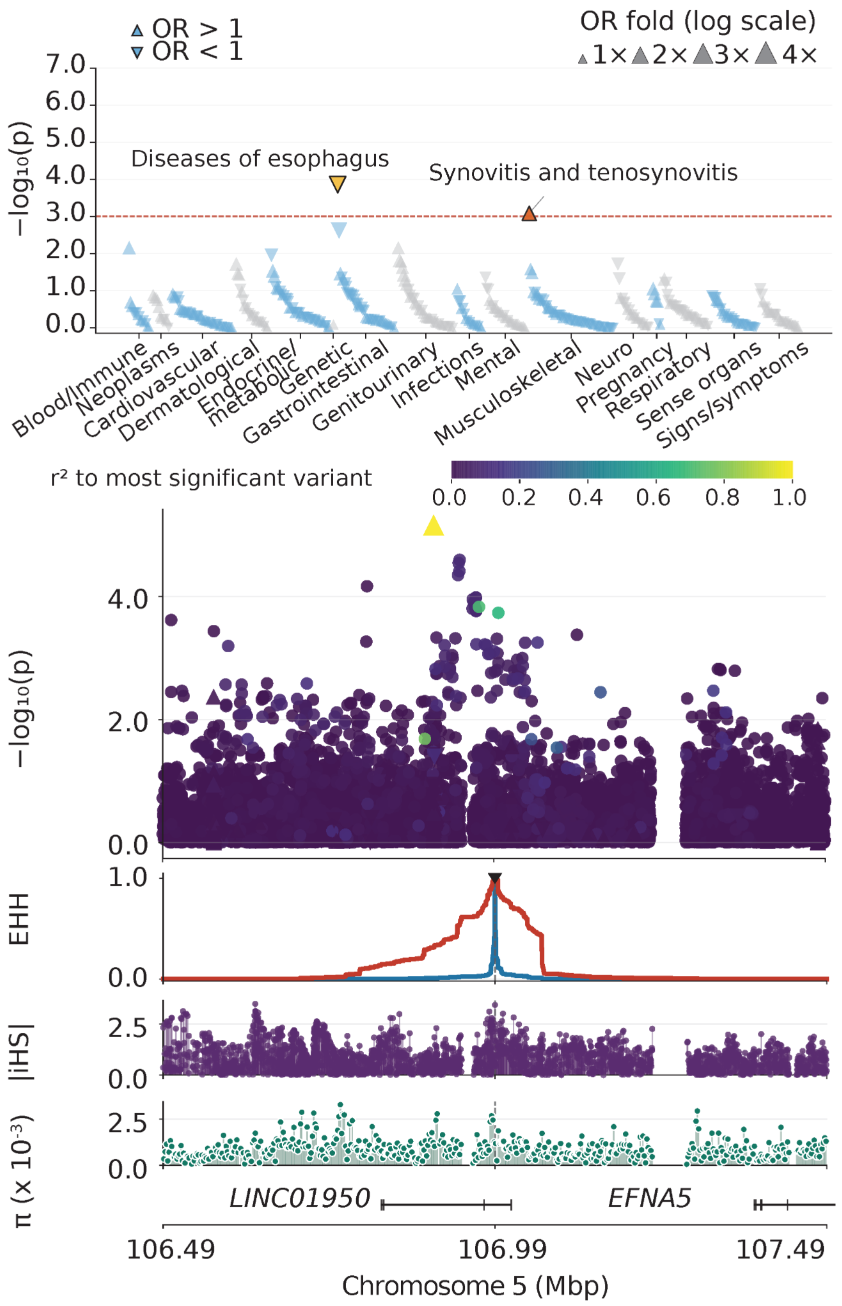


**Figure S31. Fine-mapping of the PheWAS association for "synovitis and tenosynovitis" at the chr5:106,987,252 C>T locus.** Top panel: PheWAS Manhattan plot across 447 phecodes in 16 categories. Upward and downward triangles represent increased and decreased odds for individual phecode outcomes, respectively. Orange-red and yellow indicate meta-analysis significance and nominal discovery significance, respectively. Second panel: Variant association signals across the locus. Variant colors indicate linkage disequilibrium ($r^2$) with the most significant variant. Third panel: Extended haplotype homozygosity (EHH) centered at the index variant. Blue and red indicate reference and alternative alleles, respectively. Fourth to sixth panels: Absolute normalized iHS scores, nucleotide diversity ($\pi$), and gene locations, respectively.

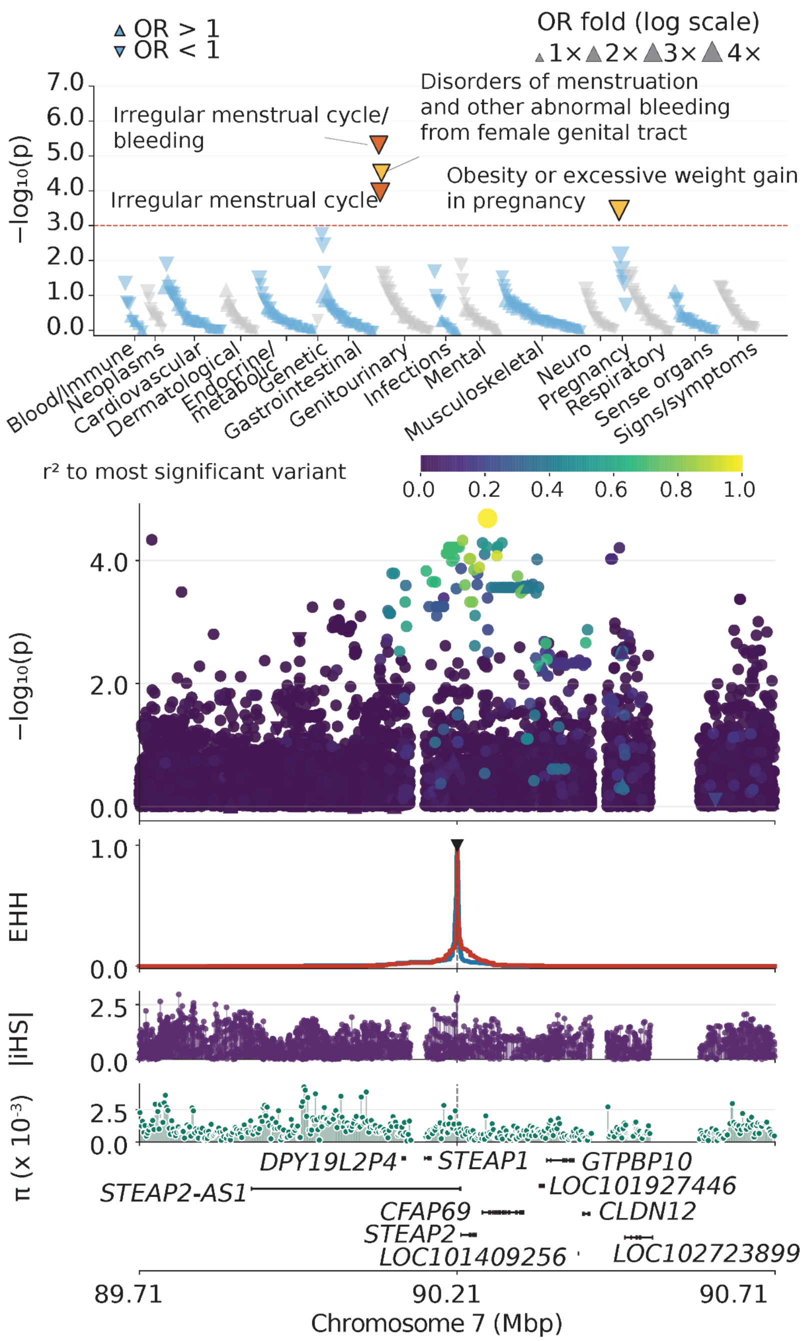


**Figure S32. Fine-mapping of the PheWAS association for "irregular menstrual cycle" at the chr7:90,206,732 G>A locus.** Top panel: PheWAS Manhattan plot across 447 phecodes in 16 categories. Upward and downward triangles represent increased and decreased odds for individual phecode outcomes, respectively. Orange-red and yellow indicate meta-analysis significance and nominal discovery significance, respectively. Second panel: Variant association signals across the locus. Variant colors indicate linkage disequilibrium ($r^2$) with the most significant variant. Third panel: Extended haplotype homozygosity (EHH) centered at the index variant. Blue and red indicate reference and alternative alleles, respectively. Fourth to sixth panels: Absolute normalized iHS scores, nucleotide diversity ($\pi$), and gene locations, respectively.

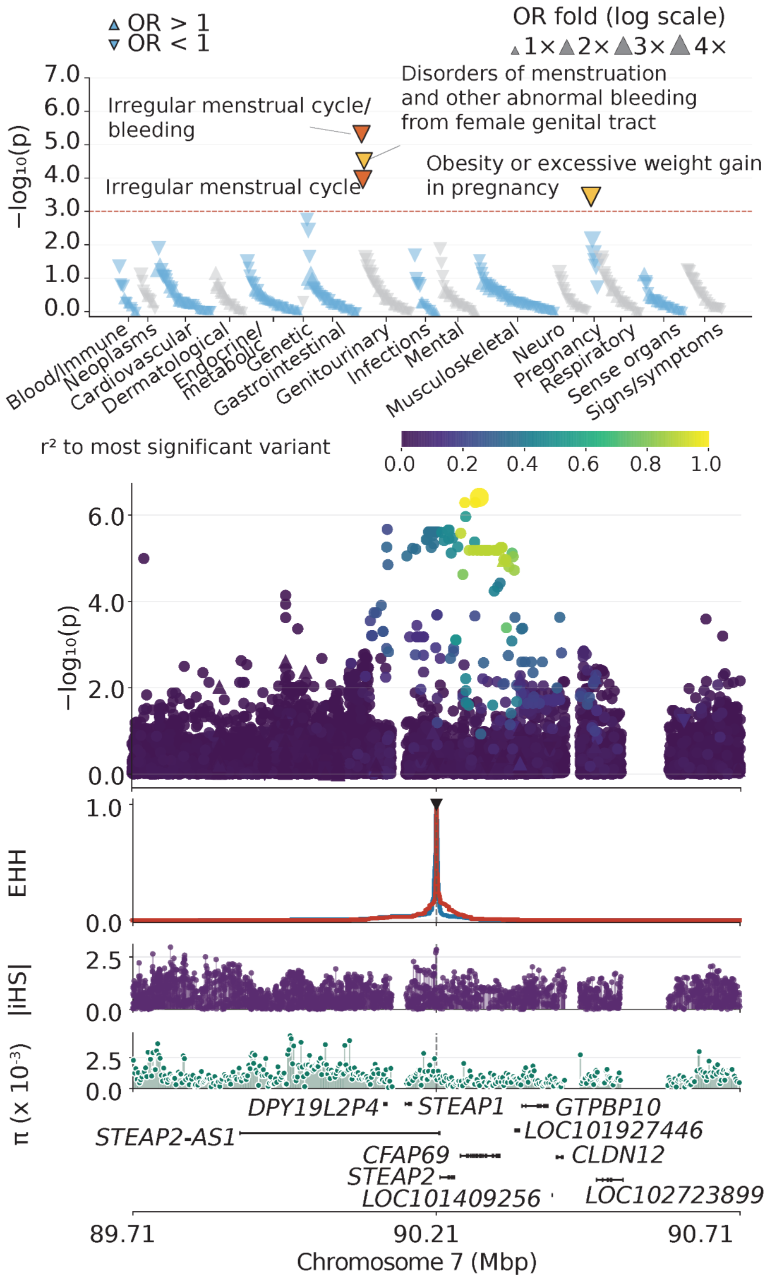


**Figure S33. Fine-mapping of the PheWAS association for "irregular menstrual cycle bleeding" at the chr7:90,206,732 G>A locus.** Top panel: PheWAS Manhattan plot across 447 phecodes in 16 categories. Upward and downward triangles represent increased and decreased odds for individual phecode outcomes, respectively. Orange-red and yellow indicate meta-analysis significance and nominal discovery significance, respectively. Second panel: Variant association signals across the locus. Variant colors indicate linkage disequilibrium ($r^2$) with the most significant variant. Third panel: Extended haplotype homozygosity (EHH) centered at the index variant. Blue and red indicate reference and alternative alleles, respectively. Fourth to sixth panels: Absolute normalized iHS scores, nucleotide diversity ($\pi$), and gene locations, respectively.

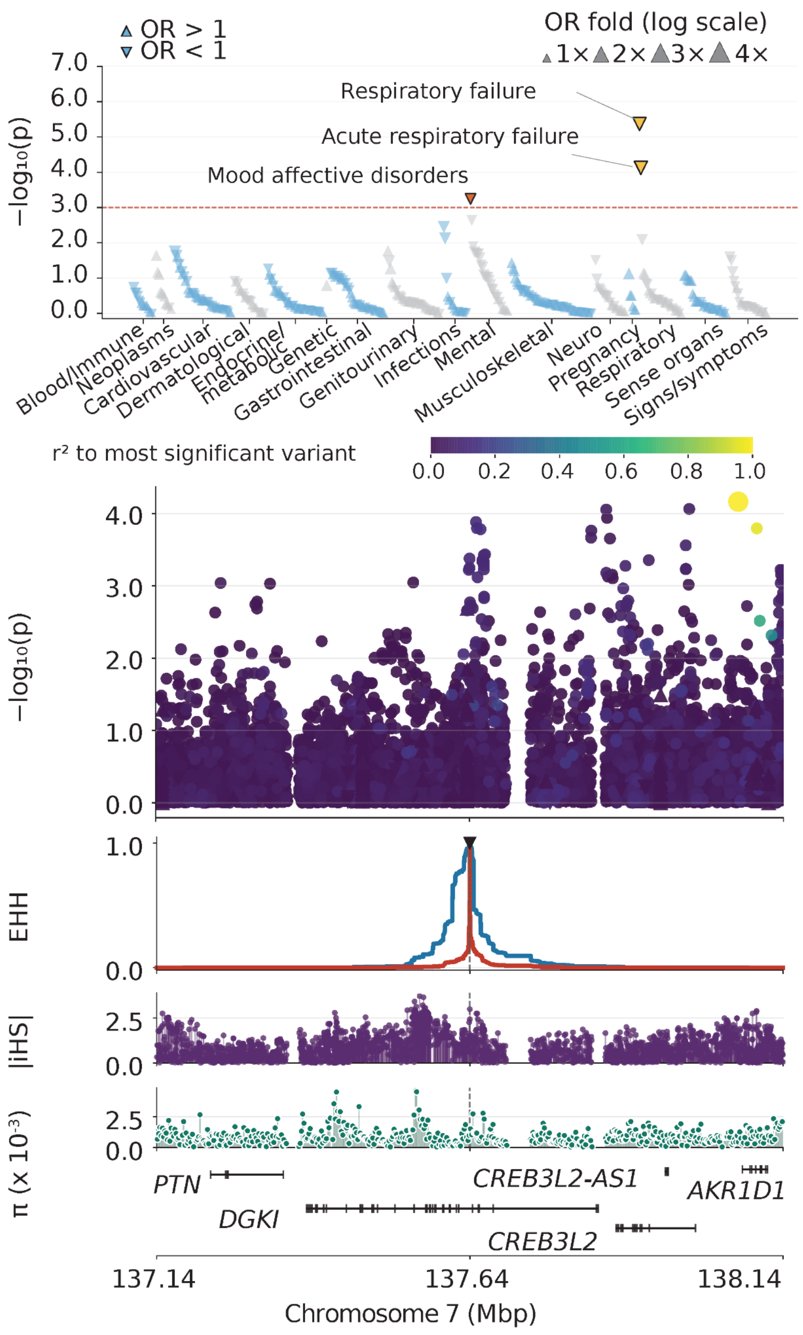


**Figure S34. Fine-mapping of the PheWAS association for "mood disorders" at the chr7:137,642,303 C>G locus.** Top panel: PheWAS Manhattan plot across 447 phecodes in 16 categories. Upward and downward triangles represent increased and decreased odds for individual phecode outcomes, respectively. Orange-red and yellow indicate meta-analysis significance and nominal discovery significance, respectively. Second panel: Variant association signals across the locus. Variant colors indicate linkage disequilibrium ($r^2$) with the most significant variant. Third panel: Extended haplotype homozygosity (EHH) centered at the index variant. Blue and red indicate reference and alternative alleles, respectively. Fourth to sixth panels: Absolute normalized iHS scores, nucleotide diversity ($\pi$), and gene locations, respectively.

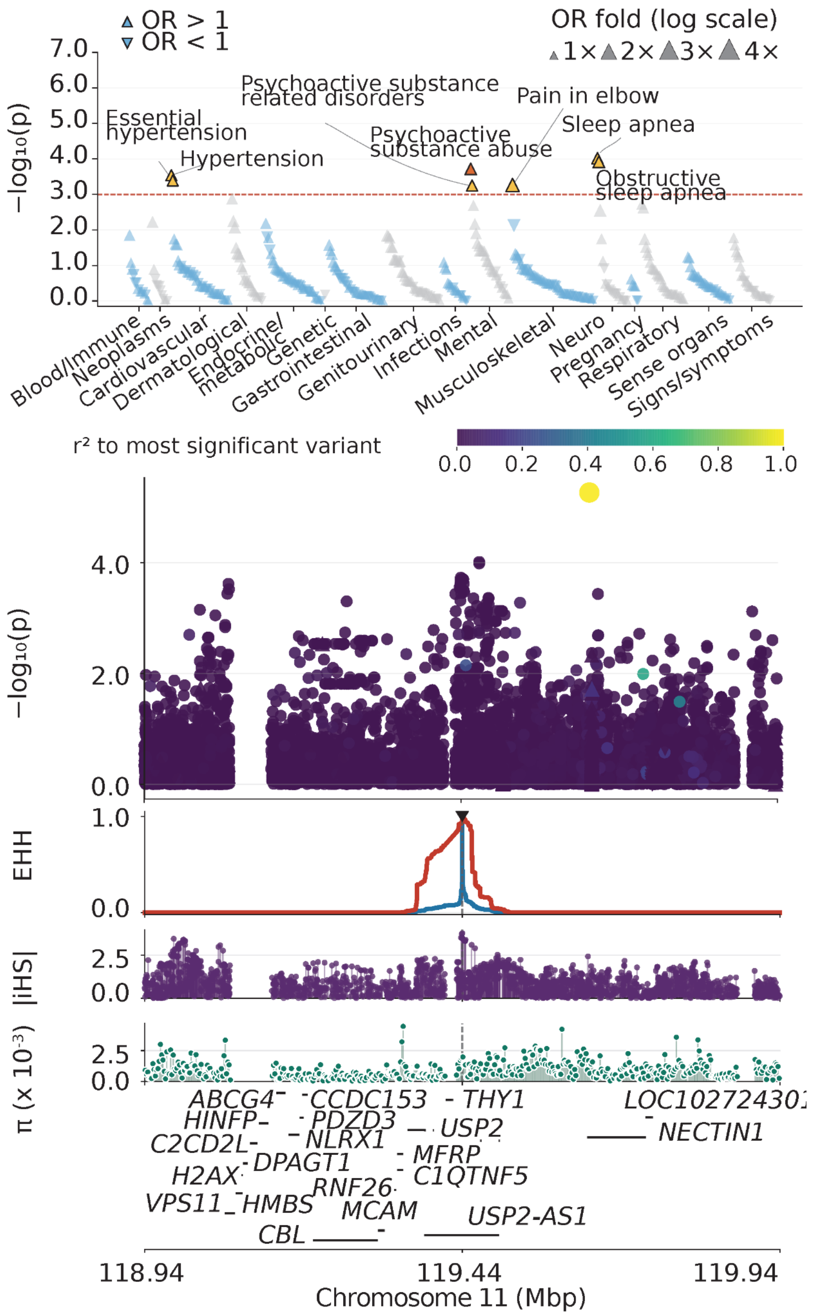


**Figure S35. Fine-mapping of the PheWAS association for "psychoactive substance abuse" at the chr11:119,441,304 G>A locus.** Top panel: PheWAS Manhattan plot across 447 phecodes in 16 categories. Upward and downward triangles represent increased and decreased odds for individual phecode outcomes, respectively. Orange-red and yellow indicate meta-analysis significance and nominal discovery significance, respectively. Second panel: Variant association signals across the locus. Variant colors indicate linkage disequilibrium ($r^2$) with the most significant variant. Third panel: Extended haplotype homozygosity (EHH) centered at the index variant. Blue and red indicate reference and alternative alleles, respectively. Fourth to sixth panels: Absolute normalized iHS scores, nucleotide diversity ($\pi$), and gene locations, respectively.

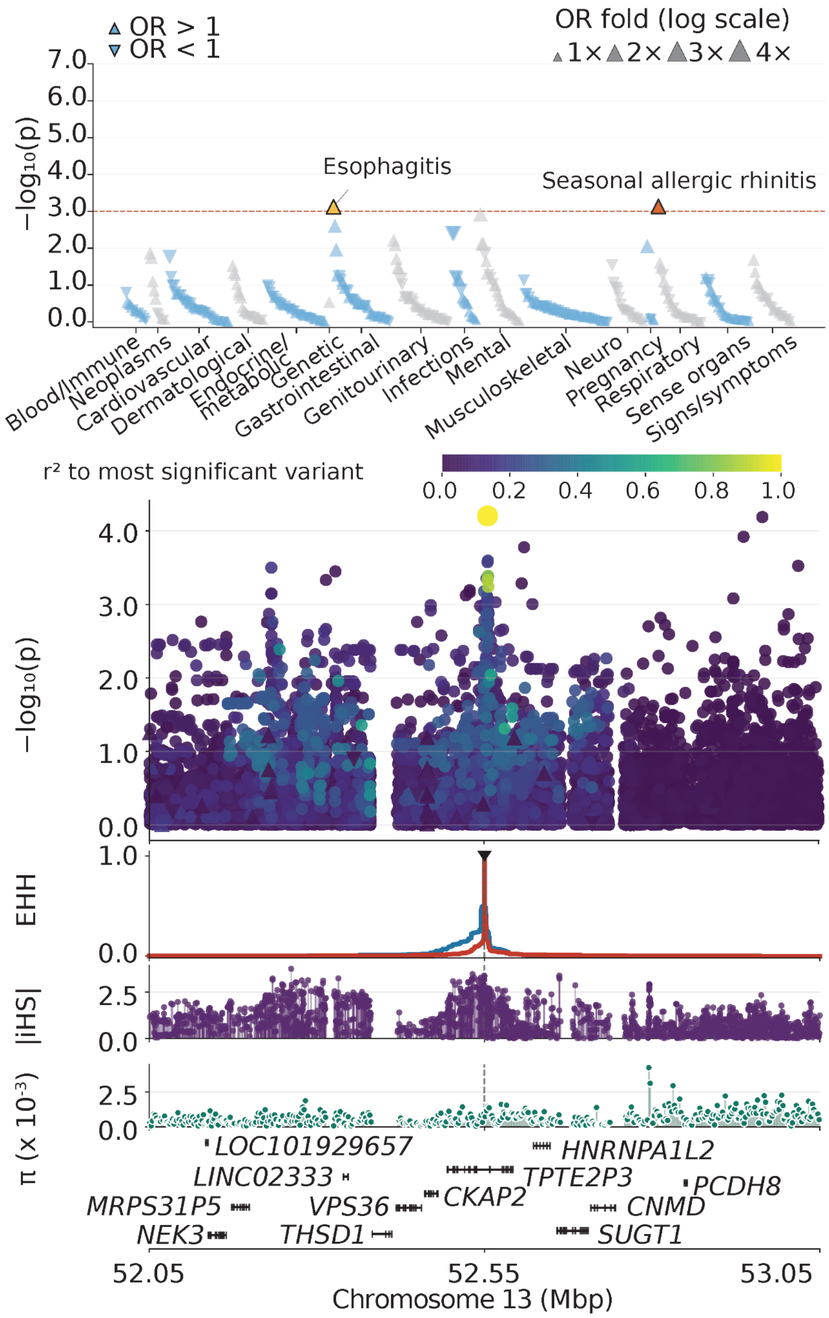


**Figure S36. Fine-mapping of the PheWAS association for "seasonal allergic rhinitis" at the chr13:52,545,567 C>A locus.** Top panel: PheWAS Manhattan plot across 447 phecodes in 16 categories. Upward and downward triangles represent increased and decreased odds for individual phecode outcomes, respectively. Orange-red and yellow indicate meta-analysis significance and nominal discovery significance, respectively. Second panel: Variant association signals across the locus. Variant colors indicate linkage disequilibrium ($r^2$) with the most significant variant. Third panel: Extended haplotype homozygosity (EHH) centered at the index variant. Blue and red indicate reference and alternative alleles, respectively. Fourth to sixth panels: Absolute normalized iHS scores, nucleotide diversity ($\pi$), and gene locations, respectively.

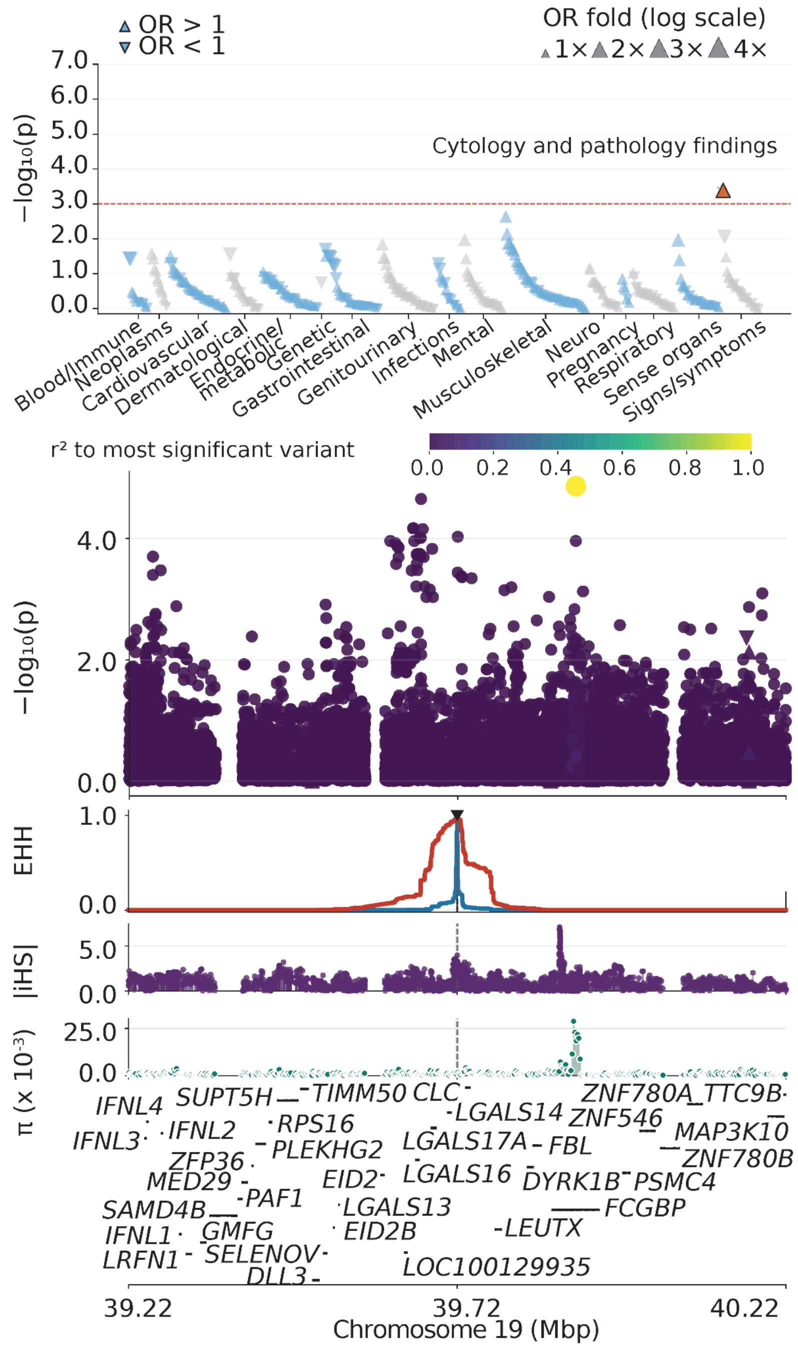


**Figure S37. Fine-mapping of the PheWAS association for "cytology and pathology findings" at the chr19:39,719,525 G>C locus.** Top panel: PheWAS Manhattan plot across 447 phecodes in 16 categories. Upward and downward triangles represent increased and decreased odds for individual phecode outcomes, respectively. Orange-red and yellow indicate meta-analysis significance and nominal discovery significance, respectively. Second panel: Variant association signals across the locus. Variant colors indicate linkage disequilibrium ($r^2$) with the most significant variant. Third panel: Extended haplotype homozygosity (EHH) centered at the index variant. Blue and red indicate reference and alternative alleles, respectively. Fourth to sixth panels: Absolute normalized iHS scores, nucleotide diversity ($\pi$), and gene locations, respectively.

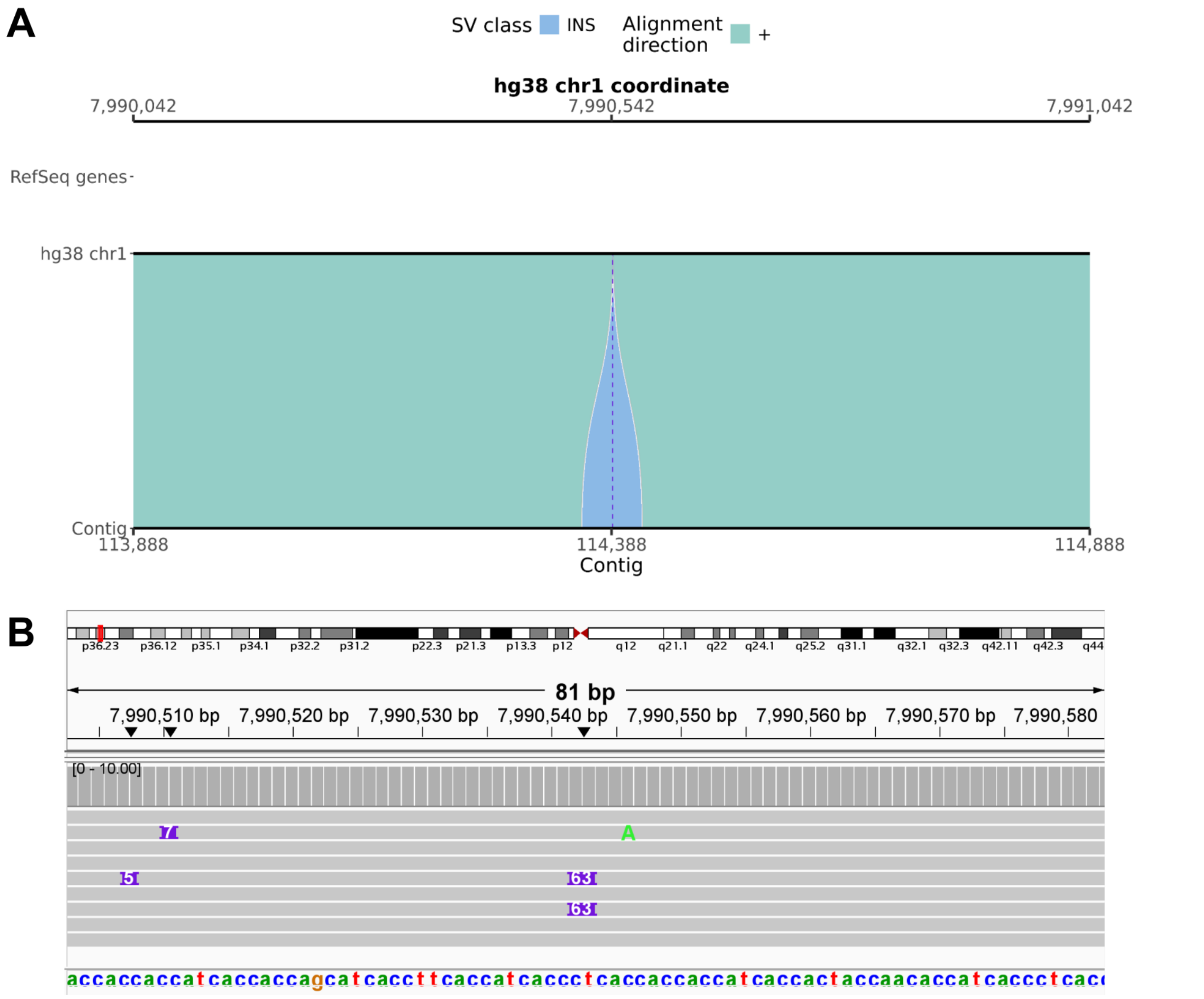


**Figure S38. svbyeye and IGV visualization of the 63-bp insertion at chr1:7,990,542.**

**(A)** Alignment of the hg38 reference sequence (top track) with a representative contig from an assembly of a sample carrying the SV (bottom track). Shaded blocks show the alignment and highlight the structural difference. The legend indicates SV class and alignment direction. Gene IDs are shown if present. The axes give reference and contig coordinates, respectively. **(B)** Read alignment at the SV locus. Coverage is shown above the reads. The SV is indicated with the SV length in purple. The hg38 reference bases are shown, with gene IDs if present.

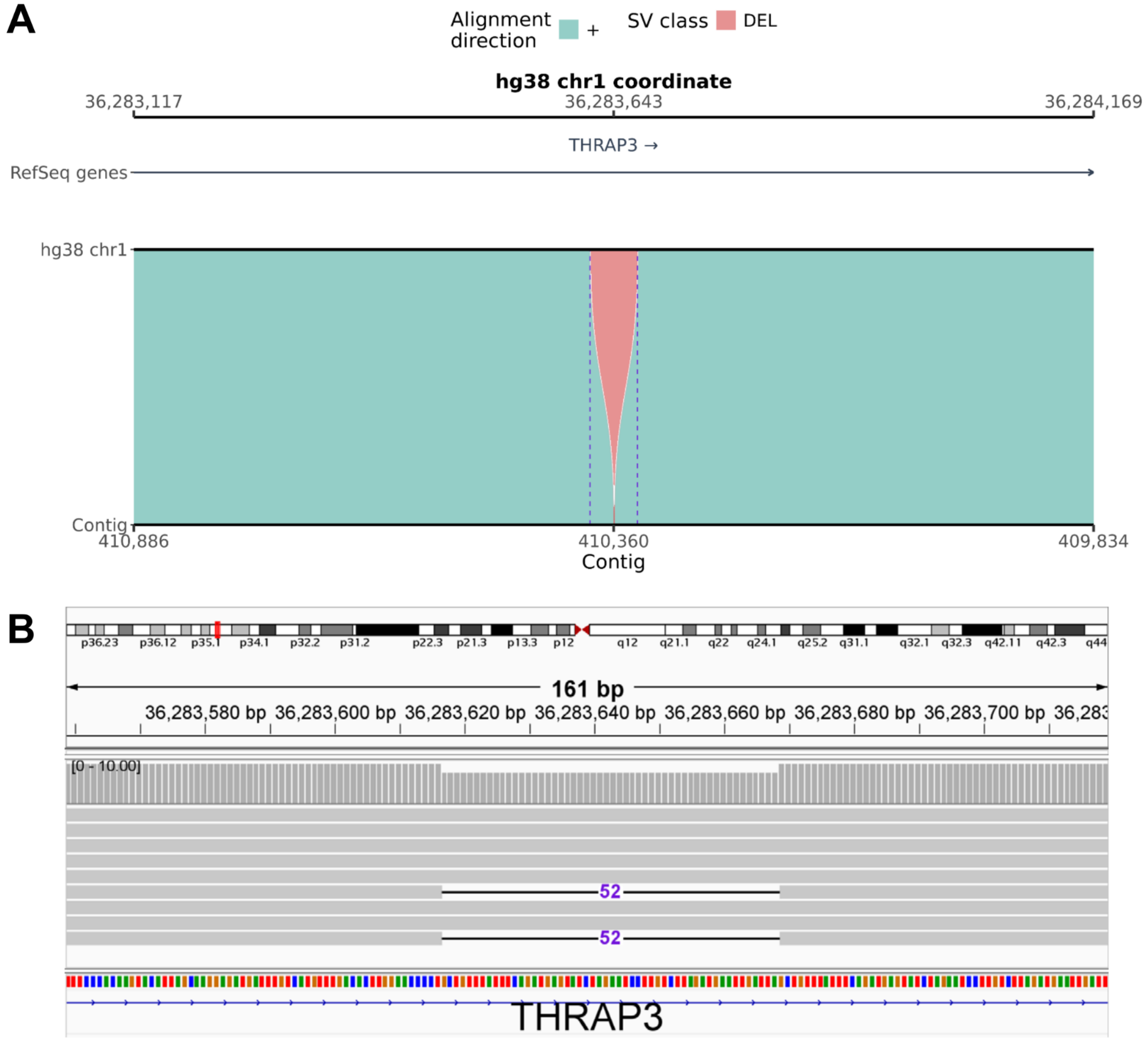


**Figure S39. svbyeye and IGV visualization of the 52-bp deletion at chr1:36,283,616.**

**(A)** Alignment of the hg38 reference sequence (top track) with a representative contig from an assembly of a sample carrying the SV (bottom track). Shaded blocks show the alignment and highlight the structural difference. The legend indicates SV class and alignment direction. Gene IDs are shown if present. The axes give reference and contig coordinates, respectively. **(B)** Read alignment at the SV locus. Coverage is shown above the reads. The SV is indicated with the SV length in purple. The hg38 reference bases are shown, with gene IDs if present.

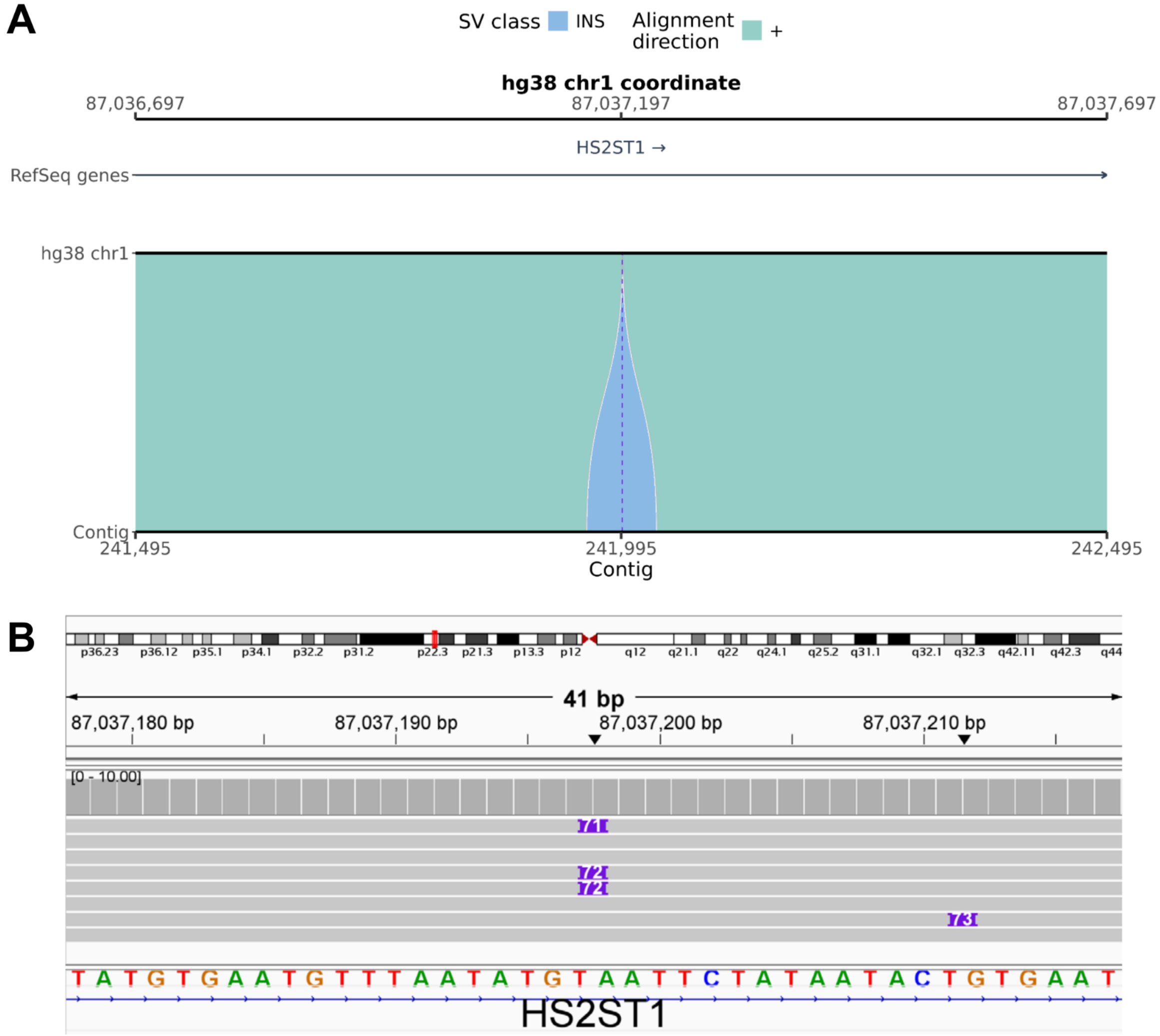


**Figure S40. svbyeye and IGV visualization of the 72-bp insertion at chr1:87,037,197.**

**(A)** Alignment of the hg38 reference sequence (top track) with a representative contig from an assembly of a sample carrying the SV (bottom track). Shaded blocks show the alignment and highlight the structural difference. The legend indicates SV class and alignment direction. Gene IDs are shown if present. The axes give reference and contig coordinates, respectively. **(B)** Read alignment at the SV locus. Coverage is shown above the reads. The SV is indicated with the SV length in purple. The hg38 reference bases are shown, with gene IDs if present.

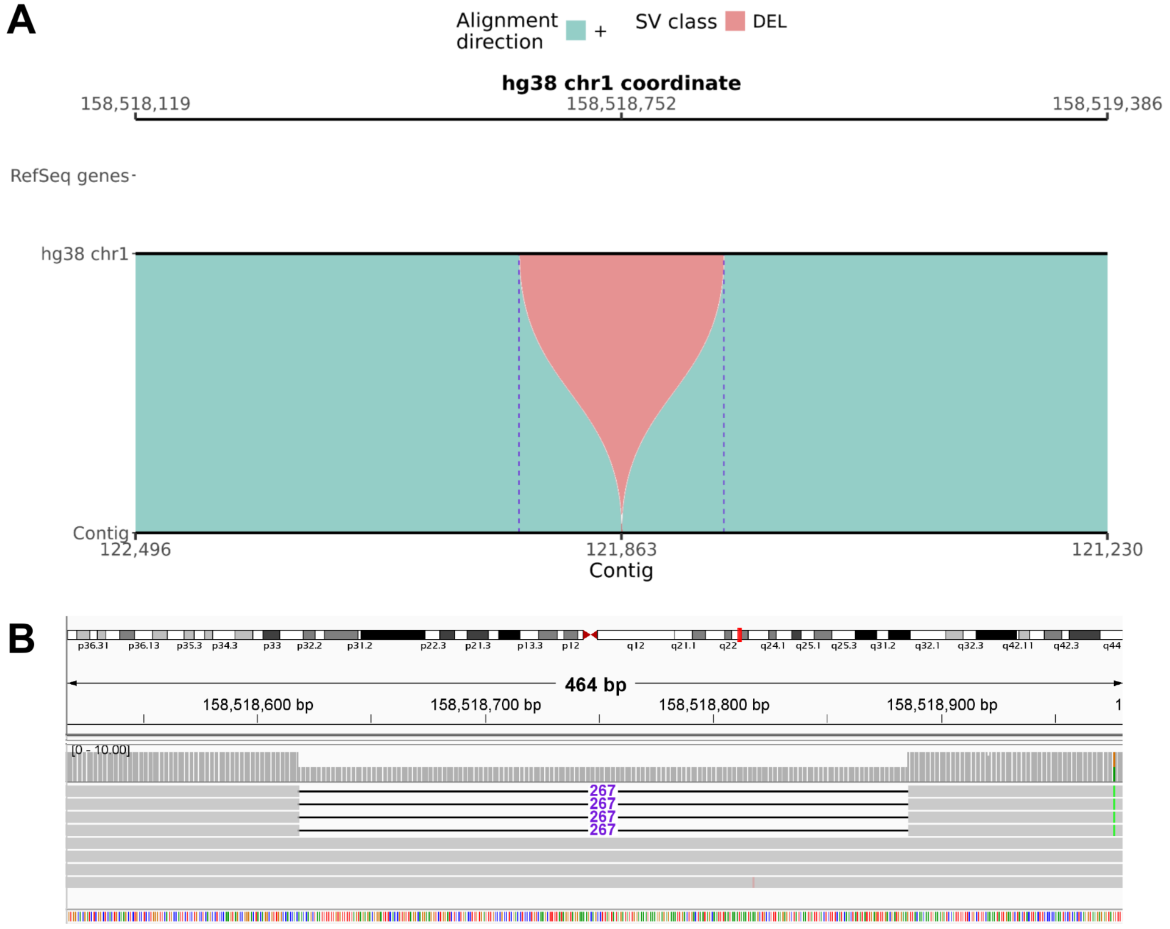


**Figure S41. svbyeye and IGV visualization of the 267-bp deletion at chr1:158,518,618.**

**(A)** Alignment of the hg38 reference sequence (top track) with a representative contig from an assembly of a sample carrying the SV (bottom track). Shaded blocks show the alignment and highlight the structural difference. The legend indicates SV class and alignment direction. Gene IDs are shown if present. The axes give reference and contig coordinates, respectively. **(B)** Read alignment at the SV locus. Coverage is shown above the reads. The SV is indicated with the SV length in purple. The hg38 reference bases are shown, with gene IDs if present.

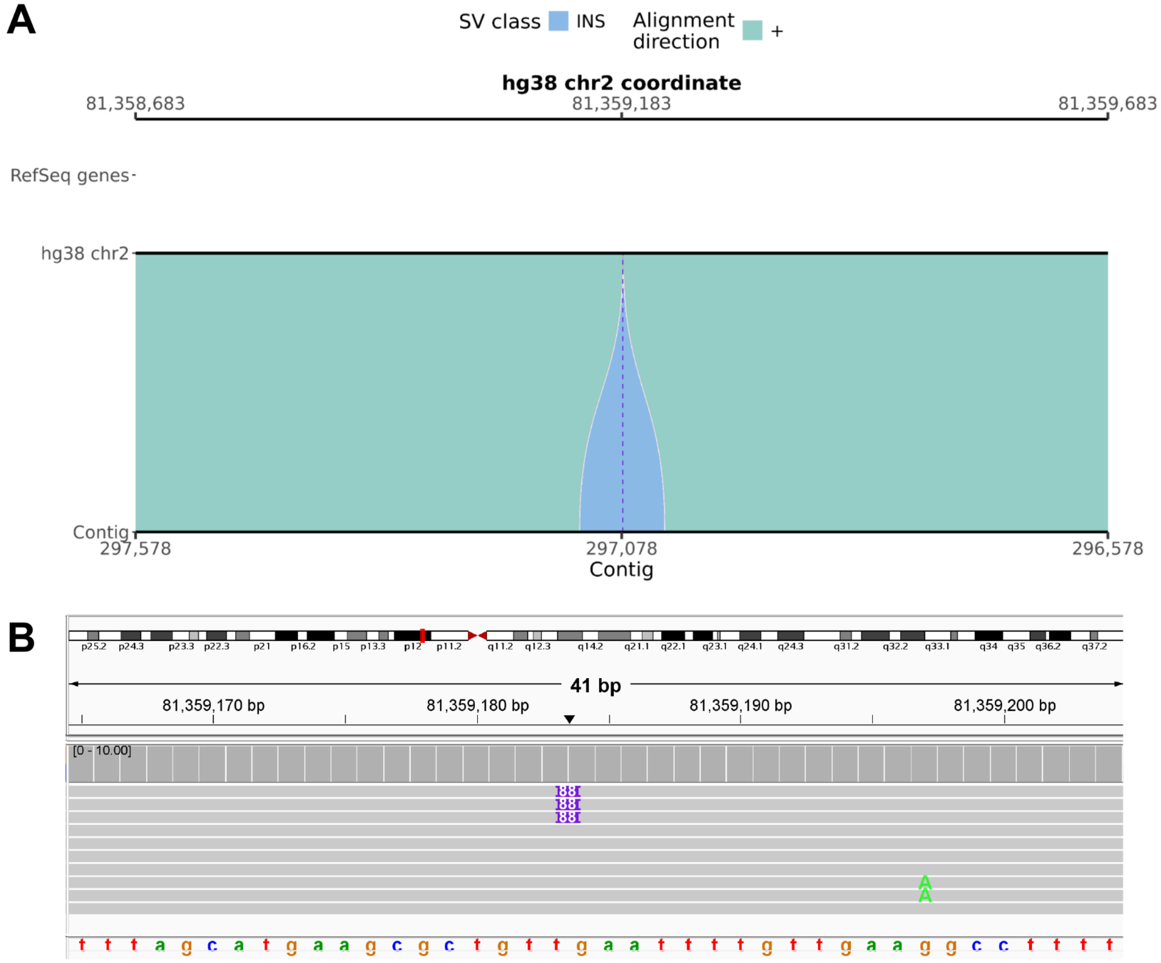


**Figure S42. svbyeye and IGV visualization of the 88-bp insertion at chr2:81,359,183.**

**(A)** Alignment of the hg38 reference sequence (top track) with a representative contig from an assembly of a sample carrying the SV (bottom track). Shaded blocks show the alignment and highlight the structural difference. The legend indicates SV class and alignment direction. Gene IDs are shown if present. The axes give reference and contig coordinates, respectively. **(B)** Read alignment at the SV locus. Coverage is shown above the reads. The SV is indicated with the SV length in purple. The hg38 reference bases are shown, with gene IDs if present.

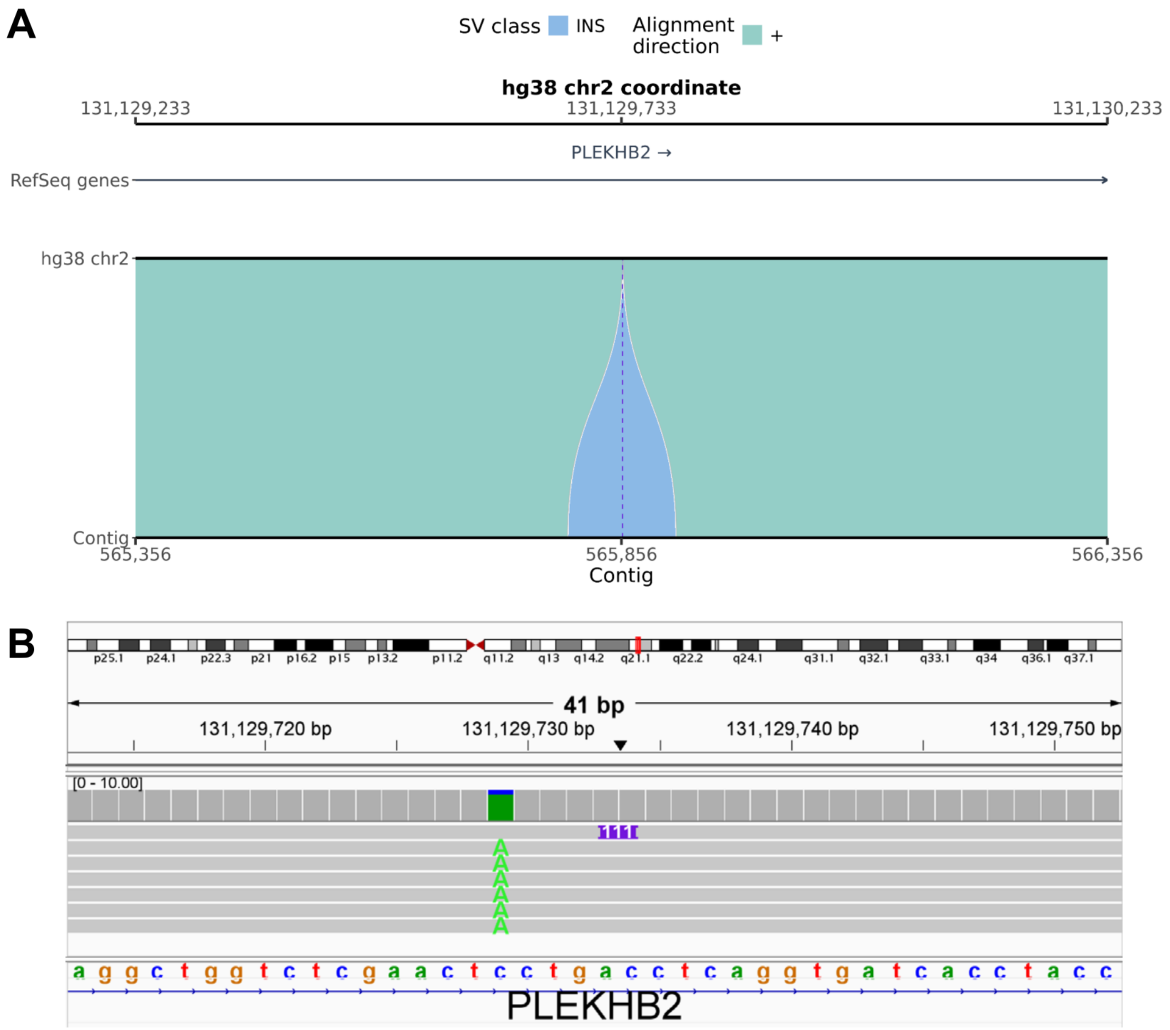


**Figure S43. svbyeye and IGV visualization of the 111-bp insertion at chr2:131,129,733.**

**(A)** Alignment of the hg38 reference sequence (top track) with a representative contig from an assembly of a sample carrying the SV (bottom track). Shaded blocks show the alignment and highlight the structural difference. The legend indicates SV class and alignment direction. Gene IDs are shown if present. The axes give reference and contig coordinates, respectively. **(B)** Read alignment at the SV locus. Coverage is shown above the reads. The SV is indicated with the SV length in purple. The hg38 reference bases are shown, with gene IDs if present.

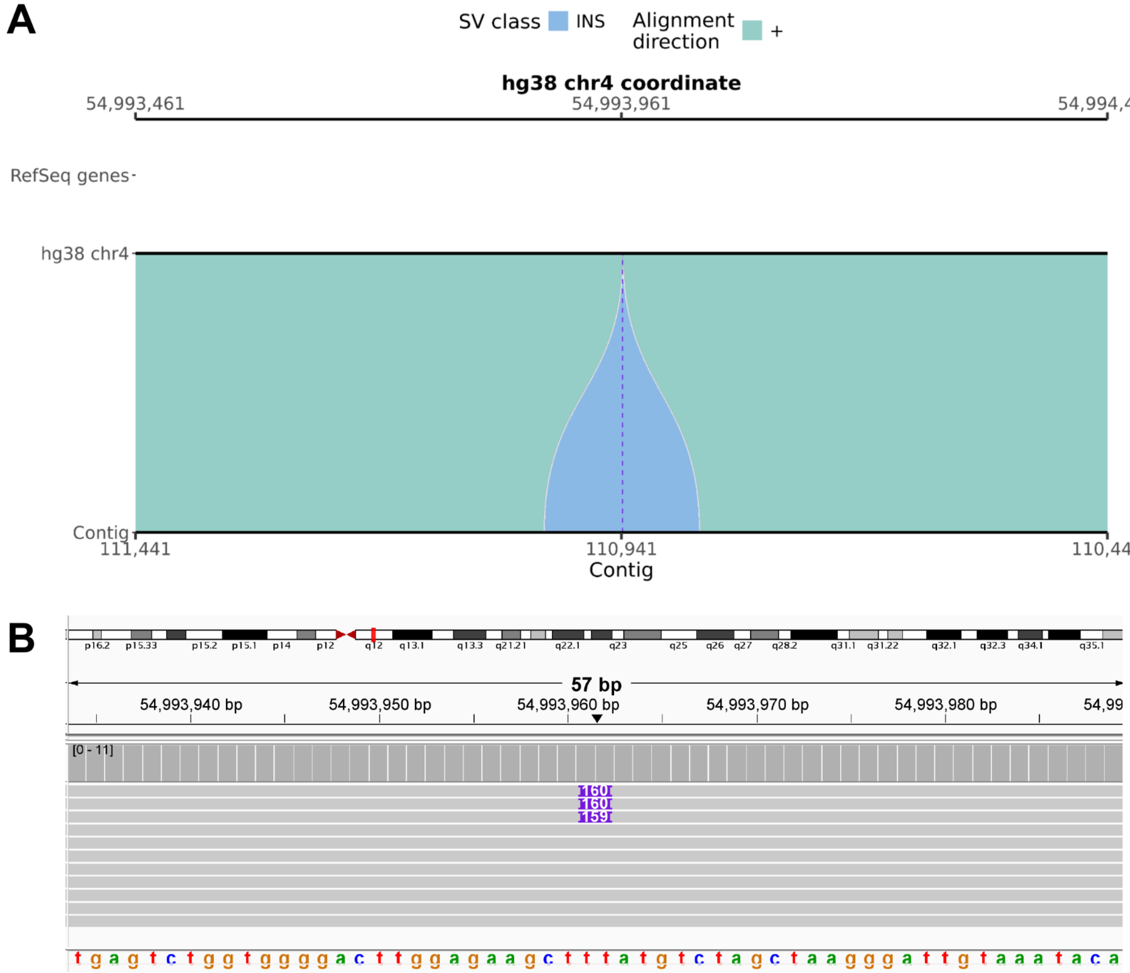


**Figure S44. svbyeye and IGV visualization of the 160-bp insertion at chr4:54,993,961.**

**(A)** Alignment of the hg38 reference sequence (top track) with a representative contig from an assembly of a sample carrying the SV (bottom track). Shaded blocks show the alignment and highlight the structural difference. The legend indicates SV class and alignment direction. Gene IDs are shown if present. The axes give reference and contig coordinates, respectively. **(B)** Read alignment at the SV locus. Coverage is shown above the reads. The SV is indicated with the SV length in purple. The hg38 reference bases are shown, with gene IDs if present.

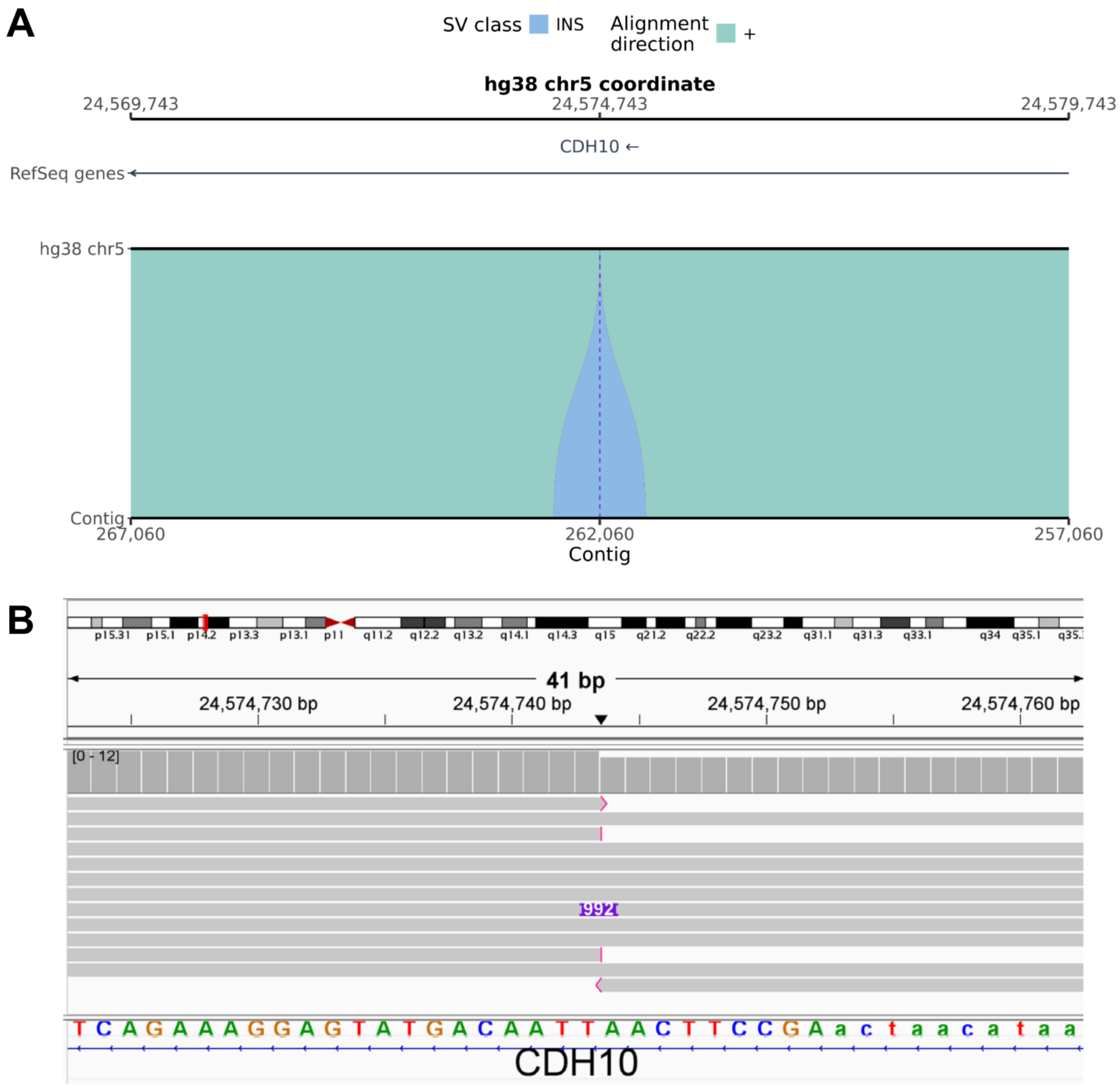


**Figure S45. svbyeye and IGV visualization of the 985-bp insertion at chr5:24,574,743.**

**(A)** Alignment of the hg38 reference sequence (top track) with a representative contig from an assembly of a sample carrying the SV (bottom track). Shaded blocks show the alignment and highlight the structural difference. The legend indicates SV class and alignment direction. Gene IDs are shown if present. The axes give reference and contig coordinates, respectively. **(B)** Read alignment at the SV locus. Coverage is shown above the reads. The SV is indicated with the SV length in purple. The hg38 reference bases are shown, with gene IDs if present.

A

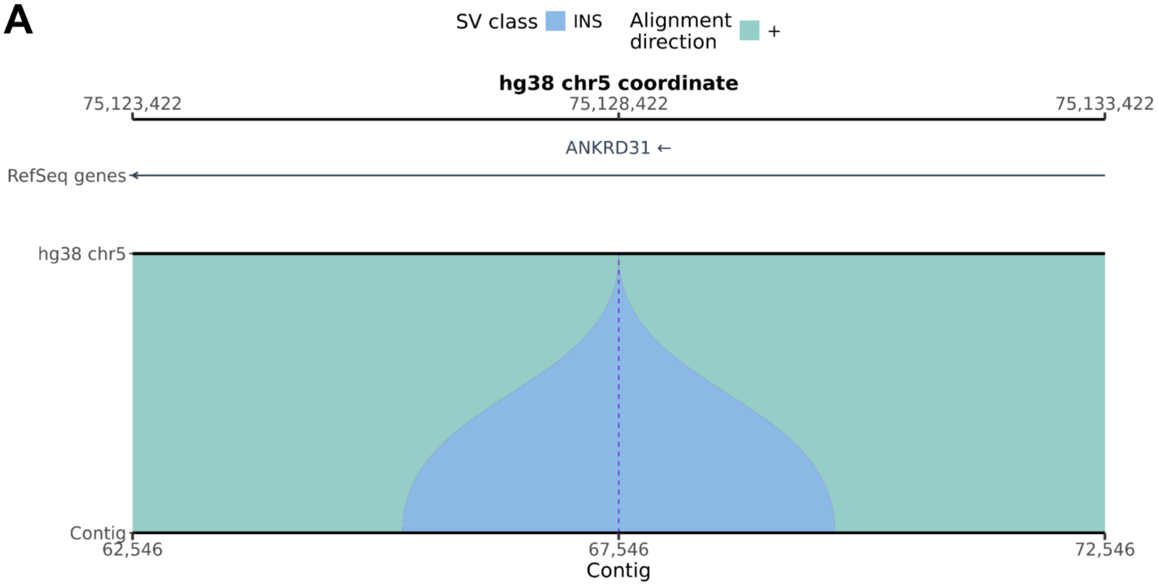


B

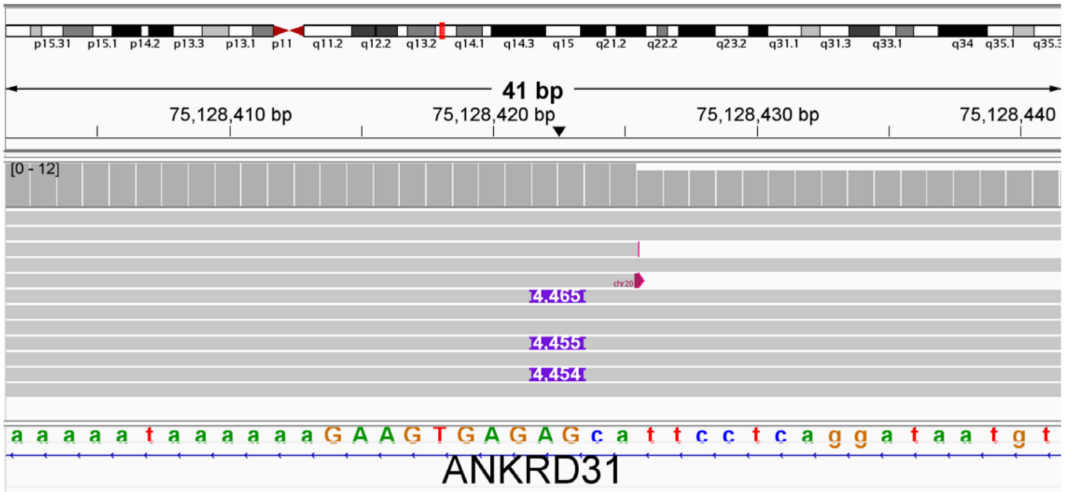


**Figure S46. svbyeye and IGV visualization of the 4,455-bp insertion at chr5:75,128,422.**

**(A)** Alignment of the hg38 reference sequence (top track) with a representative contig from an assembly of a sample carrying the SV (bottom track). Shaded blocks show the alignment and highlight the structural difference. The legend indicates SV class and alignment direction. Gene IDs are shown if present. The axes give reference and contig coordinates, respectively. **(B)** Read alignment at the SV locus. Coverage is shown above the reads. The SV is indicated with the SV length in purple. The hg38 reference bases are shown, with gene IDs if present.

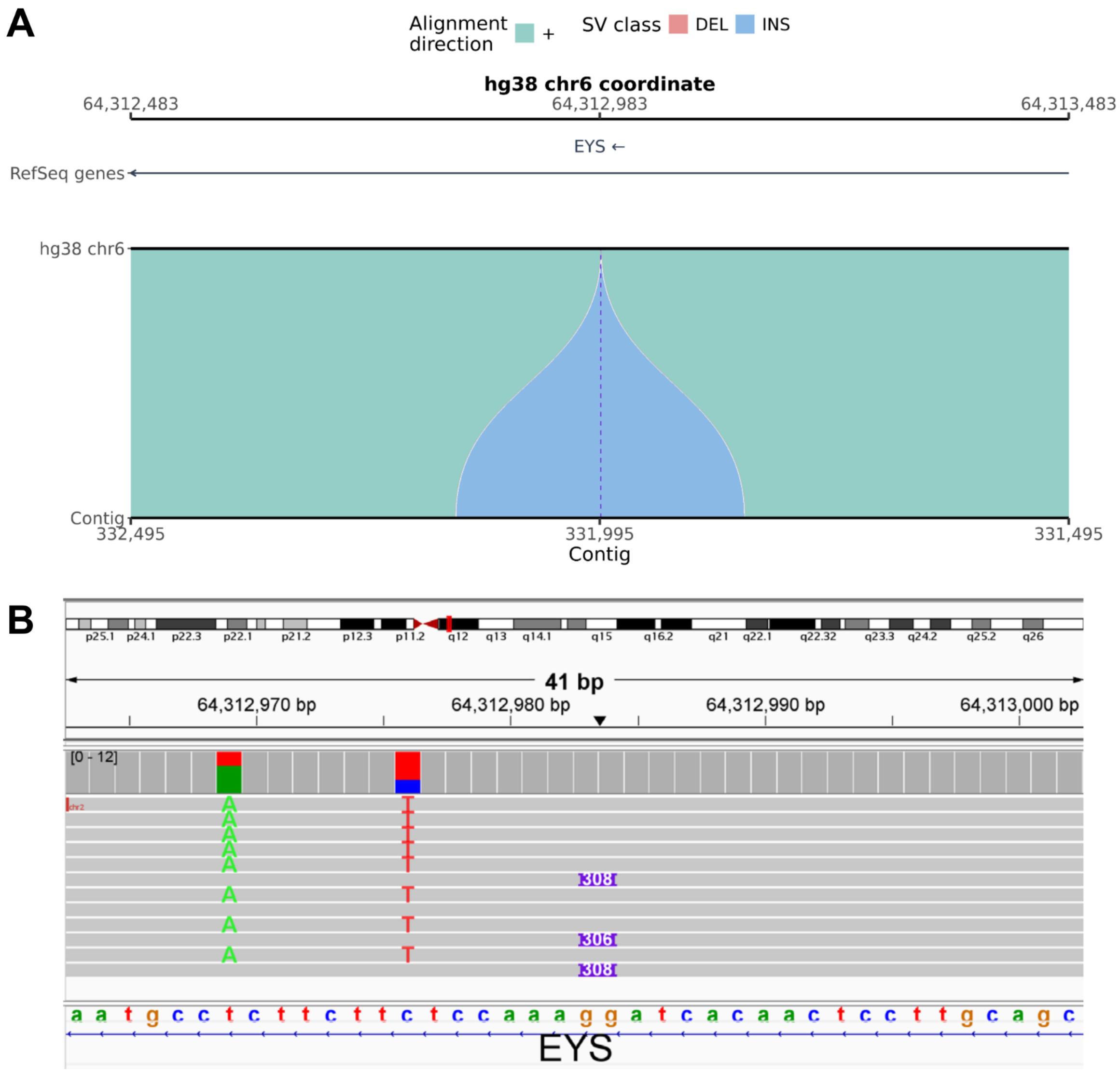


**Figure S47. svbyeye and IGV visualization of the 308-bp insertion at chr6:64,312,983.**

**(A)** Alignment of the hg38 reference sequence (top track) with a representative contig from an assembly of a sample carrying the SV (bottom track). Shaded blocks show the alignment and highlight the structural difference. The legend indicates SV class and alignment direction. Gene IDs are shown if present. The axes give reference and contig coordinates, respectively. **(B)** Read alignment at the SV locus. Coverage is shown above the reads. The SV is indicated with the SV length in purple. The hg38 reference bases are shown, with gene IDs if present.

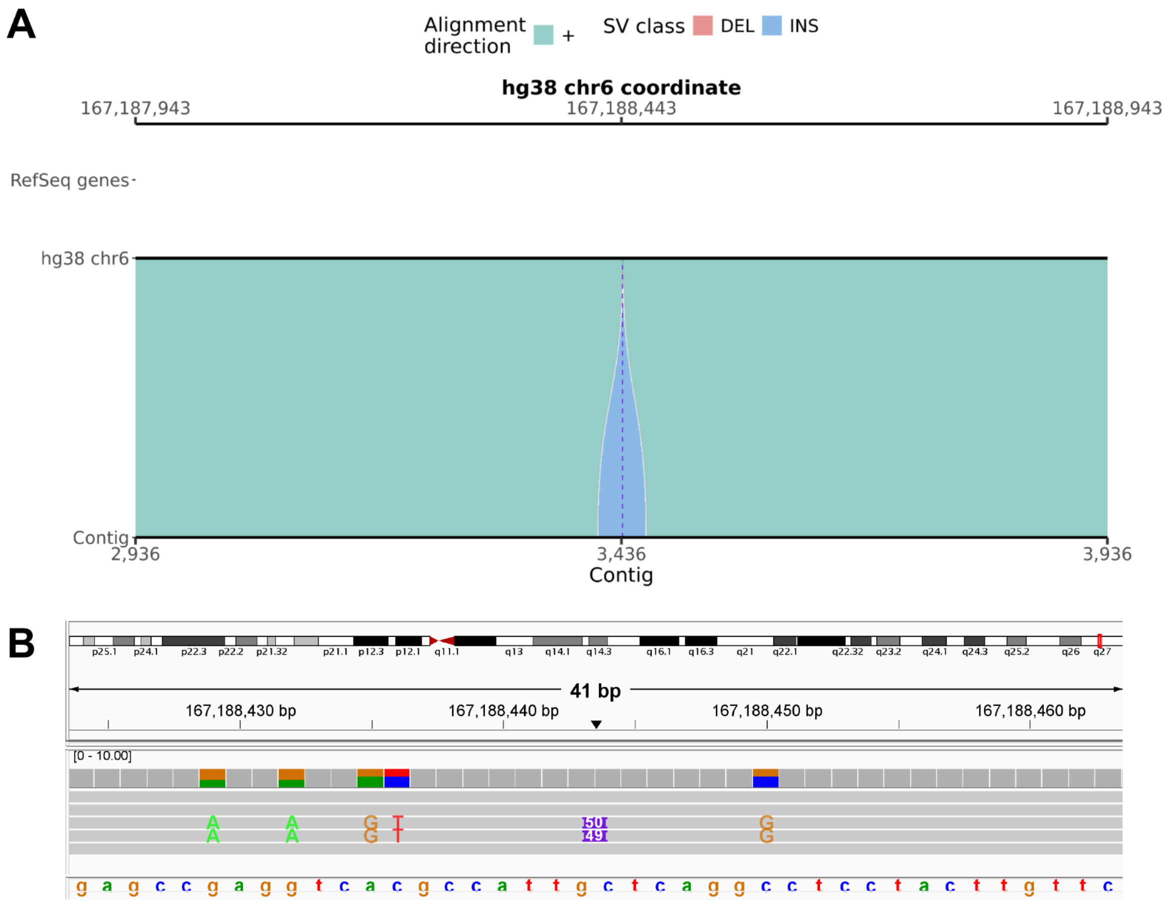


**Figure S48. svbyeye and IGV visualization of the 50-bp insertion at chr6:167,188,443.**

**(A)** Alignment of the hg38 reference sequence (top track) with a representative contig from an assembly of a sample carrying the SV (bottom track). Shaded blocks show the alignment and highlight the structural difference. The legend indicates SV class and alignment direction. Gene IDs are shown if present. The axes give reference and contig coordinates, respectively. **(B)** Read alignment at the SV locus. Coverage is shown above the reads. The SV is indicated with the SV length in purple. The hg38 reference bases are shown, with gene IDs if present.

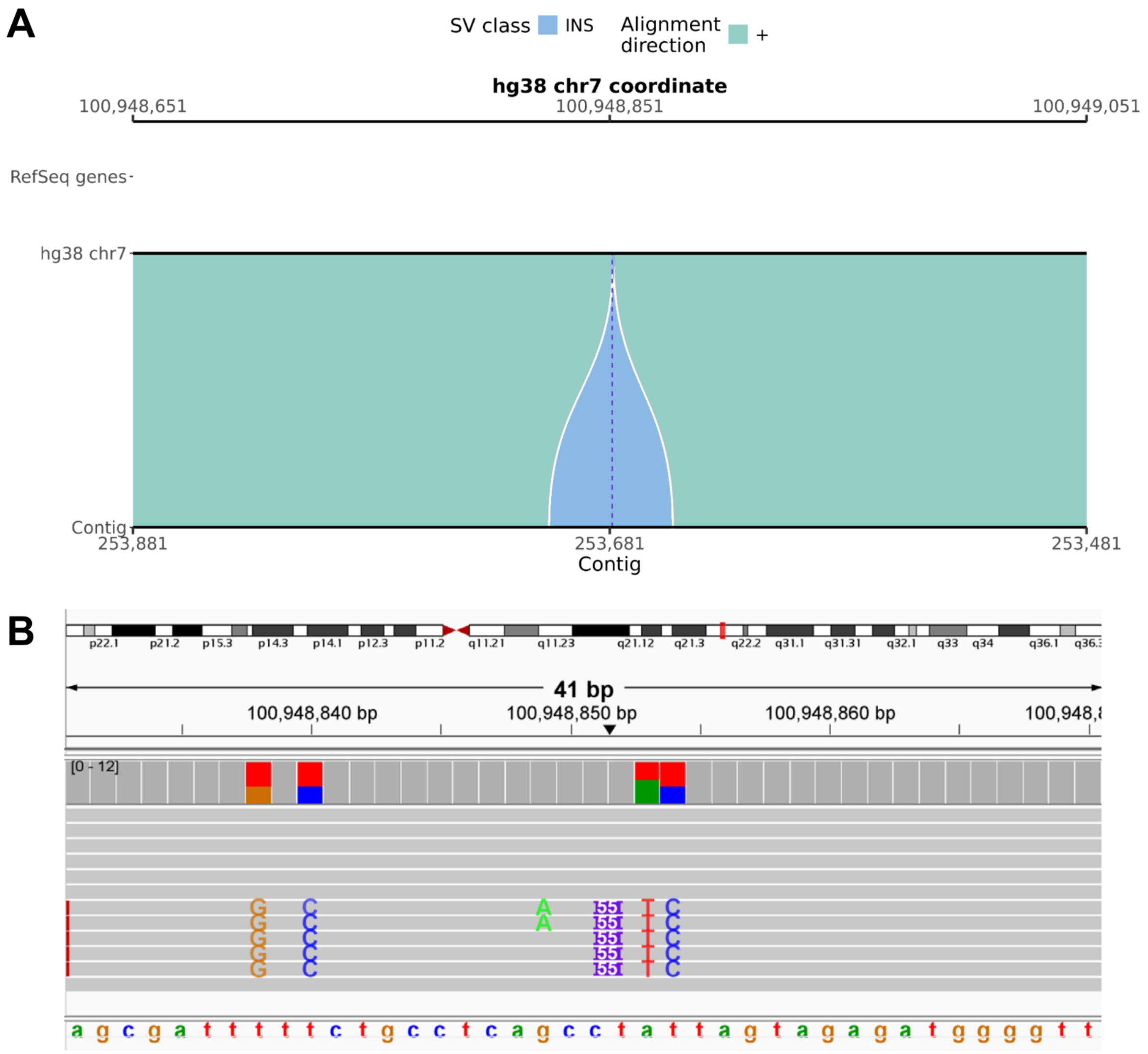


**Figure S49. svbyeye and IGV visualization of the 55-bp insertion at chr7:100,948,851.**

**(A)** Alignment of the hg38 reference sequence (top track) with a representative contig from an assembly of a sample carrying the SV (bottom track). Shaded blocks show the alignment and highlight the structural difference. The legend indicates SV class and alignment direction. Gene IDs are shown if present. The axes give reference and contig coordinates, respectively. **(B)** Read alignment at the SV locus. Coverage is shown above the reads. The SV is indicated with the SV length in purple. The hg38 reference bases are shown, with gene IDs if present.

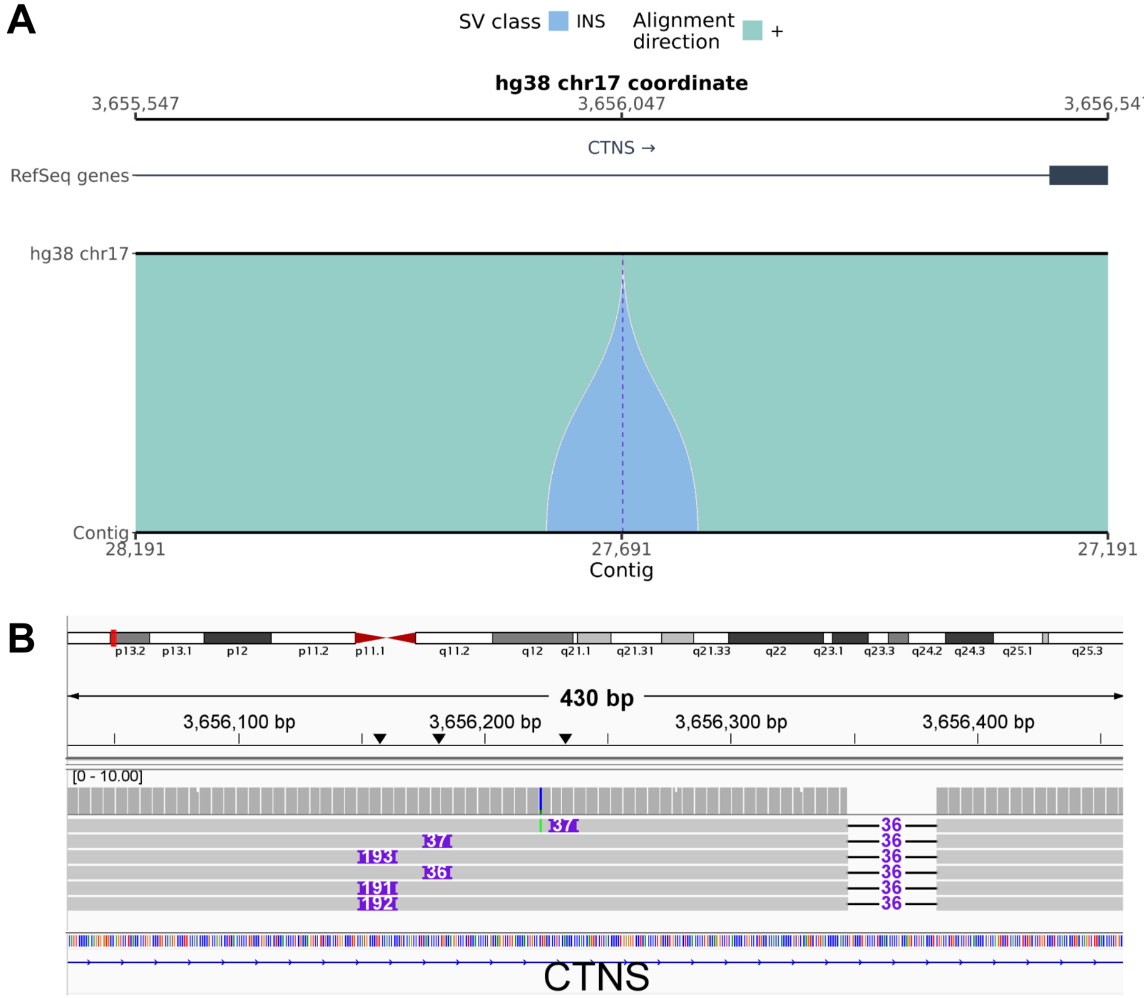


**Figure S50. svbyeye and IGV visualization of the 156-bp insertion at chr17:3,656,047.**

**(A)** Alignment of the hg38 reference sequence (top track) with a representative contig from an assembly of a sample carrying the SV (bottom track). Shaded blocks show the alignment and highlight the structural difference. The legend indicates SV class and alignment direction. Gene IDs are shown if present. The axes give reference and contig coordinates, respectively. **(B)** Read alignment at the SV locus. Coverage is shown above the reads. The SV is indicated with the SV length in purple. The hg38 reference bases are shown, with gene IDs if present.

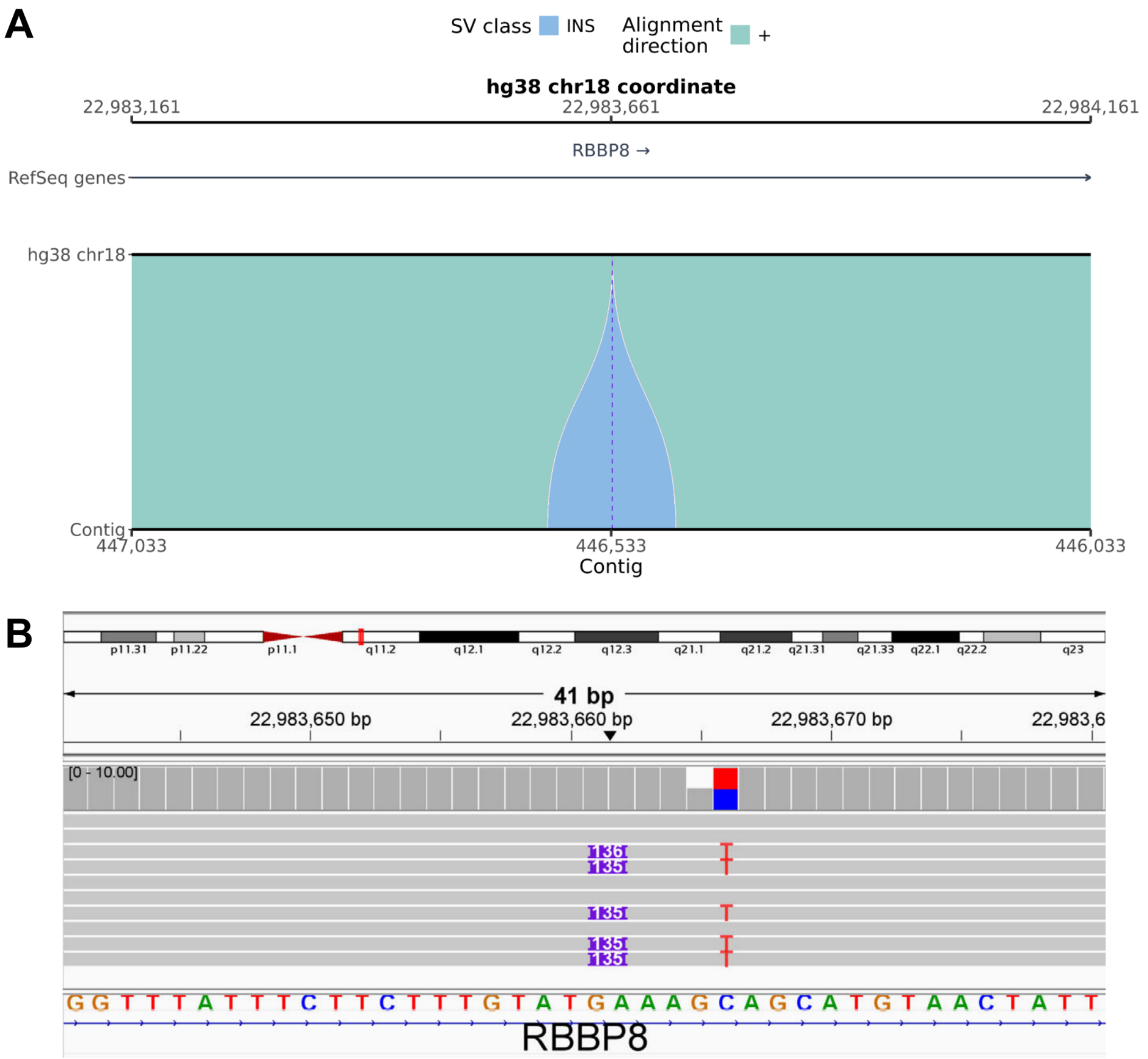


**Figure S51. svbyeye and IGV visualization of the 135-bp insertion at chr18:22,983,661.**

**(A)** Alignment of the hg38 reference sequence (top track) with a representative contig from an assembly of a sample carrying the SV (bottom track). Shaded blocks show the alignment and highlight the structural difference. The legend indicates SV class and alignment direction. Gene IDs are shown if present. The axes give reference and contig coordinates, respectively. **(B)** Read alignment at the SV locus. Coverage is shown above the reads. The SV is indicated with the SV length in purple. The hg38 reference bases are shown, with gene IDs if present.

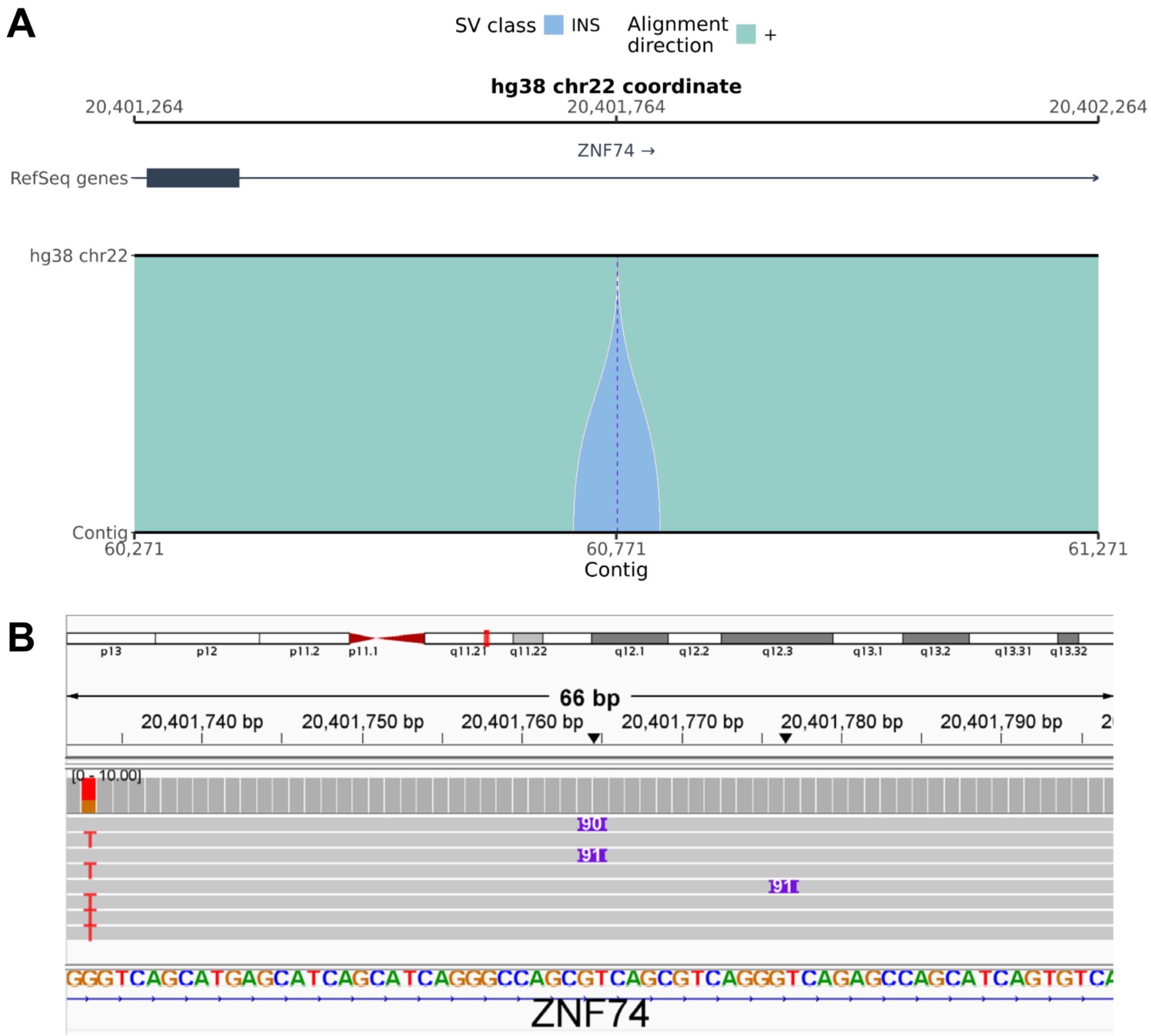


**Figure S52. svbyeye and IGV visualization of the 90-bp insertion at chr22:20,401,764.**

**(A)** Alignment of the hg38 reference sequence (top track) with a representative contig from an assembly of a sample carrying the SV (bottom track). Shaded blocks show the alignment and highlight the structural difference. The legend indicates SV class and alignment direction. Gene IDs are shown if present. The axes give reference and contig coordinates, respectively. **(B)** Read alignment at the SV locus. Coverage is shown above the reads. The SV is indicated with the SV length in purple. The hg38 reference bases are shown, with gene IDs if present.

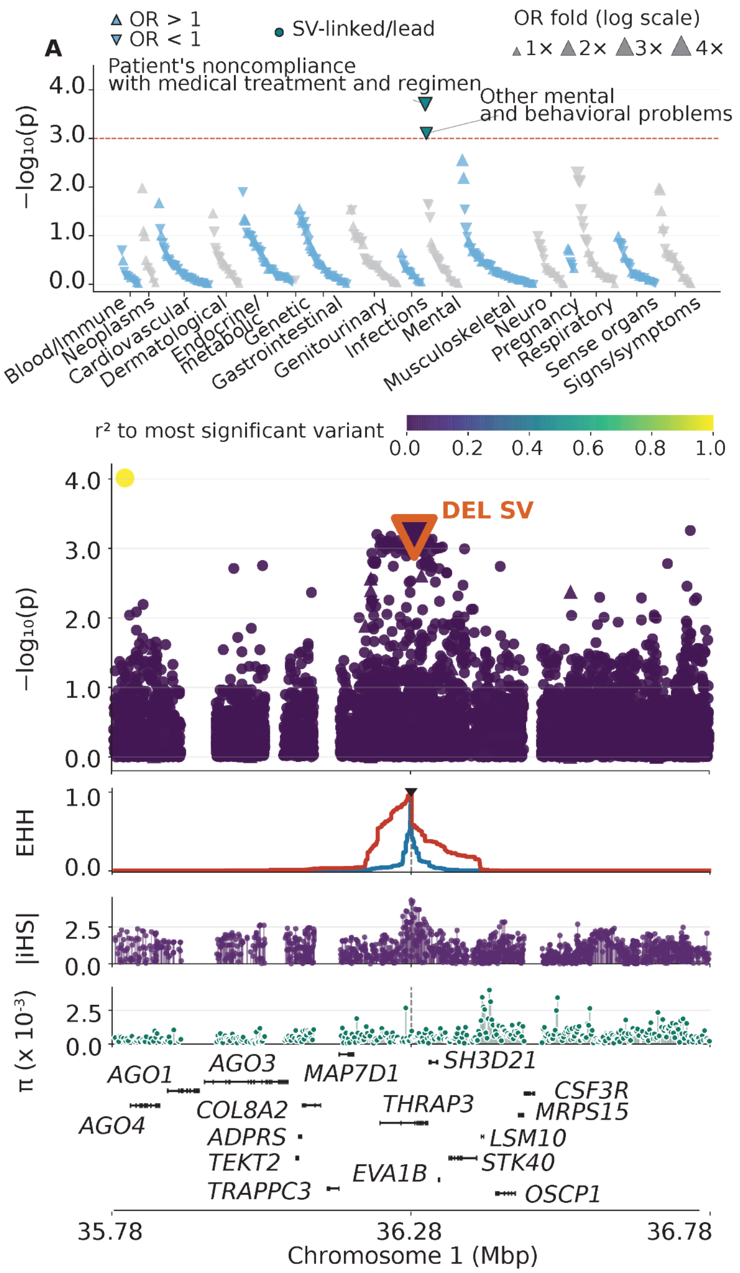


**Figure S53. Fine-mapping of the PheWAS association (chr1:36277076 C>G) for "other mental and behavioral problems" with linked 52-bp deletion locus at chr1:36,283,616.** (A) PheWAS Manhattan plot across 447 phecodes in 16 categories. Upward and downward triangles represent increased and decreased odds for individual phecode outcomes, respectively. Teal highlights the SV-linked or SV-lead association; yellow indicates nominal discovery significance. (B) Extended haplotype homozygosity (EHH) centered at the selection index variant, absolute normalized iHS scores, nucleotide diversity ($\pi$), and gene locations. Blue and red EHH curves indicate reference and alternative alleles, respectively. (C) Variant association signals across the locus, colored by linkage disequilibrium ($r^2$) with the most significant variant. Gene locations are indicated. Red-outlined triangles mark the SV in B and C.

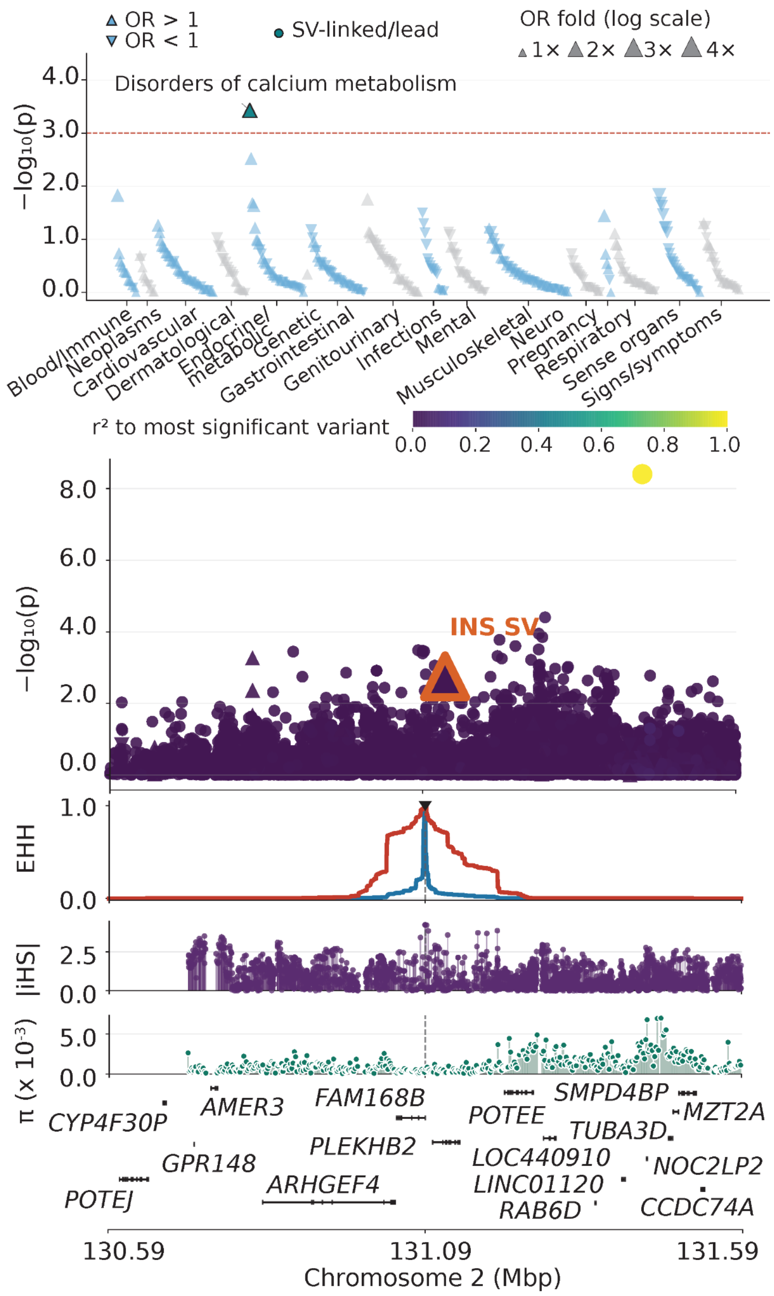


**Figure S54. Fine-mapping of the PheWAS association (chr2:131094186 A>G) for "disorders of calcium metabolism" with linked 111-bp insertion locus at chr2:131,129,733.** (A) PheWAS Manhattan plot across 447 phecodes in 16 categories. Upward and downward triangles represent increased and decreased odds for individual phecode outcomes, respectively. Teal highlights the SV-linked or SV-lead association; yellow indicates nominal discovery significance. (B) Extended haplotype homozygosity (EHH) centered at the selection index variant, absolute normalized iHS scores, nucleotide diversity (π), and gene locations. Blue and red EHH curves indicate reference and alternative alleles, respectively. (C) Variant association signals across the locus, colored by linkage disequilibrium ($r^2$) with the most significant variant. Gene locations are indicated. Red-outlined triangles mark the SV in B and C.

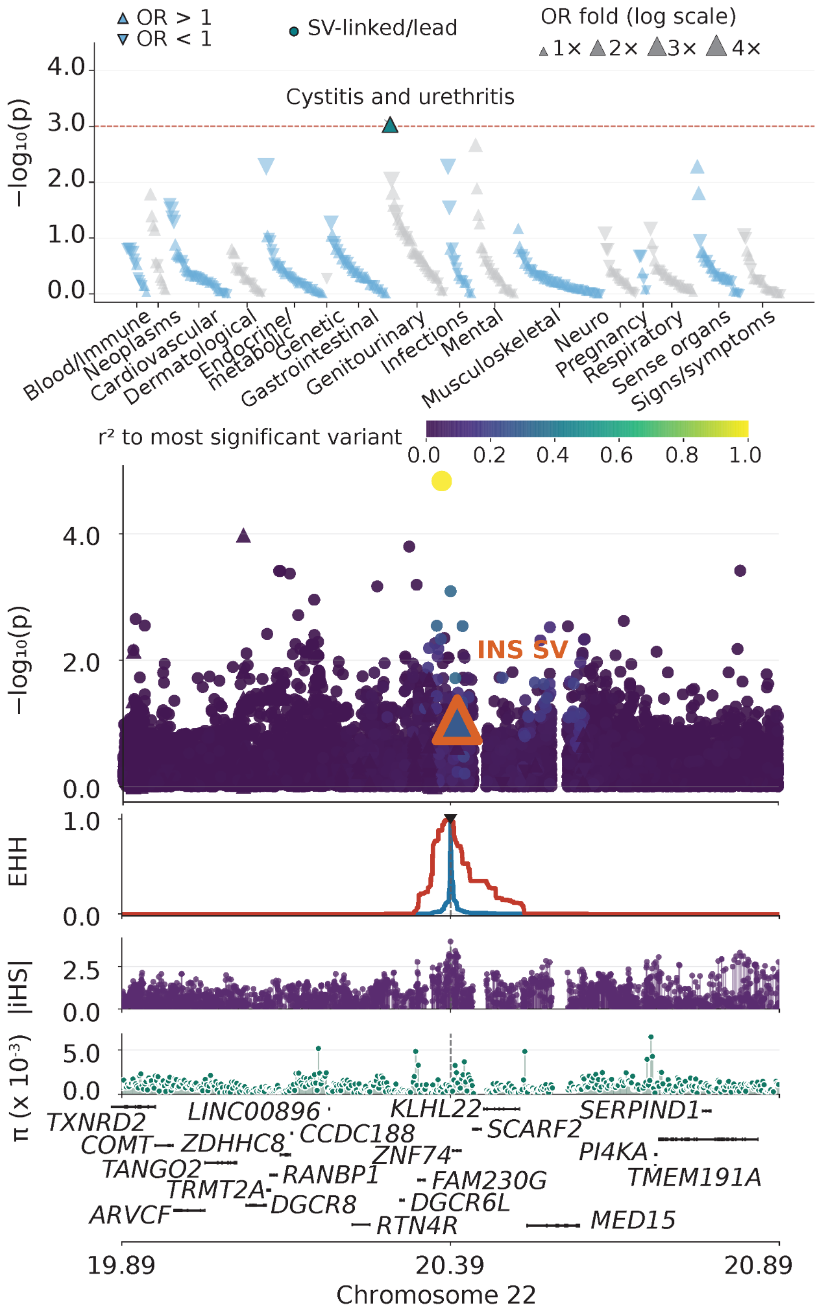


**Figure S55. Fine-mapping of the PheWAS association (chr22:20391674 T>C for "cystitis and urethritis" with linked 90-bp insertion locus at chr22:20,401,764.** (A) PheWAS Manhattan plot across 447 phecodes in 16 categories. Upward and downward triangles represent increased and decreased odds for individual phecode outcomes, respectively. Teal highlights the SV-linked or SV-lead association; yellow indicates nominal discovery significance. (B) Extended haplotype homozygosity (EHH) centered at the selection index variant, absolute normalized iHS scores, nucleotide diversity (π), and gene locations. Blue and red EHH curves indicate reference and alternative alleles, respectively. (C) Variant association signals across the locus, colored by linkage disequilibrium (r²) with the most significant variant. Gene locations are indicated. Red-outlined triangles mark the SV in B and C.
